# Defining drivers of human papillomavirus (HPV) vaccine uptake in migrant populations globally and strategies and interventions to improve coverage: a systematic review

**DOI:** 10.1101/2025.01.31.25321303

**Authors:** Michiyo Iwami, Oumnia Bouaddi, Mohammad S Razai, Rania Mansour, Beatriz Morais, Nafeesa Mat Ali, Alison F Crawshaw, Sainabou Bojang, Farah Seedat, Anna Deal, Sophie Webb, Jessica Carter, Nathaniel Aspray, Nuria Sanchez Clemente, Juan Arroyo-Laguna, Sanjeev Krishna, Yolanda Augustin, Henry M Staines, Sally Hargreaves

**Author notes:** Joint first authors (Michiyo Iwami, Oumnia Bouaddi). Joint last authors (Yolanda Augustin, Henry M Staines, Sally Hargreaves). **Corresponding author:** Prof Sally Hargreaves. City St George’s, University of London, United Kingdom, SW17 0EA.

## Abstract

**Background:** The Cervical Cancer Elimination Initiative by the World Health Organization (WHO) has set a target of 90% human papillomavirus (HPV) vaccination coverage among girls by age 15 by 2030 to dramatically reduce deaths from cervical and other HPV-related cancers. However, progress has been slow, with only 27% global coverage in 2023. Migrants are considered an under-immunised group globally for many vaccine-preventable diseases, with data showing that they may experience a high burden of HPV infection and widespread HPV under-immunisation. Better understanding of the factors influencing the ability of these communities to get vaccinated for HPV is important. We aimed to systematically synthesise evidence on drivers of HPV vaccination uptake in migrants, and explored recommended approaches, strategies, and best practices to promote uptake in these communities.

**Methods:** We searched seven databases (e.g., Medline, Global Health) and websites (WHO, IOM, Google Scholar) for literature on drivers of HPV vaccination uptake among migrants globally, published between January 2006 and December 2024 in any language. Data on influencing factors for HPV vaccination uptake in migrants were extracted for an integrated approach to synthesising findings, and recommended strategies to improve it were compiled. We conducted a hybrid thematic analysis using the WHO BeSD model and assessed risk of bias with Joanna Briggs Institute checklists. PROSPERO protocol: CRD42023401694.

**Findings:** We identified 1,806 database records and 1,756 records from websites, ultimately including 117 studies with 5,638,836 participants across 16 countries and one territory (including 933,187 first- and second-generation migrants, mostly defined as foreign-born in high-income countries). Factors negatively influencing vaccine uptake included concerns about vaccine safety, cultural beliefs, uncertainty about HPV vaccines/infection, low knowledge of HPV/HPV vaccine, gender/sex, inter-generational and family dynamics, exposure to negative information, and lack of recommendations from healthcare providers. Practical barriers included limited information on services, language issues combined with a lack of skilled interpreters, logistical challenges, and the high cost of the vaccine. Enablers mainly included positive perceptions and trust in the vaccine and healthcare providers, realistic expectations from parents regarding the sexual activity of adolescents, a sense of responsibility, as well as recommendations from healthcare providers and support from social networks. Other positive predictors of vaccine uptake included being female, and having a history of vaccine-preventable diseases or abnormal Pap test results. Findings highlighted that free-of-charge and school-based schemes were effective in increasing uptake, while mandatory or optional schemes were less popular. Key recommended approaches included culturally sensitive messaging and tailored communication for different target groups (e.g., parents/caregivers, adolescents), with an emphasis on strength framing. Deploying trusted mediators (e.g., peer school health promoters, religious champions, community health workers) and implementing practical solutions to address missed opportunities (e.g., bundling HPV vaccination with other services) and for mobile migrants (e.g., eHealth) were also emphasised. Additionally, strong provider recommendations and reducing access barriers through measures including walk-in, mobile, and outreach services were recommended, alongside addressing broader cross-cutting issues, such as strengthening vaccine monitoring systems.

**Interpretation:** This review showed that migrants worldwide face complex individual, family/social, and provider/system-level barriers to HPV vaccination, resulting in missed opportunities for protection. In many low- and middle-income countries (LMICs), the vaccine is either unavailable or has to be paid for. Achieving global commitments for universal and equitable immunisation across the life-course, making progress toward cervical cancer elimination, requires addressing these barriers through multi-pronged strategies. This includes combining effective health communication to build trust and address negative perceptions, along with efforts to eliminate physical barriers to vaccine access. Given the lack of data from LMICs, future research must urgently explore specific drivers of HPV vaccination among migrants in these regions where they are more concentrated and access to the HPV vaccine is limited, as well as develop solutions to system-level problems. Collaborative efforts with migrant communities are essential to co-develop effective, tailored delivery models that meet their unique needs.

**Funding:** This research was funded by the NIHR (NIHR300072), the Academy of Medical Sciences (SBF005\1111), and the Medical Research Council (MRC/N013638/1).

## Introduction

Human papillomavirus (HPV) causes multiple cancers (e.g., cervical, oropharyngeal, vaginal, penal, anal^1^) and genital warts. For example, HPV is responsible for over 95% of cervical cancer cases globally,^2^ which has been shown to be preventable through screening and vaccination.^3^ In 2022, there were over 662,301 new cases of cervical cancer worldwide, with approximately 94% associated deaths occurring in low- and middle-income countries (LMICs).^2,4^ HPV vaccination is a primary prevention strategy that has been employed since 2006. Various HPV vaccines (bi-/quadri-/nona-valent) have been developed to prevent different HPV-associated cancers and are reported to be safe and highly effective.^5^

The World Health Organization (WHO) prioritises all girls aged 9-14 to receive HPV vaccination before becoming sexually active^2^ and has set a global target of 90% of girls receiving HPV vaccination by the age of 15 by 2030.^6^ However, progress towards this target has been slow. In 2023, global HPV vaccination coverage (first dose) in girls was estimated at 27%,^7^ with LMICs lagging significantly behind. In addition, less than 25% of low-income countries have introduced HPV vaccination into their Essential Programmes on Immunization (EPI) by 2022.^8^ A new low-dose vaccine is anticipated to accommodate vaccination for other population groups such as boys, the elderly, and girls aged 9-14 in low-income countries not in school, and is a method to increase vaccine supply among LMICs. Globally, out of 141 HPV vaccination programmes, only 43 countries and 4 territories are reported to have universal vaccines (gender-neutral/pan-gender, for both girls and boys); and all are in high-income countries (HICs) or upper middle-income countries apart from Bhutan.^9^

Migrants (defined by International Organization for Migration [IOM] as any individuals who move away from their usual place of residence between or within a country^10^) are considered disproportionately vulnerable to HPV infection and its associated cancers. In Europe, studies in Southern and Central Italy have reported significantly higher HPV infection rates and incidence of invasive cervical cancer among migrants compared to native Italians, making them a priority group for HPV vaccine interventions.^11,12^ In one study, migrants residing in Italy were reported to be 3.5 times more likely to get cervical cancer compared to native Italians.^13^ Another study showed that the incidence of cervical cancer declined in native Italian women by 2.7% on a yearly average, but increased by 12.2% among migrants during the same period.^14^

Despite global calls for equitable and universal access to life-course immunisation,^15^ migrants (including refugees and asylum seekers) are considered an under-immunised group for many vaccine-preventable diseases.^16^ This is due to missed vaccines, doses, and boosters, the lack of availability of certain vaccines in their countries of origin,^17,18^ and well-documented barriers to routine and catch-up vaccination.^19,20^ Existing literature also demonstrates disparities in HPV vaccination access and coverage among migrants compared to host communities globally.^21^ A recent global systematic review among 31,442 migrants and refugees reported low HPV vaccination initiation and completion (defined as a total of three doses), 31.6% and 34.5%, respectively, with disparities by sex, regions (higher coverage rates in Europe compared to the Americas and Western Pacific), and migration status.^22^ Similarly, a recent systematic review in the United States of America (USA) reported low HPV initiation and completion (30% and 14%, respectively) among children of migrant parents,^23^ and that foreign-born individuals were 38% less likely to receive HPV vaccination compared to native populations.^24^ The situation is comparable in Europe, where a significant difference in HPV vaccination completion was reported between UK-born natives and Polish-born migrants (87.2-89.8% versus 69.7-77.2%).^25^

The low vaccine coverage in migrant groups, below internationally set targets, and in comparison to host communities, and multi-level barriers often experienced by migrants suggest complex barriers to HPV vaccination uptake unique to this population group need to be explored. Multiple studies have investigated this issue; however, no comprehensive effort has been made to synthesise this information to identify the drivers of HPV vaccination uptake in migrants worldwide. Such synthesis is key for identifying intervention targets aimed at increasing vaccine uptake and achieving cervical cancer elimination as well as global HPV vaccination goals.^6^ We thus aimed to systematically synthesise first, the evidence on drivers of HPV vaccination uptake in migrants—using WHO’s Behavioural and Social Driver of Vaccination Framework (BeSD)^26^—and second, the recommended approaches, strategies, and best practices to promote uptake among this population.

## Methods

This systematic review was guided by the PRISMA guidelines 2020.^27^ The protocol for this review has been registered in PROSPERO (CRD42022347513).^28^

### Inclusion and exclusion criteria

Eligibility criteria were developed using the PICOS framework (**Table S1**). We included primary studies published in any language from the inception of HPV vaccine delivery (i.e., 2006 in the USA^29^) onwards. We focused on ‘migrants’ defined as foreign-born nationals, and included first- and second-generation migrants. These studies reported on factors influencing HPV vaccine uptake among adolescent and adult migrants, children of migrant parents eligible for HPV vaccination programmes, and other stakeholders, including healthcare providers. We included qualitative and quantitative cross-sectional studies, cohort studies, and randomised controlled trials (RCTs). Primary outcomes included factors positively/negatively influencing HPV vaccination, as well as recommended approaches, strategies, and best practices reported by authors/participants. Comparison groups (i.e., host communities) were included where possible. Studies were excluded if they did not disaggregate data for migrants, meet our definition of a migrant, or report on factors influencing HPV vaccination.

### Search strategy and screening

We searched seven databases (Medline, Embase, APA PsycINFO, Global Health, CINAHL, Scopus; and Cochrane Database of Systematic Reviews and Trials) for records published between 2006 and the search dates (March 2022, updated in March 2023, and again in December 2024) in any language and country. The search combined free text and subject heading terms for migrant, vaccination, and HPV separated by Boolean operators (**Table S2** for full search strategy). We also performed an extensive grey literature search, including websites of relevant international organisations (WHO, GAVI, IOM, UNHCR, ReliefWeb, Refworld) and Google Scholar. We also hand-searched reference lists of relevant systematic reviews identified from the above. All records were uploaded onto Covidence.^30^ Duplicates were removed, and three reviewers (MI, MSR, and RM) performed title/abstract screening, and full-text review. Any disagreements between the reviewers were resolved through discussion and corroborated by a senior researcher (SH).

### Data extraction

Two reviewers (MI, MSR) performed data extraction using a pre-defined form which was piloted and refined. The extracted information included: study characteristics (study design, country of study, year of study, setting, etc); participant characteristics (number of participants, participant type, gender/sex, age, nativity, country of origin, race/ethnicity, migratory status, etc); aim(s); methodology (data collection and analysis methods, participant recruitment); outcomes (factors negatively/positively influencing uptake); recommendations (by participants/authors). Any discrepancies were resolved by consensus with input from the senior author (SH).

### Risk of bias assessment

Three reviewers (MI, RM, OB) independently performed the risk of bias assessment using Joanna Briggs Institute (JBI) Critical Appraisal tools.^31^ Each study type was assessed using the corresponding JBI checklist.^31^ Mixed-methods studies were appraised using a combination of both the qualitative and cross-sectional quantitative JBI checklists. Each item within the checklists was rated as ‘yes’ (score 1), ‘no’ or ‘not sure’ (score 0), or ‘not applicable’ (excluded from the total item count). The risk of bias in each study was presented as the mean percentage of ‘yes’ scores. Studies scoring below 60% were considered at high risk of bias, those scoring 60-80% were considered at moderate risk of bias, and those scoring 80% and over were considered at low risk of bias. No studies were excluded based on risk of bias assessments, but the results were considered in the qualitative synthesis of the results and in discussion.

### Data analysis and synthesis

We performed hybrid thematic analysis^32,33^ using the WHO-BeSD as an *a priori* framework to systematically organise and structure the data synthesis.^26^ This framework contains four domains which influence uptake of recommended vaccines: ‘what people think and feel’; ‘social processes’; ‘motivation’; and ‘practical issues’ (**Figure 1**). MI identified preliminary codes which were then grouped as sub-themes mapped onto *a priori* constructs of the BeSD framework and refined iteratively. Where it was not possible to map onto any *priori* constructs, new domains were added. All results were refined and validated by another researcher (OB) and the senior author (SH).

**Figure 1.**
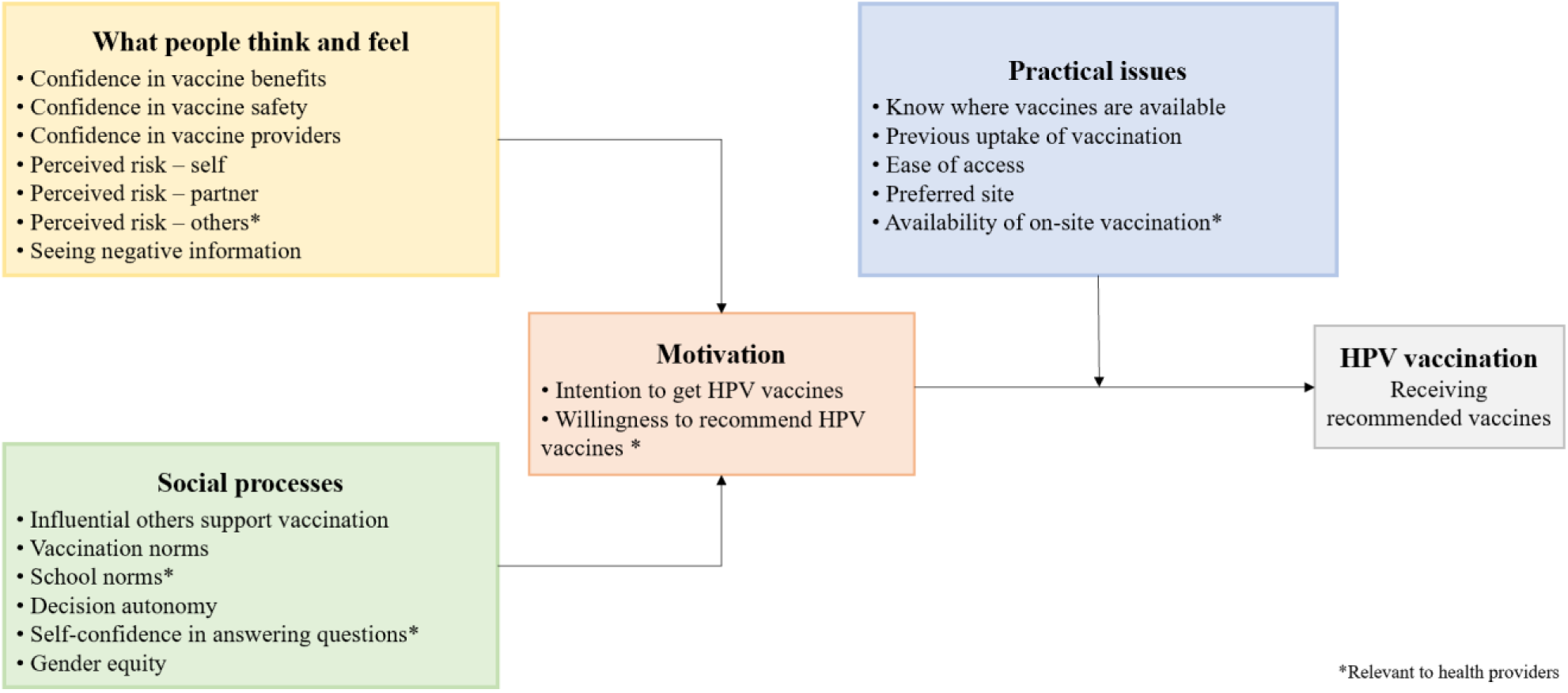
The WHO Behavioural and Social Drivers of Vaccination Uptake Framework.

## Results

### Overview of included studies

A total 3,562 records were retrieved, of which 2,340 underwent title and abstract screening. After full-text review, 117 studies involving 5,638,836 participants across 16 countries and one territory (including 933,187 first- and second-generation migrants) were included in the final analysis (**Figure 2**).^34–150^ Most studies were from HICs, with 66% (n=77/117) conducted in the USA or US territory,^34,35,47,54,57–61,63–65,67,68,70–74,76,78,79,81,84,86–89,94–96,98–119,121,122,125–128,130–133,135,137–146,148–150^ with the remainder primarily in Europe (24%; n=28/117),^36–38,40–46,48–50,53,55,56,69,75,77,80,83,90–93,120,123,124^ particularly Scandinavian countries (57%; n=16/28).^37,38,40–46,48–50,53,56,93,123^ Only one study was conducted in a LMIC (Nepal).^97^ See **Figure 3** for study location map. Only one study was conducted in the WHO Eastern Mediterranean Region,^136^ and none in Latin America or Africa. Few studies reported specific migrant status, such as refugee (n=13),^42,51,52,63,66,71,85,97,101,105,120,129,143^ student (n=7),^60,65,136,142,145,148,150^ or migrant farmworker (n=3),^68,76,86^ but the majority of studies either provided no or unclear information. The most frequent countries of origin were Mexico (n=17),^47,61,64,68,81,84,86–89,105,109,110,113,121,125,130^ followed by China (n=12)^39,41,60,90,104,115,122,129,135,145,147,150^ and Somalia (n=9),^35,41,42,53,57,58,78,116,143^ and most represented ethnic groups were Hispanic^35,63,68,72,84,98–101,103,105,109,113,126,132,133,138,142^ or Latina/o/x (n=32),^54,59,63,73,76,96,101,103–105,114,117,119,121,127,135,137,142^ White, including non-Hispanic/Latino White (n=27),^35,47,68,73,84,90,98–100,104,109,112–115,119,126,127,132,133,135,138,139,142,143,148,150^ Asian (n=22),^35,51,73,100,104,109,112–115,119,122,127,132,135,138,139,142,143,148–150^ Black, including non-Hispanic Black (n=21),^35,57,78,84,98–100,103,109,110,113,119,126,127,129,132,139,142,143,148,150^ and African American (n=16).^59,63,74,78,96,101,103–105,112,115,118,132,133,135,150^ See **Figure S1** for a visual depiction of country/region of origin and race/ethnicity. The studies included various participant groups, including vaccine recipients only (n=34),^39,41,42,48,49,52,59,60,65,66,78,81,84,98,99,102,110–112,115,118,119,122,124,132,133,136,139,141–143,145,148,150^ parents/caregivers only (n=21),^36,57,63,64,68,69,75,86,88,94–96,100,101,105,106,125,128,129,138,149^ Mother only (n=17),^34,47,58,71,74,79,82,87,89,97,107,121,131,135,137,140,144^ Father only (n=1),^67^ and healthcare providers only (n=3).^62,72,73^ The majority of studies (n=79) were considered at low risk of bias,^36,38,40–43,45–49,51–53,56–82,87,90–94,96–100,102,108–117,119,122–124,126–128,131–134,137,138,140,142,149,32^ studies were considered at moderate risk of bias,^35,37,39,44,50,54,55,83–86,89,95,101,103–105,118,120,121,125,129,130,135,136,139,141,143–146,148^ and only six studies were considered at high risk of bias.^34,88,106,107,147,150^ See **Table 1** and **Table S3** for further details.

**Figure 2.**
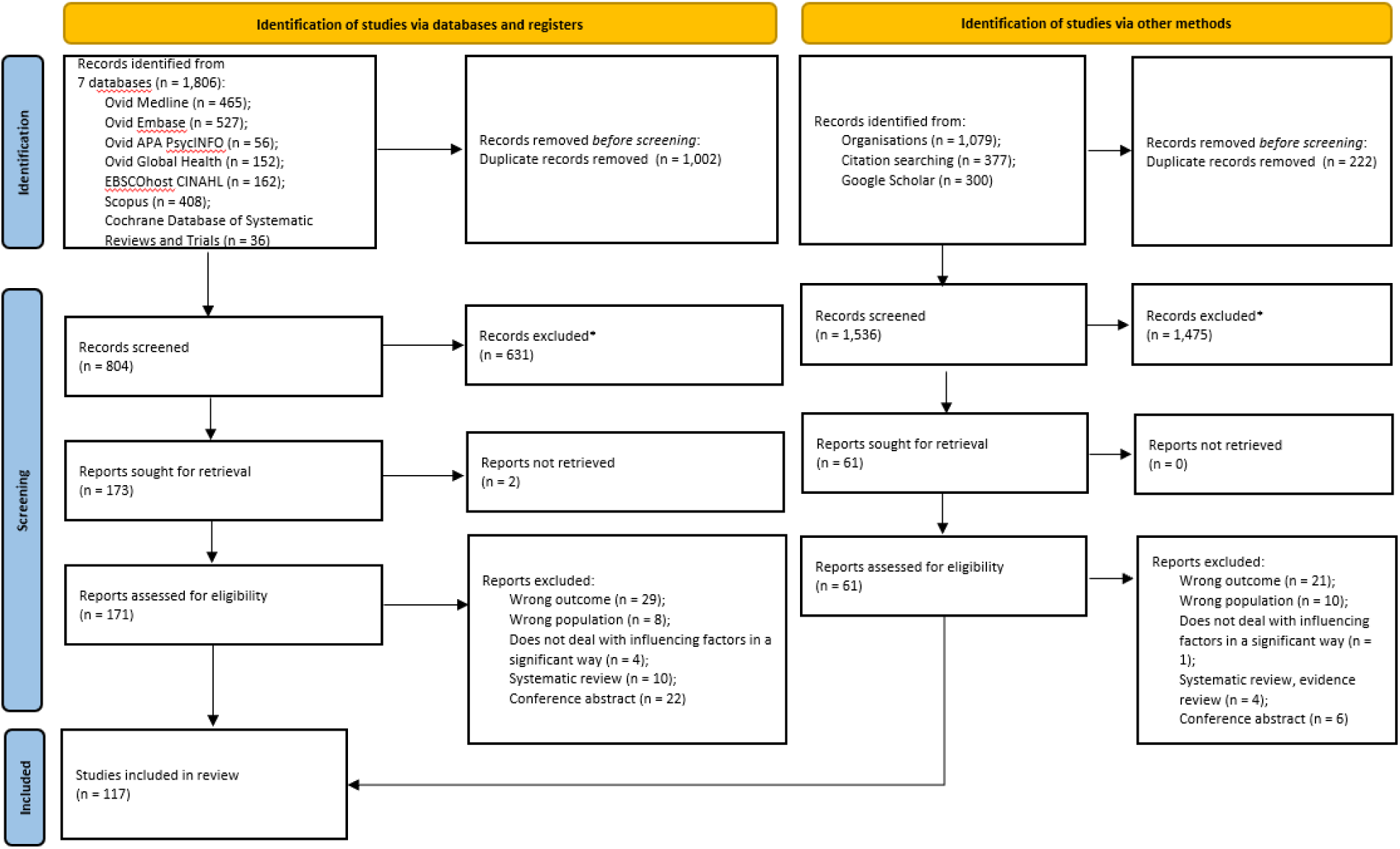
PRISMA Flow Chart.

**Figure 3.**
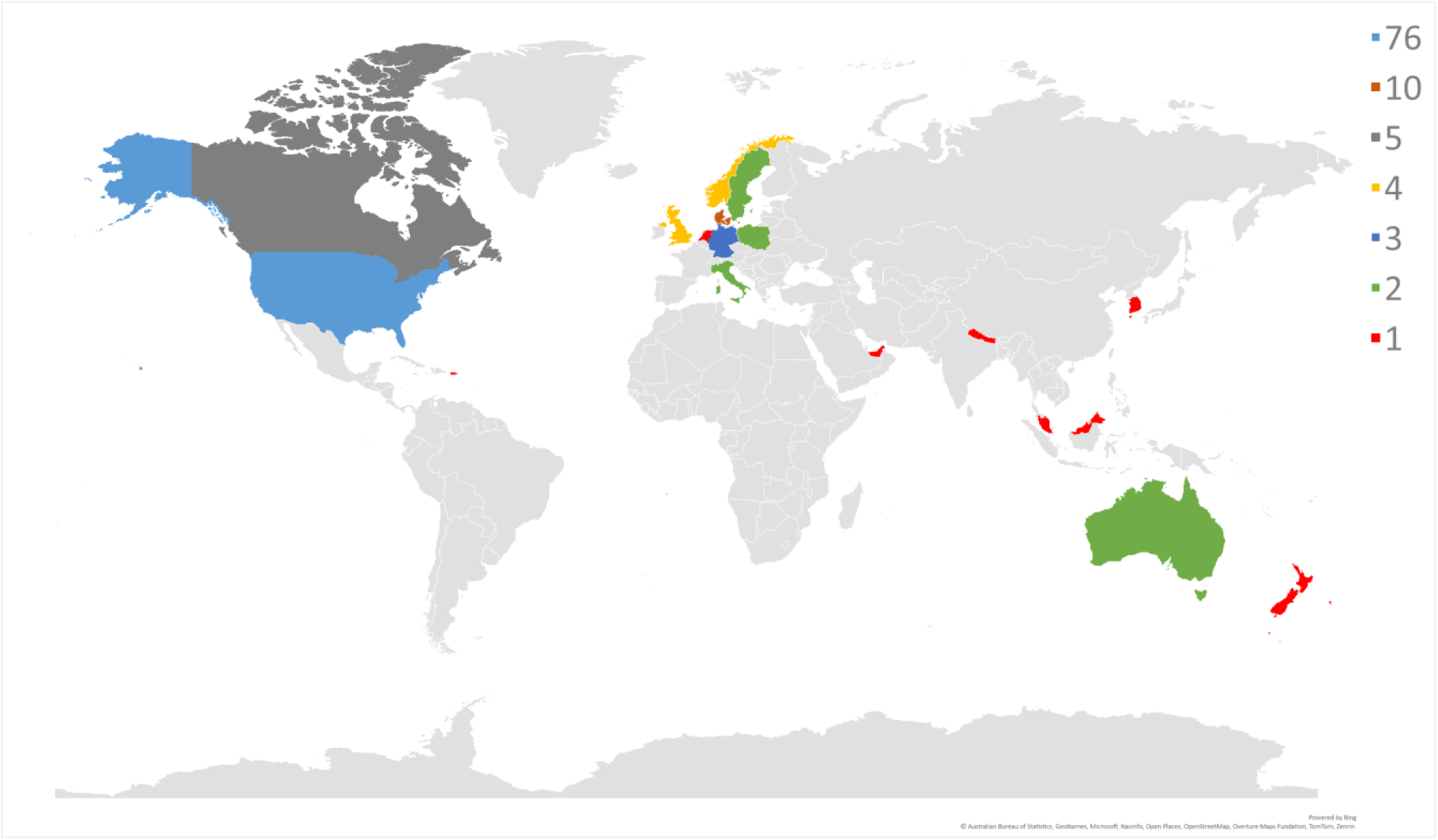
Map of Study Location.

### Drivers of HPV vaccination: Thematic analysis using the BeSD framework

The BeSD framework was adapted based on the findings (**Figure 4** and **Table S4**). Domains 5 (*Programme design and delivery modes*) and 6 (*Socio-demographic and other factors*) emerged from the data and were added to the framework. We present the findings separately for each domain below.

**Figure 4.**
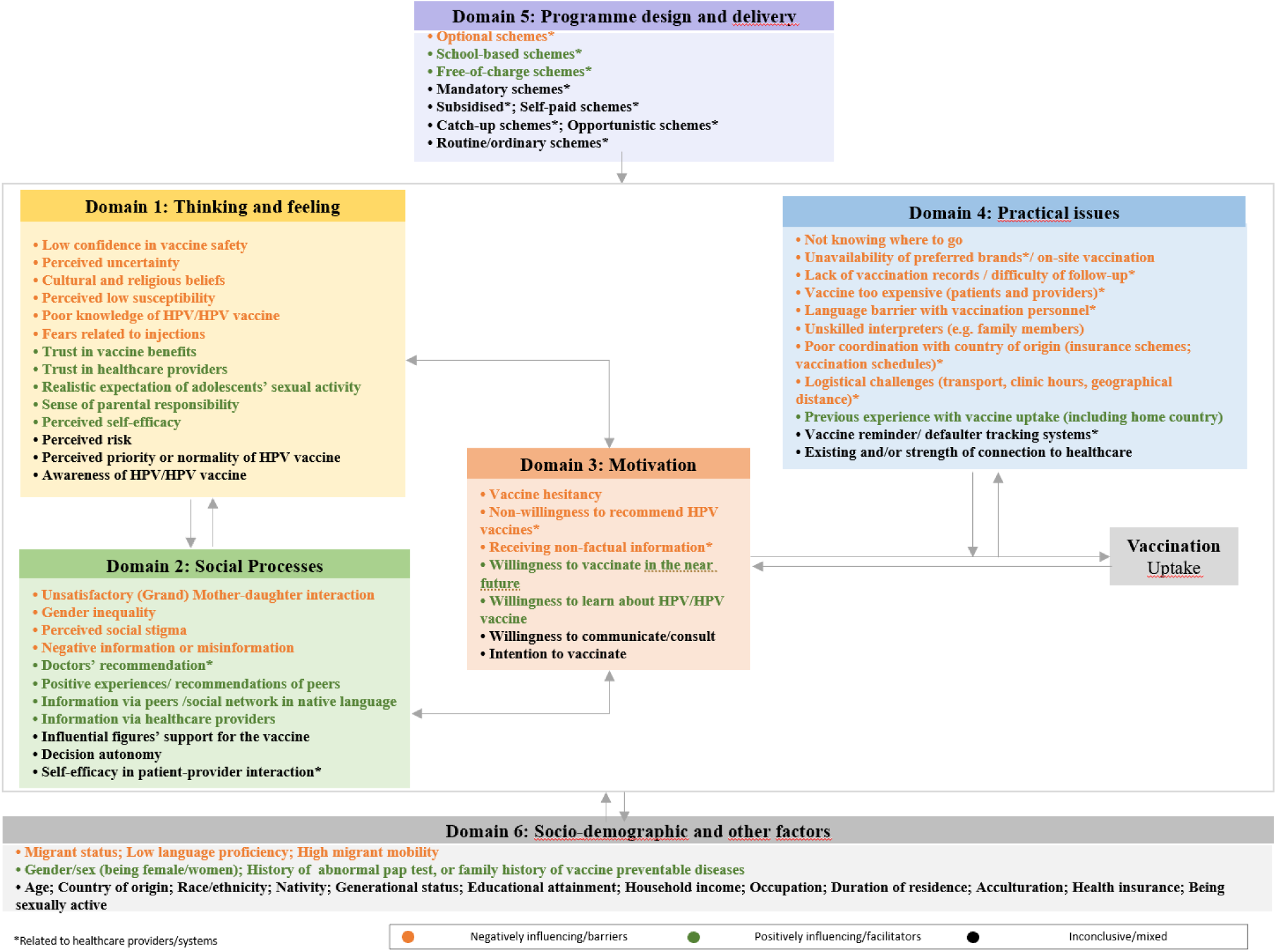
Behavioural and Social Drivers of HPV Vaccination Uptake in Migrants.

**Table 1.** Overview of Characteristics of Included Studies (N=119)

| Category | Details |
| --- | --- |
| **Study Design** | Quantitative, cross-sectional [n=62]; Qualitative [n=28]; Quantitative, cohort [n=16]; Mixed-methods [n=10]; RCT [n=1] |
| **Year of Publication** | 2007 [n=1]; 2009 [n=1]; 2010 [n=2]; 2011 [n=3]; 2012 [n=5]; 2013 [n=8]; 2014 [n=3]; 2015 [n=7]; 2016 [n=8]; 2017 [n=9]; 2018 [n=13]; 2019 [n=8]; 2020 [n=7]; 2021 [n=13]; 2022 [n=13]; 2023 [n=11]; 2024<sup>†</sup> [n=5]. |
| **Countries** | USA/US territory [n=76, plus Puerto Rico n=1]; Denmark [n=10]; Canada [n=5]; Norway [n=4]; UK [n=4, including England n=2, and Scotland n=1]; Germany [n=3]; Sweden [n=2]; Poland [n=2]; Italy [n=2]; Australia [n=2]; The Netherlands [n=1]; New Zealand [n=1]; Republic of Korea (South Korea) [n=1]; Nepal [n=1]; Malaysia [n=1]; United Arab Emirates [n=1] |
| **WHO Region** | AMR [n=82]; EUR [n=28]; WPR [n=5]; EMR [n=1]; SEAR [n=1] |
| **Country Classification by Income Level** | High income [n=115]; Upper Middle Income [n=1]; Low Middle Income [n=1] |
| **Study Setting** | Community setting only [n=48]; Household setting only [n=37]; Clinic setting only [n=10]; College/university setting only [n=8]; Both Community & Clinic [n=7]; Both Clinic & Household [n=2]; Multi-level/systems setting [n=2]; Camp [n=2]; Both Clinic & High school [n=1] |
| **Participant Groups** | Recipient of vaccine/vaccine eligible only [n=34]; Parents/caregiver/guardian only [n=21]; Mother only [n=17]; Father only [n=1]; Healthcare provider only [n=3]; Other stakeholders [n=1];<br>Combination of stakeholders [n=40]: {Recipient of vaccine/vaccine eligible and Parent/caregiver (including mother only) [n=34]; Parent/caregiver (including mother only) and Healthcare provider [n=3]; Recipient of vaccine/vaccine eligible and Regional coordinators [n=1]; Recipient of vaccine/vaccine eligible, Parent/caregiver (including mother only), and Healthcare provider [n=2]} |
| **Number of Participants** | N=5,638,836 |
| **Gender/sex of participants** | Female only [n=51]; Both gender [n=49]; Both gender or female only (depending on the group) [n=13]; Male only [n=3]; Not available [n=1] |
| **Residence** | Urban/super urban [n=48]; Nation-wide [n=33]; Mixed [n=15]; Rural [n=5]; Suburban [n=2]; Not clear [n=14] |
| **Number of Migrants<sup>††</sup>** | N=933,187 (Mean 61.1% [SD=37.6]) |
| **Migrant Type** | Refugee [n=13]; Student [n=7]; Labour migrant, e.g., migrant farmworker [n=3], economic migrant [n=1]; Not specified/Not clear [n=93] |
| **Risk of Bias** | Low risk [n=79]; Moderate risk [n=32]; High risk [n=6] |

<sup>†</sup>Up to 4 December 2024.
<sup>††</sup>This includes immigrants/first-generation and descendants/second-generation migrants.
AMR: Americas; EUR: Europe; WPR: Western Pacific; EMR: Eastern Mediterranean; SEAR: South-East Asia.

#### Domain 1: Thoughts and feelings about HPV vaccination

Main factors negatively influencing uptake included concerns around vaccine safety,^34–36,47,56–59,61,64,67,68,70,71,74–76,79–81,128,129,140,142,144,145,147,149,150^ cultural and religious beliefs (e.g., perceived risk of premarital sex or promiscuity, concerns about pork gelatine),^34–36,55,57,61,62,66–70,73,74,76,78,79,81,107,116,128,140,144^ uncertainty around HPV vaccines and infection (often linked to perceived needs for more information),^34–36,55,57–62,66,68,70,71,74,80,81,96,107,116–118,120,128,129,147,149^ parents/caregivers thoughts that vaccination was unnecessary because their daughters are too young or because it was only relevant within the context of marriage,^34–36,55,57,60,62,65–67,70,74,78,81,85,107,128,129,144^ and limited knowledge about HPV/HPV vaccine^34–36,56,57,59–62,64–67,69–71,74,76,78–81,83,85,86,100,101,107,113,116,118,120,122,128,129,134,136,140,142,145,147^.

Positively influencing factors included a sense of parental responsibility for getting their eligible child vaccinated,^36,47,55,56,66,68,71,76,79,81,96,144^ parents’ more realistic understanding of children’s sexual activity,^64,71,73,96^ especially among Latinx parents,^64^ self-efficacy and confidence in one’s ability to engage in preventive action,^64,66,118,121,142^ confidence in vaccine benefits,^34,35,58,59,61,66,67,69–71,76,79,80,85,96,118,140,142^ and trust in healthcare providers.^34,35,62,67,73,74,76,82,129,140^ The influence of perceived risks of HPV infection/associated cancers to one’s child^36,47,54,61,62,71,74,76,121,137^ or oneself^36,54,60,66,75,78,117,121,137^ showed mixed results, and one’s partners were rarely investigated or voiced.

#### Domain 2: Social processes

Factors impeding HPV vaccine uptake included tense or unsatisfactory (grand)mother-daughter interactions (including spouse communication) and relationships (often resulting in ineffective communication about child vaccination and sexual health),^34–36,55–58,66,71,74,78,79,81,82,85,140,149^ as well as gatekeeping behaviours of grandmothers regarding health-seeking for daughters and daughters-in-law.^80^ These poor relationships were reported to be more common in second-generation adolescent migrants, where a mismatch in communication exists along with differential peer effects, often leading to a clash of norms between mothers and daughters.^56^ Overall, perceived social stigma was reported as a key negative influencing factor, often rooted in cultural histories and negative past experiences (e.g., unethical experimentation practices during the HIV epidemic among Haitians and historical relations between Black Americans and physicians).^34,118^ Social stigma was also rooted in heightened embarrassment, as HPV vaccination was perceived to be linked to promiscuity or to sexually transmitted infections (STIs).^144,150^ However, one study found that greater knowledge was linked to reduced stigma around HPV infection and vaccination, particularly among international students.^60^

Gender (in)equality was also reported as a key factor, whereby power dynamics within families—especially strong paternal influence over maternal authority in health decision-making— was shown to complicate the vaccination process for daughters.^36^ Finally, negative information,^55,56,75,76,107^ and the spread of vaccine misconceptions or misinformation^35,36,66,76,80,81,107,140^ (via social media, mini-videos, or peers^81^) were consistently reported as deterrents to seeking HPV vaccination.

Facilitators of HPV vaccine uptake included hearing positive experiences or advice from those who had themselves or their children vaccinated,^47,57,67,73,79^ receiving information about the vaccine through doctors’ recommendations,^34,55,57,59,61–63,66,67,70,71,73,74,76,79,80,82,85,107,136,137,149^ and receiving information about the vaccine via healthcare providers/schools,^34,36,56,58,63,64,68,69,74,79,83,86,94,97,117,120,128,129,140,149^ or peers/social networks (in one’s native language).^47,55,56,58–60,63,64,67,70,72,75,76,79,80,82,97,120,140,149^ The impact of information received depended on multiple factors, including the strength and framing of recommendations, communication methods, the characteristics of both sources and recipients,^57^ and personal preferences regarding information formats (oral versus written, or both; direct versus indirect communication with clinicians).^68^

#### Domain 3: Motivation

Negatively influencing factors included hesitancy to vaccinate among parents/caregivers,^34,37,78,80,85,107,121,137,140,145^ limited willingness of providers to recommend HPV vaccines (often due to low perceived priority, competing priorities, or preconception about migrants’ cultural beliefs),^76,107^ or provision of non-factual information.^76^ Factors positively influencing uptake included parent/caregiver willingness to get oneself or their child vaccinated in the near future^34,64,66,67,71–73,75,77,80,83,85,106,107,118,120,137,138,140,149^ and to learn about HPV/HPV vaccines.^34,56,59,80,81^ Influences of intention to vaccine were inconclusive.^70,74,81,105,116,121,137,144,145^

#### Domain 4: Practical issues

This domain included more negatively influencing factors than other domains. Practical issues preventing uptake included migrants’ lack of knowledge about where vaccines are available,^62,76,80,103,116,120,128,140,150^ unavailability of preferred brands,^75^ unavailability of on-site vaccination,^61,68,76,77,82,107^ lack of vaccination records,^57,72,76,80,82^ and difficulty or loss to follow-up.

Affordability was another barrier, as vaccines were reported to be too expensive for beneficiaries and providers due to lack of funding.^59,61,62,66,72,76,77,80,81,103,118,128,140,147,150^ Poor access was reported due to multiple logistical challenges, including difficult transport, time constraints, inadequate clinic hours, and geographical distance.^61,62,68,71,76,80,81,85,128,129^ Poor coordination between host and home countries (e.g., regarding insurance, vaccine schedules) was also reported as a contributing factor to poor continuity of care.^68,75,76,80^

Language barriers between vaccination personnel and migrants posed another challenge, with unskilled interpreters sometimes providing insufficient or inaccurate information (e.g., young daughters acting as interpreters).^61,71,73,75,76,78,80–83,107,129^ Vaccine reminders or defaulter tracking systems showed inconsistent effects on promoting uptake.^45,68,76^

Facilitators included previous vaccination uptake (including in the home country)^37,45,56,67,68^ and the strength of connection to healthcare, such as having a usual place to seek care or the number of healthcare visits in the past year; however, the latter was not statistically significant in migrants,^98,99,108,137,146^ as opposed to non-migrants.^98,99^

#### Domain 5: Socio-demographic and other factors

Factors within this domain extended beyond health or vaccination-specific issues, yet are likely to influence uptake. However, they were largely mixed or inconclusive. Overall, significant predictors or critical factors of low uptake included migrant status,^61,76,80,142^ high migrant mobility,^43,61,68,69,71,72,75,76,81,82^ and low language proficiency of vaccine recipients and their parents (e.g. inability to understand the language used in host country, dialect-speaking migrant families).^61–63,65,71,75,76,78,80,83,102,107^ Predictors of high uptake included previous experience or family history with vaccine-preventable diseases (cervical cancer) or abnormal Pap test results.^47,67,72,73,76,96,140^ Gender/sex exhibited moderate effects on initiation, completion, and uptake of the HPV vaccine,^98,112,127,132^ but substantial effects on HPV/HPV vaccine awareness,^117,120^ with women generally performing better than men.

In contrast, duration of residence,^36,41,42,51,55,63,72,84,86,88,93,99,101,102,104,105,107,109,117,118,120,125,127,130,135,137–139,142^ acculturation,^56,58,60,63,71,74,87–89,94,95,117,126,130,142,147,149^ educational attainment,^49,51,56,73,76,85,86,94,98,99,107,108,110,117,120,121,123,134,136–138,142,144,149^ and household income^49,51,52,64,73,76,94,98,107,108,112,123,134,136–138,146,149^ showed inconsistent findings. Nativity, generational status, country/region of origin, and race/ethnicity also showed inconsistent results. They were also intertwined with gender/sex, with women generally showing higher initiation than men^98,110,112^ and immigrants showing lower uptake compared to descendants.^43^ Age exhibited inconsistent effects, with older individuals sometimes showing better initiation or uptake rates.^39,48,109^

#### Domain 6: HPV vaccination programme design and delivery modes

The studies included various HPV vaccine delivery approaches with differential effects on HPV vaccination. School-based programmes were among the most consistently effective approaches for uptake,^38–40,49,62,69,96,123^ with free school-based catch-up programmes showing the most favourable outcomes in terms of equitable HPV vaccine initiation compared to non-school-based free catch-up approaches and free school-based ordinary programmes.^40^ New data indicates school type is important. In one study, public schools demonstrated the best performance in HPV vaccination initiation, then private schools, whereas special need education schools performed poorly.^53^Another study also reported a lack of HPV vaccine information provision for an adolescent of an Arabic-speaking mother who attended a special need education school, resulting in no HPV vaccine uptake.^82^

Overall, free vaccination programmes showed consistently positive effects on HPV vaccine initiation in migrants,^38–41,43,44,49,76,93,117,129^ compared to self-payment schemes. Rates differed across migrant groups, duration of residence, generational status, income, and type of programme (catch-up versus routine). For example, migrant girls had significantly lower HPV vaccination uptake compared with native girls in both routine^43^ and catch-up programmes,^42^ but this difference only remained significant for routine vaccination when adjusting for household income.^42^ Generational status affected initiation rates in routine and free-of-charge catch-up programmes, with lower rates among immigrants (first-generation) compared to descendants and natives.^41,43^ Country of origin also influenced initiation rates in catch-up programmes, with higher uptake among migrants from non-Western compared to those from Western countries.^41,43^

Mandatory schemes showed mixed impact.^47,78,96^ Support levels for these programmes varied, with some foreign-born parents often showing stronger support for school mandates, compared to other ethnic groups,^96^ while others expressed negative sentiments toward mandatory programmes, particularly when tied to legal residency requirements,^47^ or generating a sense of violation of their rights because of perceived reduced autonomy.^78^

Finally, optional vaccination schemes produced consistently negative results, often due to a perceived lack of priority given to HPV vaccination by both healthcare providers and vaccine recipients.^62,76,80^

### Approaches and strategies to promote uptake

The studies recommended numerous strategies and reported various effective approaches to strengthen HPV vaccination among migrants (**Table 2**).

**Table 2.** Approaches and Strategies to Promote HPV Vaccination Uptake in Migrants.

| Recommendation category | Key approaches recommended by participants and authors | Barriers addressed (BeSD model) |
| --- | --- | --- |
| **Implementing effective health information, education, and communication** | **Appropriate messaging, formats, channels**<br>- Provide culturally and linguistically tailored messages in native language, focusing on strength framing, heightening risk perception or stressing as health promotion, and aligning with parents/ caregivers' values.<br>- Offer clear and balanced information on benefits versus risks (e.g., vaccine benefits versus risks of HPV infection).<br>- Target all stakeholder groups (e.g., beneficiaries, parents/ caregivers, providers).<br>- Use suitable formats for young adults, such as infographics, statistics, narrative videos, and content in native languages on TV or radio.<br>- Use suitable formats for adolescents (e.g., comic books).<br>- Use suitable formats for parents, such as radio programmes, photo/ radionovelas, interactive forums, adverts, and pamphlets.<br>**Leveraging trusted mediators**<br>- Use trusted messengers to reach diverse groups.<br>- Involve peer health promoters as mediators in educational settings.<br>- Engage religious champions, community health workers (CHWs), and key opinion leaders.<br>- Co-design culturally and religiously sensitive messages with CHWs and migrant representatives.<br>**Male-targeted communication interventions**<br>- Involve male celebrities in health messages.<br>- Implement school-based programmes for boys to reduce stigma around HPV vaccination. | - Poor knowledge about HPV/ HPV vaccine.<br>- Negative information or misinformation.<br>- Low confidence in vaccine safety.<br>- Perceived low susceptibility.<br>- Perceived uncertainty.<br>- Cultural and religious.<br>- Gender inequality and mother's lack of autonomy.<br>- Vaccine hesitancy.<br>- Language barrier.<br>- Receiving non-factual information from healthcare providers. |
| **Improving patient/parent-provider or parent-child communication and interaction** | - Encourage providers to actively promote HPV vaccination with appropriate framing (e.g., aligning with parental values, normalising vaccination).<br>- Use strong and clear recommendations.<br>- Build trust and encourage proactive vaccine behaviour.<br>- Provide continuous education for healthcare providers on migrant-sensitive and culturally responsive care.<br>- Better message framing for patient/ parent-provider or parent-child communication (e.g., emphasising as health promotion, heightening risk perception about partner). | - Non-willingness of providers to recommend vaccine.<br>- Receiving non-factual information from healthcare providers.<br>- Low confidence in vaccine safety.<br>- Cultural and religious beliefs.<br>- Poor knowledge about HPV/ HPV vaccine.<br>- Vaccine hesitancy. |
| **Addressing accessibility issues** | **Proximity and free services**<br>- Provide walk-in, mobile clinics, community pharmacies, and transportation to clinics.<br>- Invest in outreach services and on-site vaccination<br>- Publicly funded free HPV vaccination | - Not knowing where to go.<br>- Difficulty/ loss to follow-up.<br>- Cost of vaccines.<br>- Language barrier. |
|  | - Invest in safety-net clinics<br>- Provide high-quality interpretation services, and multilingual providers at service points.<br>**Bundling and piggybacking**<br>- Bundle HPV vaccination with other services or campaigns (e.g., HIV programmes, school curricula, sexual education).<br>- Piggyback on opportunistic vaccination by leveraging all healthcare visits (e.g., GP visits, maternal care, paediatric visits) and existing school-based programmes to provide catch-up. | - Not knowing where to go.<br>- High migrant mobility.<br>- Difficulty/ loss to follow-up. |
| **Strengthening vaccine monitoring** | - Establish national, long-term monitoring systems for vaccine uptake, including data on country of origin, migratory status, and ethnicity.<br>- Implement innovative strategies for mobile groups (e.g., electronic vaccination booklets/ records, eHealth tools).<br>- Implement reminder systems (e.g., texts, calls, letters, and hotlines). | - Lack of vaccination records/ difficulty of follow-up.<br>- High migrant mobility. |

#### Effective health information, education, and communication (IEC)

Information on communication approaches and strategies was abundant, and focused on providing culturally and linguistically tailored messaging in suitable formats and venues, and through trusted messengers, to reach diverse groups effectively and to provide clear information on both advantages and disadvantages (vaccine benefits versus risks of HPV infection and associated cancers).^125,126^

However, it was emphasised that these behavioural change interventions should not be divorced from efforts to improve actual accessibility.^121,145^ Priority was given to multi-level communication strategies targeting all stakeholders, including direct beneficiaries, parents/caregivers, and providers. These included comic books for adolescents, educational forums for mothers,^58,116^ online continuous education courses for providers to enhance responsiveness and sensitivity to the needs of direct beneficiaries and parents.^35^

#### Using appropriate messaging, formats, and channels

It was considered essential to include key messages in vaccine information sheets, such as Halal certification,^82^ and positive vaccination testimonials.^145^ Clear, accessible information about the HPV/HPV vaccine – covering the ‘what, why, and how’ in lay terms – was considered important.^129^ This includes details on prevalence of HPV infection in the host country, modes of transmission, exposure risks, and susceptibility.^118,149^

For young adult migrants, preferred formats included concise information, infographics, statistics;^118,150^ narrative videos, and audiovisual content in native languages broadcast on TV or radio.^65,70^ Other effective channels were educational workshops,^68^ comic books,^58^ and targeted social media platforms like college health centre Facebook pages.^150^ For parents/caregivers, recommended formats included radio programmes (for older parents),^105^ culturally relevant short stories,^36^ photo/radionovelas presented by community health workers (CHWs) in low literacy communities,^125^ and interactive educational forums.^116^ Flyers and pamphlets about the HPV vaccine were also distributed at community venues like clinics and churches.^68^ Hotlines staffed by doctors from the same country, such as Ukrainian doctors in Poland, were also utilised.^80^ Since migrants often use multiple information sources, education focusing on how to access reliable health information and making the most of diverse resources was recommended.^140^

Male-targeted interventions were prioritised to challenge gender-related misconceptions about HPV vaccination. Recommended approaches included featuring male celebrities in health campaigns^36^ and implementing school-based programmes for boys to reduce stigma.^62^

#### Leveraging trusted mediators

The role of trusted mediators is crucial in delivering HPV vaccination messages. Recommended strategies included using peer health promoters in schools to educate students and serve as liaison with healthcare professionals,^111^ and engaging religious champions and CHWs to foster acceptance among caregivers.^36,82^ Emphasis was placed on involving CHWs and migrant representatives not only to deliver messages but also to co-design messages in line with cultural and religious values.

#### Improving patient/parent-provider or parent-child communication and interaction

The role of healthcare providers in promoting HPV vaccination was emphasised, with recommendations for more active involvement in promotion and follow-up. Key strategies included framing vaccination in alignment with parents’ values, normalising the HPV vaccine, providing strong and clear recommendations, building trust, and encouraging proactive behaviour.^76^ Communication preferences varied: some migrant groups preferred in-person interactions,^57^ while others favoured oral and written communication in their native language, either at clinic or sent home.^68,76^

In framing communication, it was recommended to present HPV vaccination as part of general health promotion rather than solely as a measure to prevent STIs.^57,62^ It is also important to highlight that girls can be at risk for HPV through their partners,^36^ a point often under-reported. The focus should be on risk perception, rather than stigmatising promiscuity.^36^

#### Addressing accessibility issues

Although accessibility challenges were major barriers to vaccine uptake, recommendations in this area were limited. Suggested solutions included walk-in centres,^76^ mobile clinics,^68^ school vaccine clinics,^129^ catch-up vaccine days,^129^ community pharmacies,^76,142^ transportation to clinics,^68^ and outreach services.^80,84,104,131^ Free or affordable vaccination,^46,61,62,76,80,103,105,110,112,118,131,135,136,139,142^ and interpretation services at points of care were also recommended.^41,61,68,80,98,147^

Several studies highlighted the effectiveness of bundling and piggybacking approaches. Bundling involved integrating HPV vaccine with other services or campaigns, such as HIV programmes,^118^ sexual education,^60,118^ social and mass media campaigns,^118^ vaccination awareness events on campus,^136^ seasonal flu shot clinic on campus,^148^ and COVID-19 vaccination hubs^145^. Similarly, piggybacking on routine healthcare visits, like GP visits, maternal healthcare, paediatric visits, college health centre visits, or childhood immunisation visits, was recommended to either administer the vaccine or raise awareness.^55,76,84,115,118,147,150^ This strategy ensures dedicated time for discussions between parents and providers, increasing engagement and uptake.^68^

### Strengthening vaccine monitoring

Establishing robust vaccine documentation system was deemed essential. Recommendations included national, long-term monitoring systems to track vaccine uptake, including data on country of origin, migratory status, and race/ethnicity.^76,84,109^ Setting vaccination rate goals for educational institutions was also considered important.^145^ For highly mobile groups (e.g., farmworkers), innovative strategies were suggested, including electronic vaccination booklets/records,^76^ eHealth tools for sharing health data with clinicians in host countries,^68^ and specialised health assessments for refugees upon arrival.^143^ Additionally, reminders via text, phone calls, or postcards were recommended to alert eligible populations and prompt follow-ups for incomplete vaccinations, with lists sent to GPs for tracking.^46^

## Discussion

This systematic review included 933,187 migrant participants from 16 countries and one territory, primarily in HICs. The review identified various barriers, including vaccine safety concerns, cultural beliefs, limited HPV knowledge, gender and family dynamics, negative information, lack of provider recommendations, language barriers, and high vaccine costs. Facilitators included trust in healthcare providers, positive peer experiences, and free, school-based catch-up delivery models. To improve uptake, recommended strategies focus on addressing missed opportunities through bundling and piggybacking approaches, using age-appropriate communication channels, and culturally and linguistically tailored messaging that emphasises positive framing, aligns with parental values, and appeals to their sense of responsibility. Leveraging trusted mediators like peers, CHWs and religious leaders, to deliver and co-design messages, alongside effective provider communication, was also emphasised. Addressing physical barriers through outreach services (mobile clinics) and strengthening vaccination data systems, especially for mobile groups like migrant workers, were key recommendations.

In this study, individual-level knowledge, perceptions, beliefs, and social norms surrounding the vaccine among parents/caregivers and recipients were recurring factors, reflected in multiple recommendations focusing on effective IEC on health. This is consistent with previous literature reviews in non-migrant groups which shows low levels of knowledge about HPV/HPV vaccination among global indigenous communities,^151^ rural populations in the USA,^152^ non-immigrant parents of female adolescents in ASEAN countries,^153^ and ethnic minority adolescent girls.^154^ This highlights the importance of education in improving knowledge and shaping attitudes for migrants and vulnerable host populations alike. A prior systematic review of 2,206 immigrant parents’ perceptions of HPV vaccination found low levels of awareness and negative perceptions, which often improved with information.^155^ In this review, we found that knowledge- and perception-related barriers may be offset by receiving accurate and effective communication through trusted messengers and appropriate channels, including doctors from same background recommending the vaccine or peers sharing positive experiences with vaccination services. However, we also found that healthcare providers might hesitate to recommend the vaccine if it conflicts with the cultural beliefs of migrants. Provider recommendations play an important role in HPV vaccine uptake,^152^ and their absence negatively affects uptake, even when the vaccine is provided for free. A global meta-analysis on the impact of provider communication on HPV vaccination among 265,083 patients in the USA has shown that provider recommendations substantially increased HPV vaccine initiation compared to no recommendation (24% versus 60%, pooled OR=10.1, 95% CI: 7.6–13.4), as well as vaccine completion.^156^ Discussions with healthcare providers were also similarly associated with higher HPV vaccine initiation (OR=12.4, 95% CI: 6.3–24.3).^156^ This further corroborates our findings, emphasising the need for active provider involvement in promoting vaccination.

Accessibility issues related to the vaccine were a major factor affecting uptake in this review. While global HPV coverage remains significantly low, HICs have achieved better rates, though still falling short of WHO’s 90% target.^157^ For example, first-dose HPV coverage in 2023 was below this target in Europe, the Americas, and the Western Pacific (62%, 57%, and 70%, respectively).^157^ Vaccination rates among migrants and refugees are similarly low, with a recent systematic review suggesting completion rates of 63.4% (48.0–78.8%) in Europe, 6.0% (3.9–8.2%) in the Americas, and 7.8% (7.09–8.52%) in the Western Pacific Region.^22^ Notably, 95% of vaccinated populations—both migrants and non-migrants—against HPV globally are in HICs,^22^ suggesting migrants in LMICs face greater challenges, which remain under-explored. There is an urgent need for affordable, locally manufactured HPV vaccines. A recent analysis of HPV programmes in 18 Asian LMICs identified major implementation challenges, including vaccine shortages, lack of subsidies, and reliance on out-of-pocket payments, contributing to low affordability.^158^ They also reported a lack of national surveillance data on HPV vaccination.^158^ These issues impact both local populations and migrants, with the latter facing additional barriers due to lower healthcare utilisation and high mobility, especially in groups like migrant farmworkers. Fortunately, the WHO’s single-dose HPV vaccine guidelines^159^ present an opportunity to streamline vaccination delivery, particularly for mobile groups like migrants.

We found school-based programmes were reported to achieve consistently superior results in promoting HPV vaccination initiation and uptake among migrants in HICs. School-based programmes are considered gold-standard models in HICs that have achieved high vaccination coverage (e.g., Sweden and Australia).^160^ These programmes have also proven successful in LMICs, outperforming routine and facility-based immunisation approaches.^161^ However, their applicability to migrants in non-HICs, especially newly arrived or undocumented migrants, may be limited. Therefore, it is crucial to ensure that out-of-school girls, a common group among migrant populations in LMICs, are also reached. Some LMICs have implemented hybrid models combining school-based, health centre, and campaign-based delivery to reach out-of-school girls,^161^ and their potential for reaching migrant groups warrants further investigation.

The evidence base on HPV vaccination drivers and strategies has significant gaps that need future research. There is an alarming lack of studies in LMICs, especially low-income countries with limited vaccine availability. Regions like the Middle East and North Africa (MENA), which hosts half the world’s migrant workers and countries which have the highest refugee populations *per capita*, are also underrepresented. There is a scarcity of studies on undocumented migrants, with unique needs and often exhibit more reluctance to engage with mainstream vaccination services or existing services may not be adapted for them. There is also limited data on the role of fathers in HPV vaccine decision-making and the impact of community engagement on improving uptake and acceptance. Behavioural and social factors influencing vaccination uptake are not systematically collected, and current methods lack standardisation, making cross-study comparison difficult. Global and regional efforts are needed to standardise data collection as part of routine reporting. This is particularly important for migrants, where vaccine entitlement does not guarantee uptake. Moreover, logistical challenges such as vaccine supply chains, funding, and political support need attention, as well as tools to navigate vaccine access for newcomers. Exploring the sharing of electronic health records including digital vaccine histories and key information (contacts and country of origin, race/ethnicity, migratory status for mobile migrants) and harmonising relevant regional and global legislation and infrastructure could improve service delivery. It is also important to consider integrating national statistics on migratory status into vaccination registries for better global and regional monitoring. Going beyond educational interventions alone or a one-level approach has also been called for elsewhere.^162^ Furthermore, studies exploring interventions and their implementation are scarce, and the actual empirical impact of interventions remains under-documented. Future implementation research should explore successful models from other vaccination programmes, such as co-design approaches,^163^ COVID-19 delivery models,^164^ and equitable vaccination strategies,^165^ to adapt them for HPV vaccination. Additionally, given the current evidence on barriers and facilitators to screening in migrants,^166^ addressing targeted screening and vaccination programmes for migrants should be prioritised.

The strengths of this review include providing a comprehensive understanding of the factors influencing HPV vaccination uptake among migrants, using an established framework and highlighting successful strategies informed by stakeholder recommendations. It also addresses less prominent and inconclusive factors often overlooked in the literature, illustrating the complexity of uptake drivers and their overlap with socio-demographic and economic factors. We examined programme design and delivery modes, showing how venue, timing, and legal status can affect uptake. However, several limitations should be considered. First, although inherent to migrant health research, the definition of ‘migrant’ varied across studies, complicating meta-analyses or subgroup analyses. Second, there were no studies on sexual minority migrants or migrant sex workers, which would have been relevant. Third, family reunification data was rarely found in our review, which is critical as registry-based studies may not contain such data along with other types of residence permit (refugee, labour migrant, student).^43^ Data on visa transitions can also be incomplete.^51^ Fourth, trust in health systems and its link to vaccine hesitancy may be underestimated, as only nine studies were conducted post-2020, when COVID-19 impacted perceptions. Finally, the broad scope of the review, encompassing diverse migrant groups, limits the depth of insights for specific populations, resulting in a broad overview of best practices rather than targeted policy guidance.

In conclusion, this review highlights that, despite global commitments to equitable vaccination across the life-course, persistent social, behavioural, and systemic barriers continue to block HPV vaccine uptake among migrants. While we have identified multiple studies globally, migrants in LMICs are significantly underrepresented, putting them at risk of being left behind in the fight against preventable cervical cancers and progress toward cervical cancer elimination. It is essential to prioritise these populations in research, identify key drivers of vaccine uptake, and collaborate with migrant communities to create tailored, effective delivery models that meet their specific needs.

## Contributors

SH and MI generated the protocol with input from MSR and RM. MI did the database search, MI, MSR, and RM did title/abstract screening and full-text review. MI and MSR did data extraction. MI, RM, and OB did risk of bias assessments. MI did the data analysis with input from OB and SH. MI and OB wrote the first draft of the manuscript with input from SH and all other authors. SH supervised the work. SH and MI accessed and verified the data.

## Data Sharing Statement

Data are available on reasonable request from the corresponding author.

## Data Availability

All data produced in the present study are available upon reasonable request to the authors

## Acknowledgments

This work was funded by NIHR Advanced Fellowship (NIHR300072), the Academy of Medical Sciences (SBF005\1111), and the Medical Research Council (MRC/N013638/1). SH is also funded by the National Institute for Health and Care Research (NIHR300072 and NIHR134801), MRC (MRC/N013638/1), Wellcome Trust (318501/Z/24/Z), La Caixa Foundation (LCF/PR/SP21/52930003), and WHO. AD is funded by the Medical Research Council (MRC/N013638/1). OB is funded by La Caixa Foundation (LCF/PR/SP21/52930003), MSR is a Clinical Lecturer in primary care funded by NIHR. HMS, SK, NMA & YA are grateful for funding from the St George’s Hospital Charity.

---

**Table S1.** Inclusion and exclusion criteria.

| Inclusion criteria | Exclusion criteria |
| --- | --- |
| Published from 1 January 2006 onwards; any language; any geographical regions/ areas.<br>Community (including a humanitarian setting) and/or clinical settings.<br>Adolescent and adult migrants (defined as foreign-born) and children of migrant parents who are eligible for HPV vaccination programme irrespective of gender, sexuality, migratory status, and socioeconomic status.<br>Studies on other stakeholders' views/perceptions about barriers/ facilitators to uptake (/service delivery) of HPV vaccines in (/for) migrants.<br>Qualitative studies, observational studies (e.g. cohort/ cross-sectional/ case-control/ longitudinal studies; case studies; natural experiments, quasi experiments), randomised controlled trials (RCTs), and before-after studies, which reported primary data on negatively or positively influencing factors to the uptake (/service delivery) of HPV vaccines among migrants globally.<br>Secondary data analysis, as long as meeting the above criteria.<br>Studies that migrants were analysed as sub-group, as long as meeting the above criteria.<br>Studies that HPV vaccination were included in wider vaccination/ vaccine studies, as long as meeting the above criteria.<br>Grey literature, including theses, dissertation, non-peer-reviewed literature. | Internal migrants.<br>Migrant clinicians.<br>Animals (e.g. migrant animals are excluded).<br>Editorials, opinion pieces, commentaries, protocols), policy documents, and guidelines; conference abstracts/ posters/ proceedings, case (series) reports, systematic/scoping/rapid/umbrella reviews.<br>Economic studies.<br>Not transparently report on migrants or migratory status.<br>Out of scope - Reports that were thoroughly about other vaccines (e.g. flu, MMR) but did not include HPV vaccines, or not about HPV vaccines for prophylactic use but only for therapeutic use.<br>Out of scope - Focusing on factors that influenced uptake of cervical/ anal/ oral cancer screening, HPV testing, self-sampling, HPV infection rates, ethics of HPV vaccines etc, but not thoroughly about factors that influenced uptake (/service delivery) of HPV vaccination.<br>Out of scope - Reporting on vaccine design/ development/ production, or tool development (including survey tools).<br>Out of scope - Reporting on lab studies, sero-surveillance, molecular virology, immunology, biological assays, genome studies.<br>Not available on internet, beyond institutional access, or full-text is not retrievable.<br>Duplicate. |

**Table S2.**
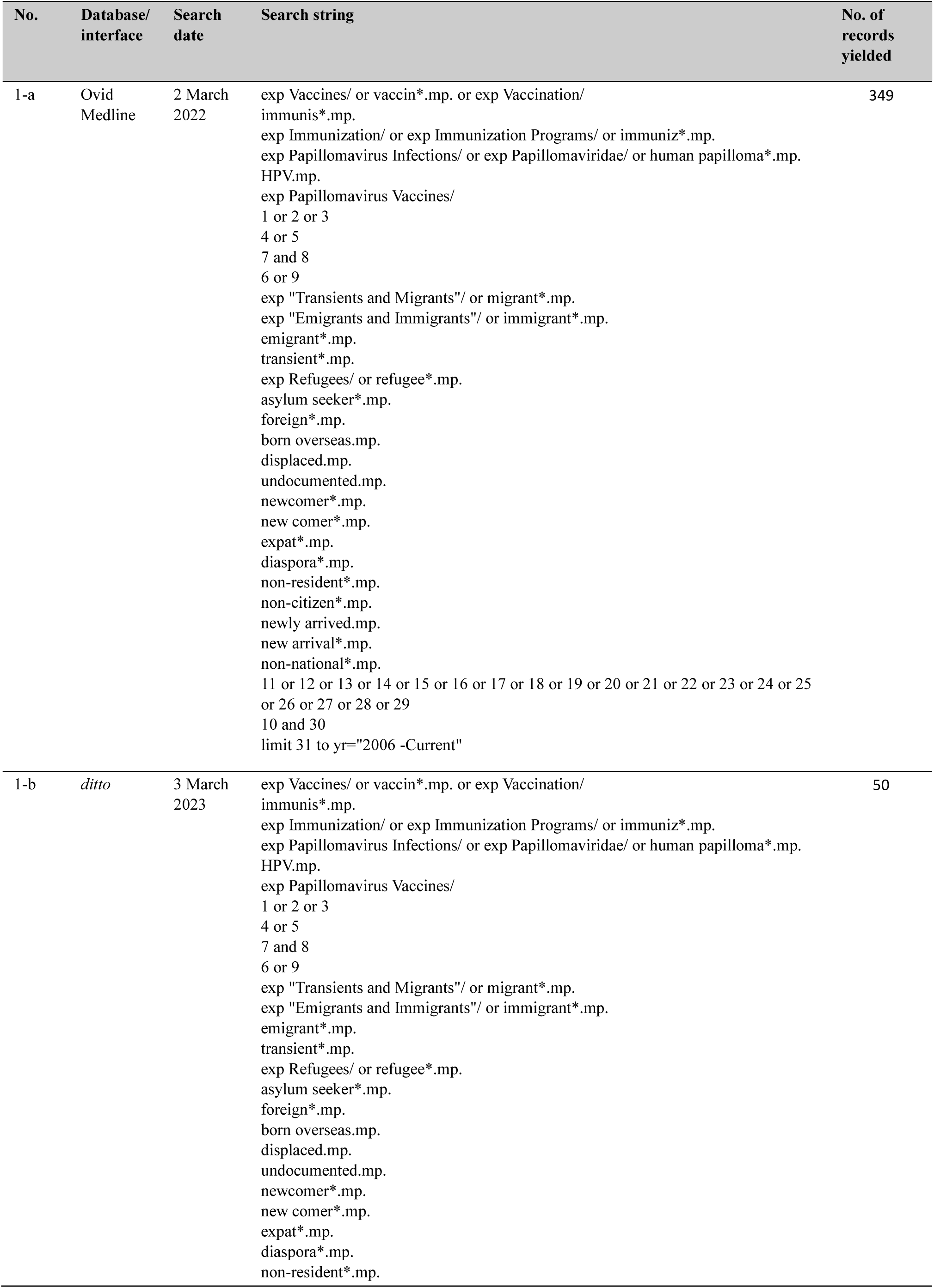
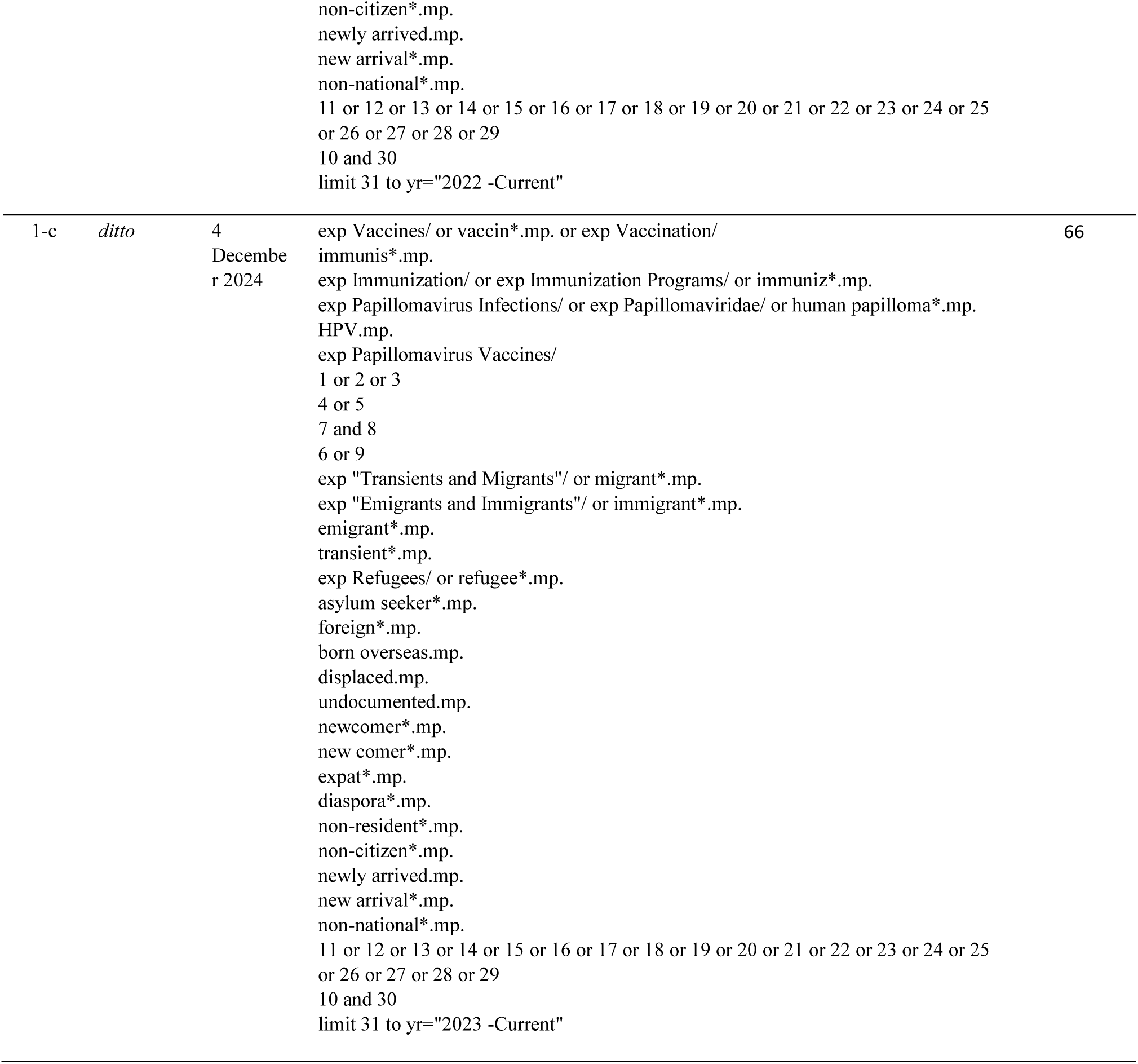
Search strategy MEDLINE (Ovid) (January 2006 – December 2024)

**Table S3.**
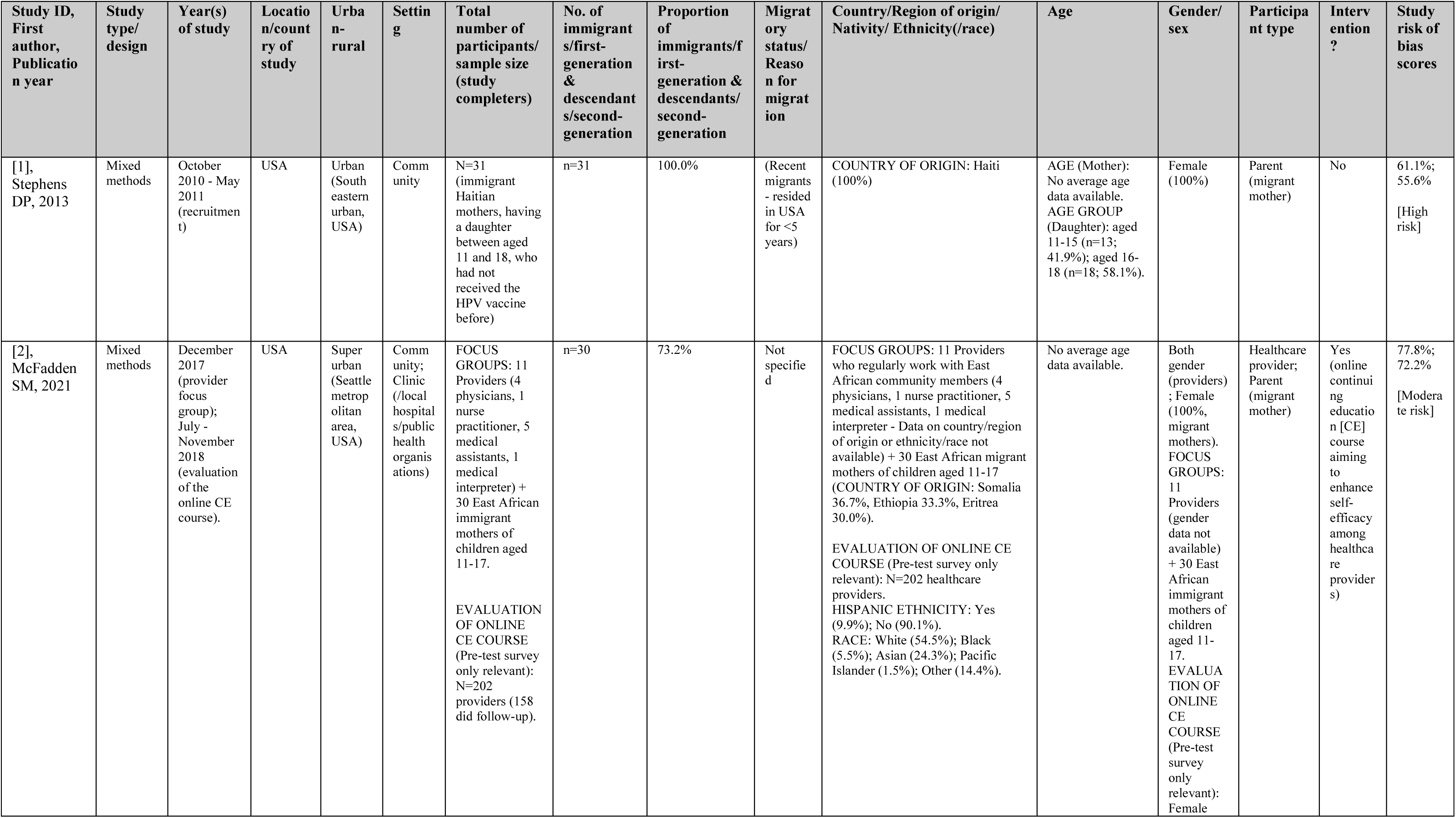
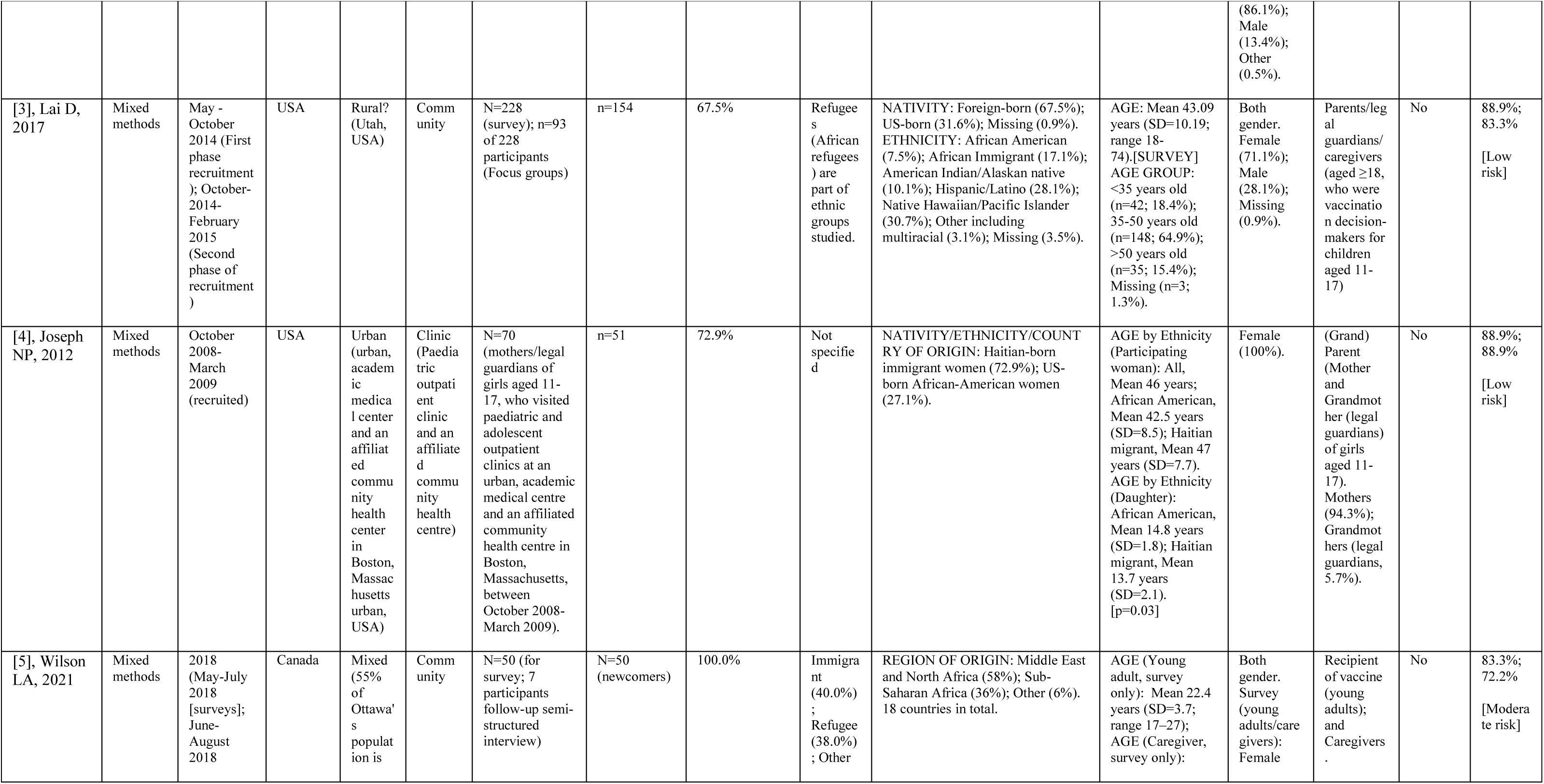
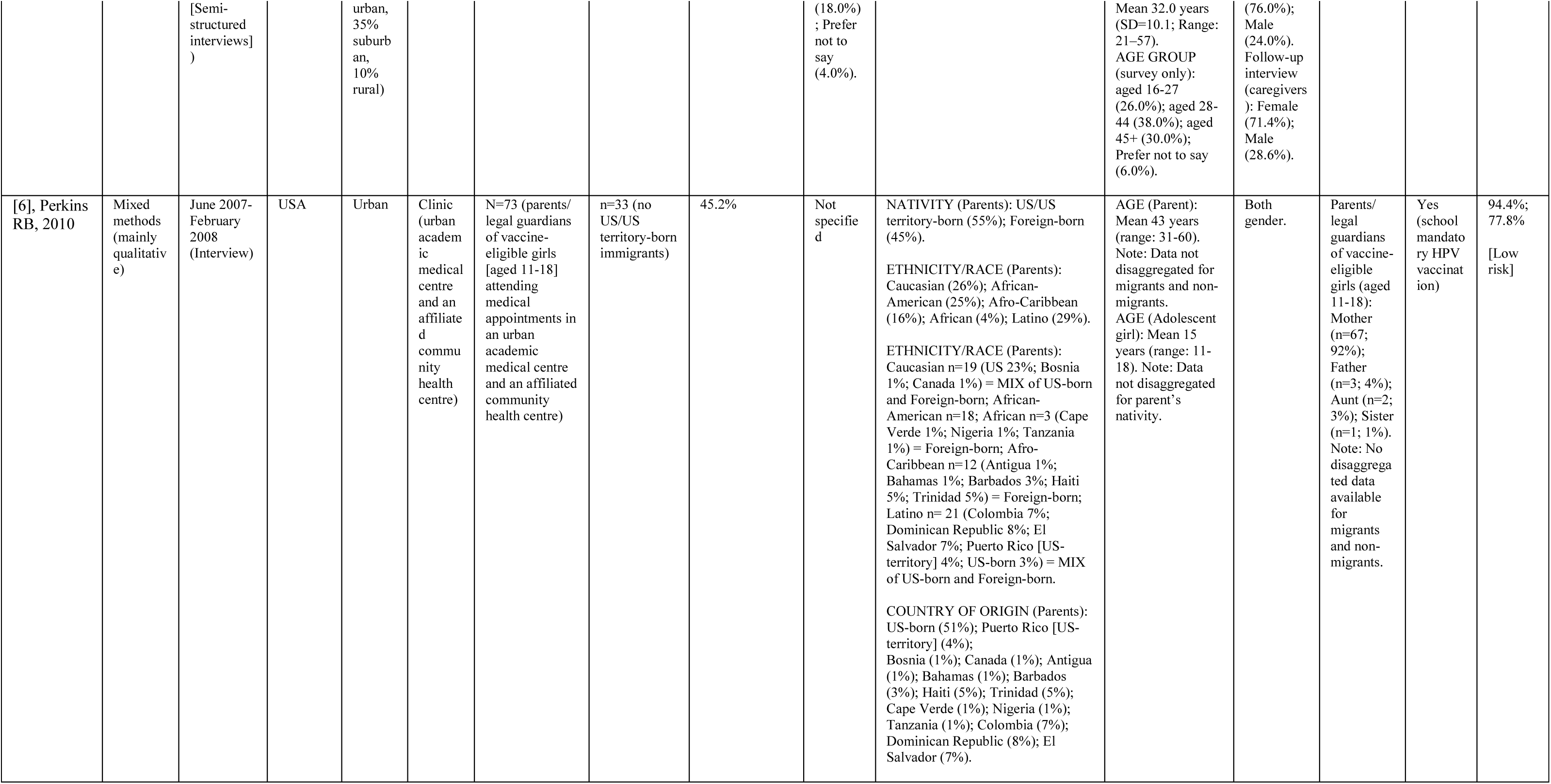
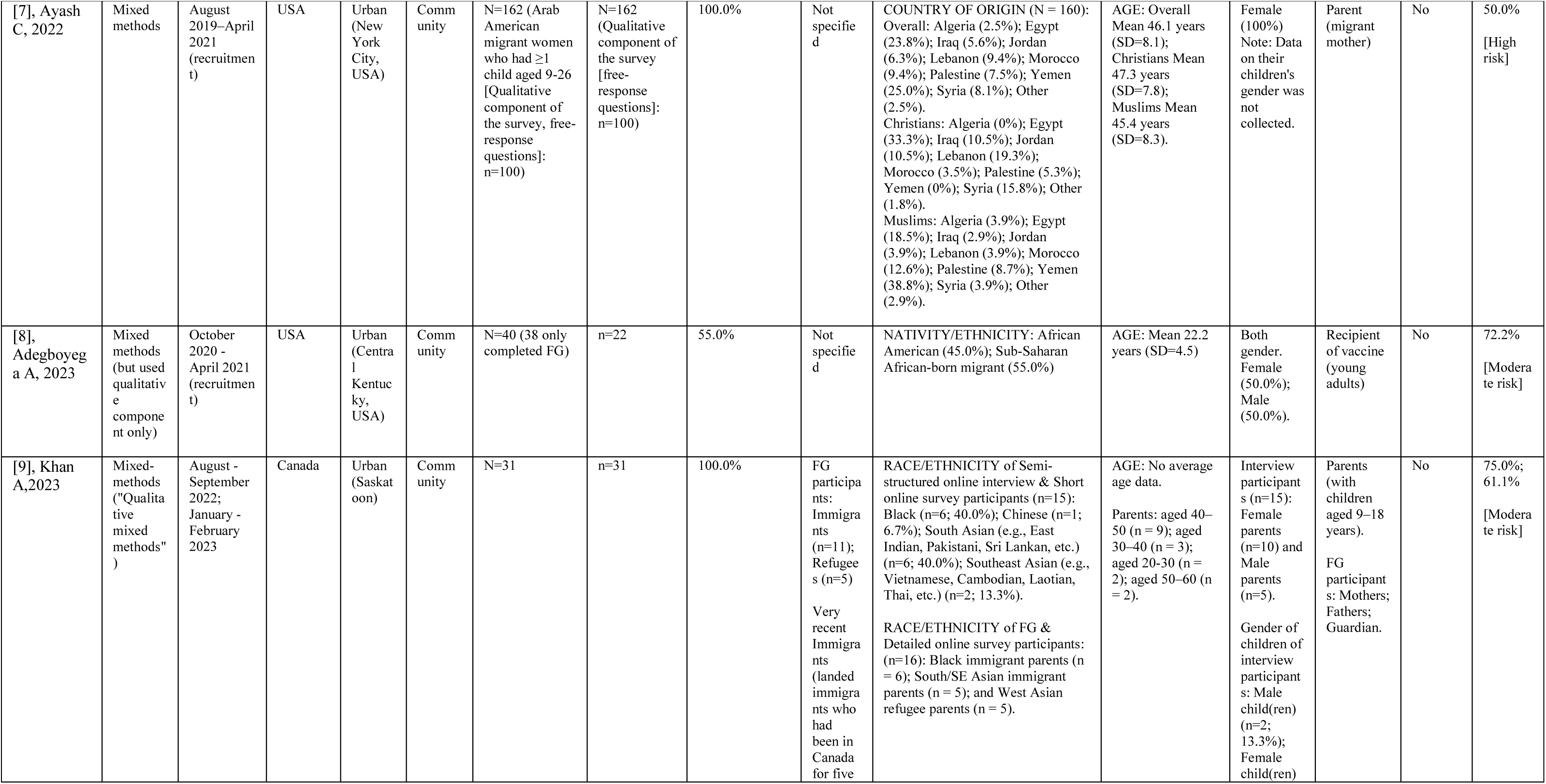
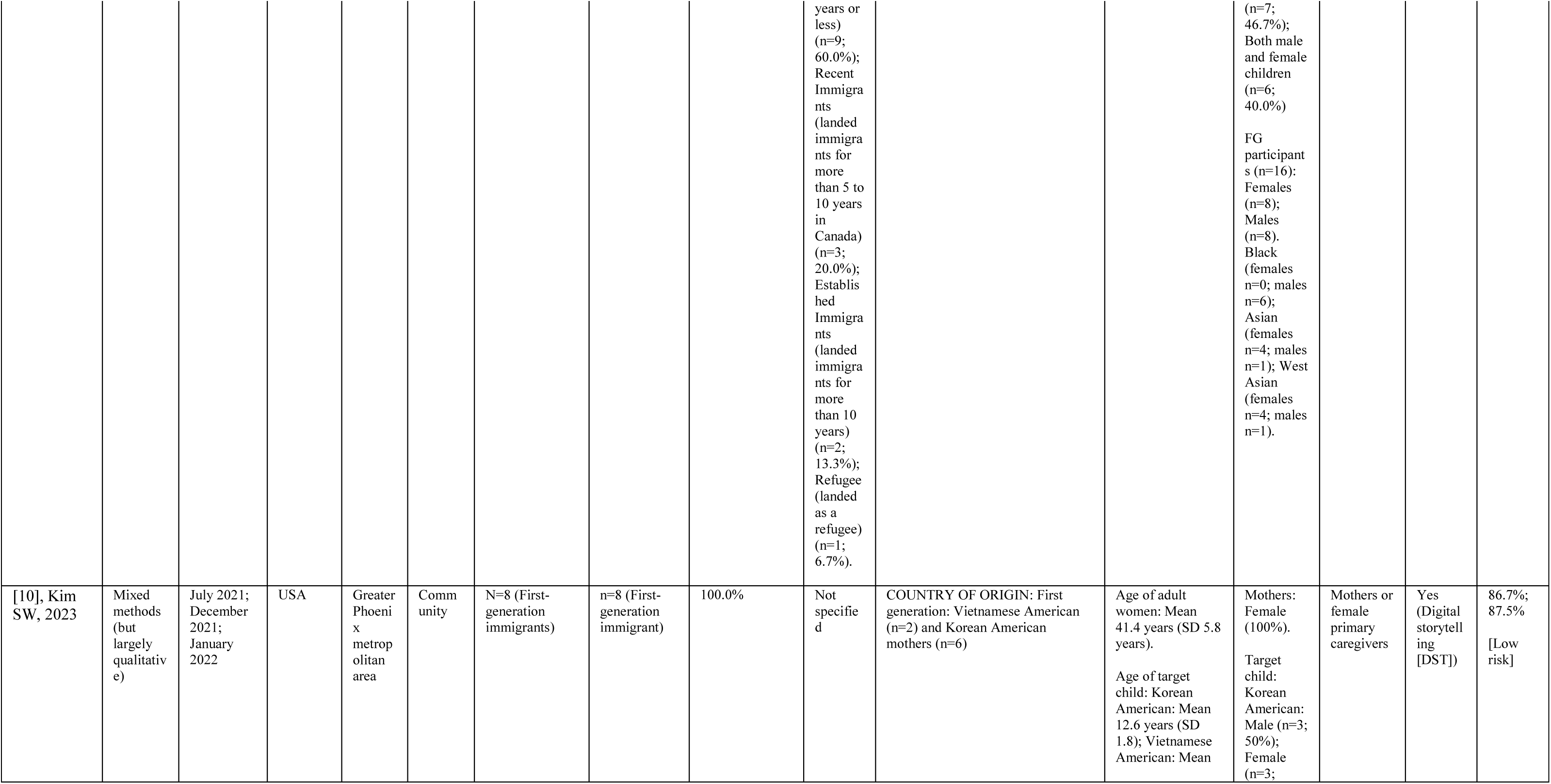
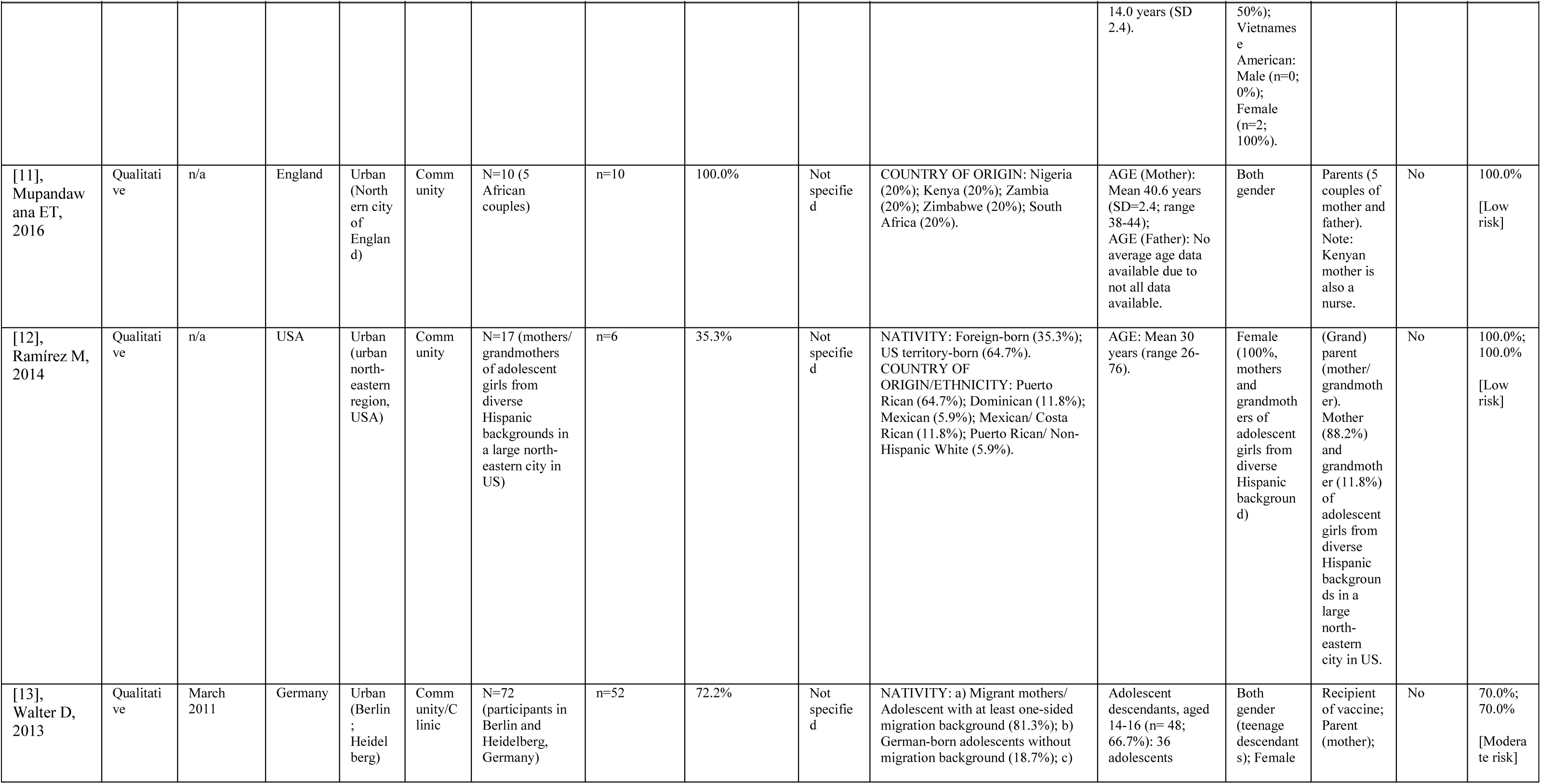
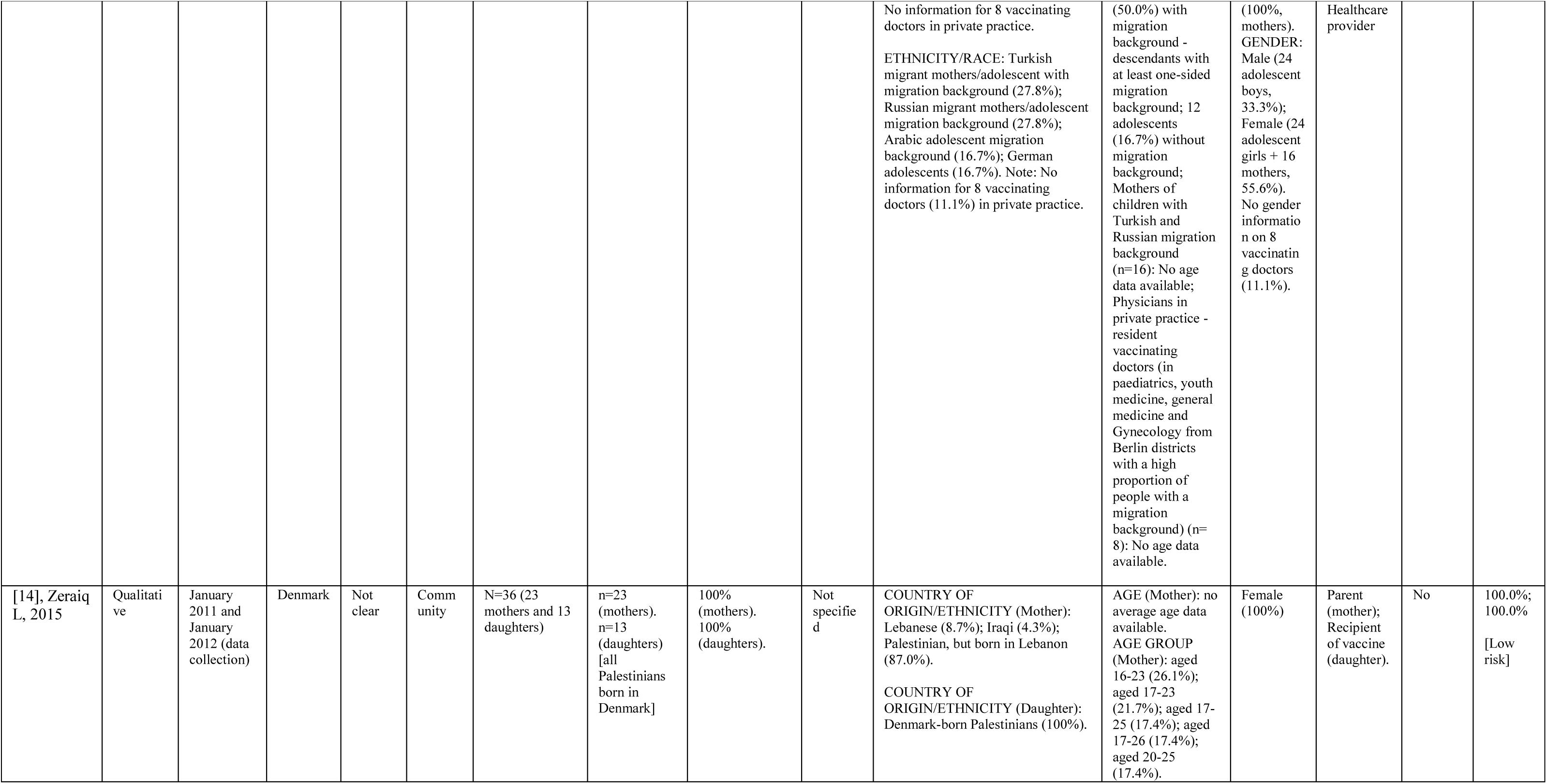
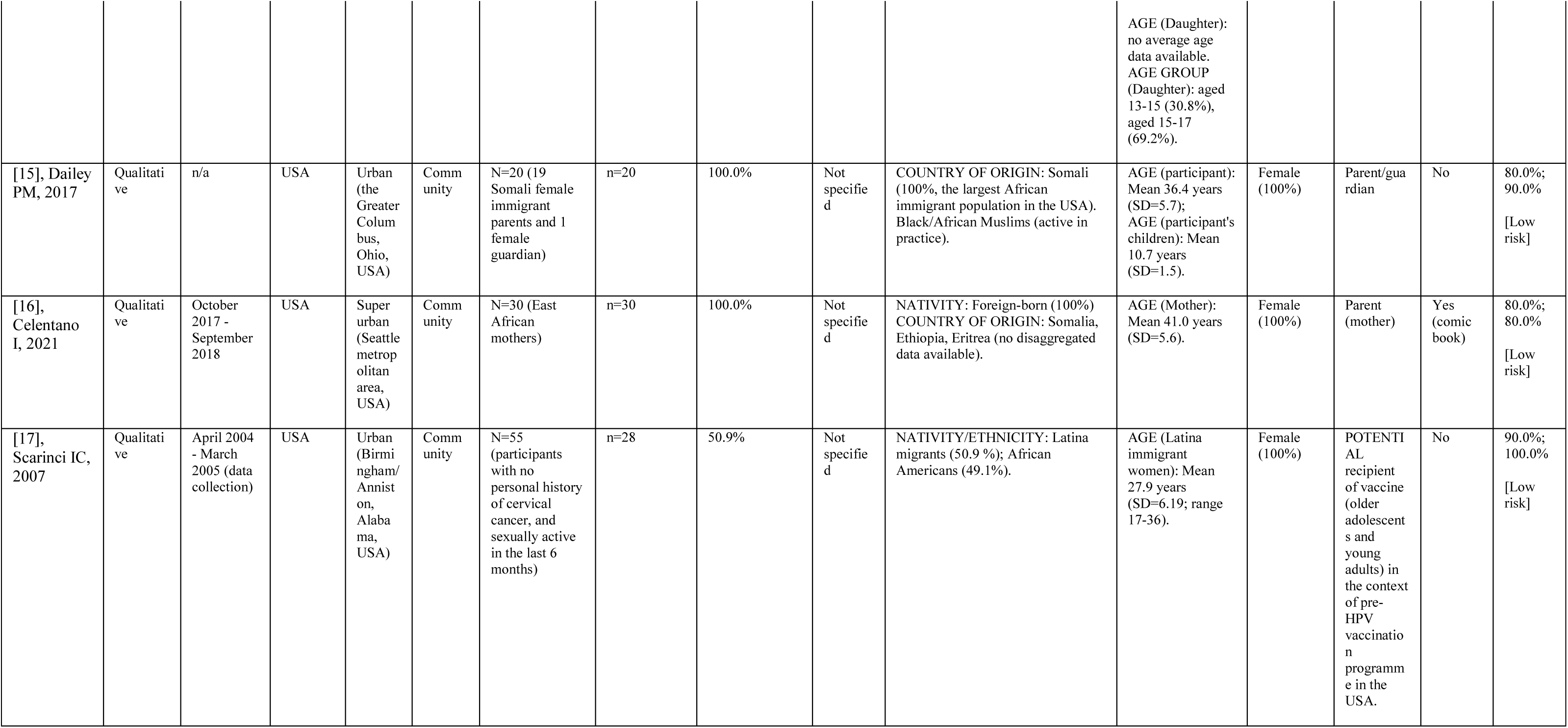
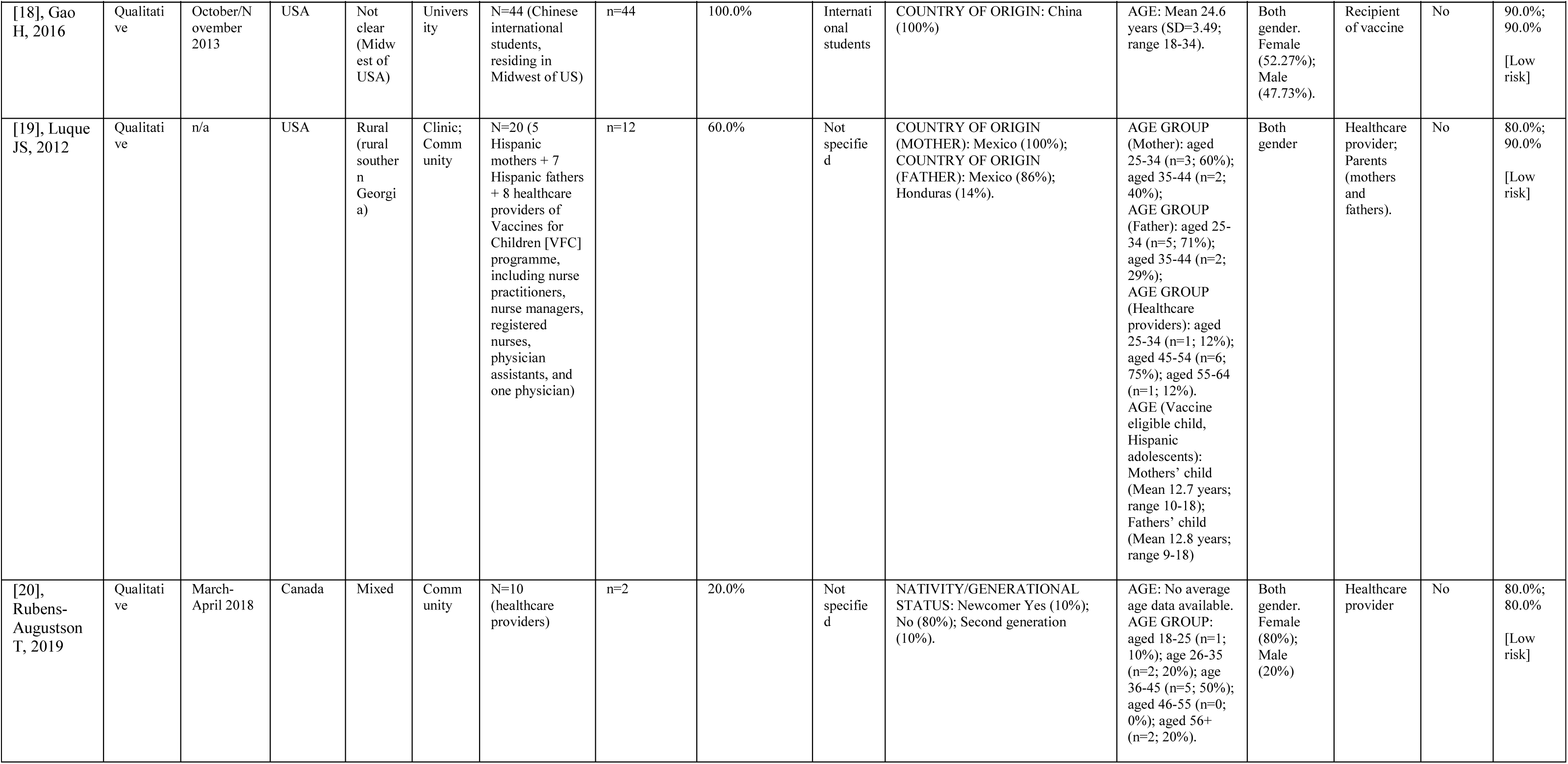
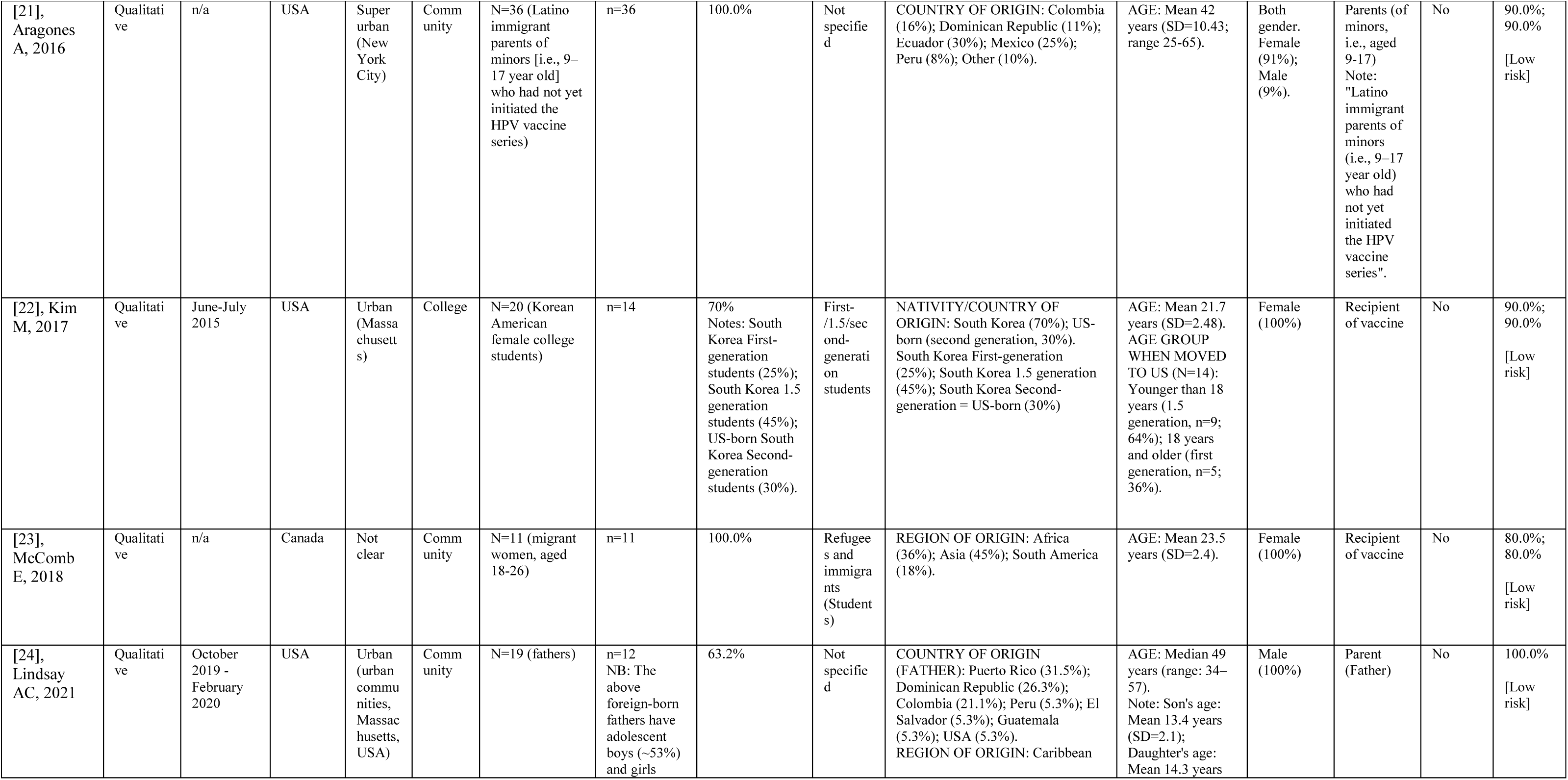
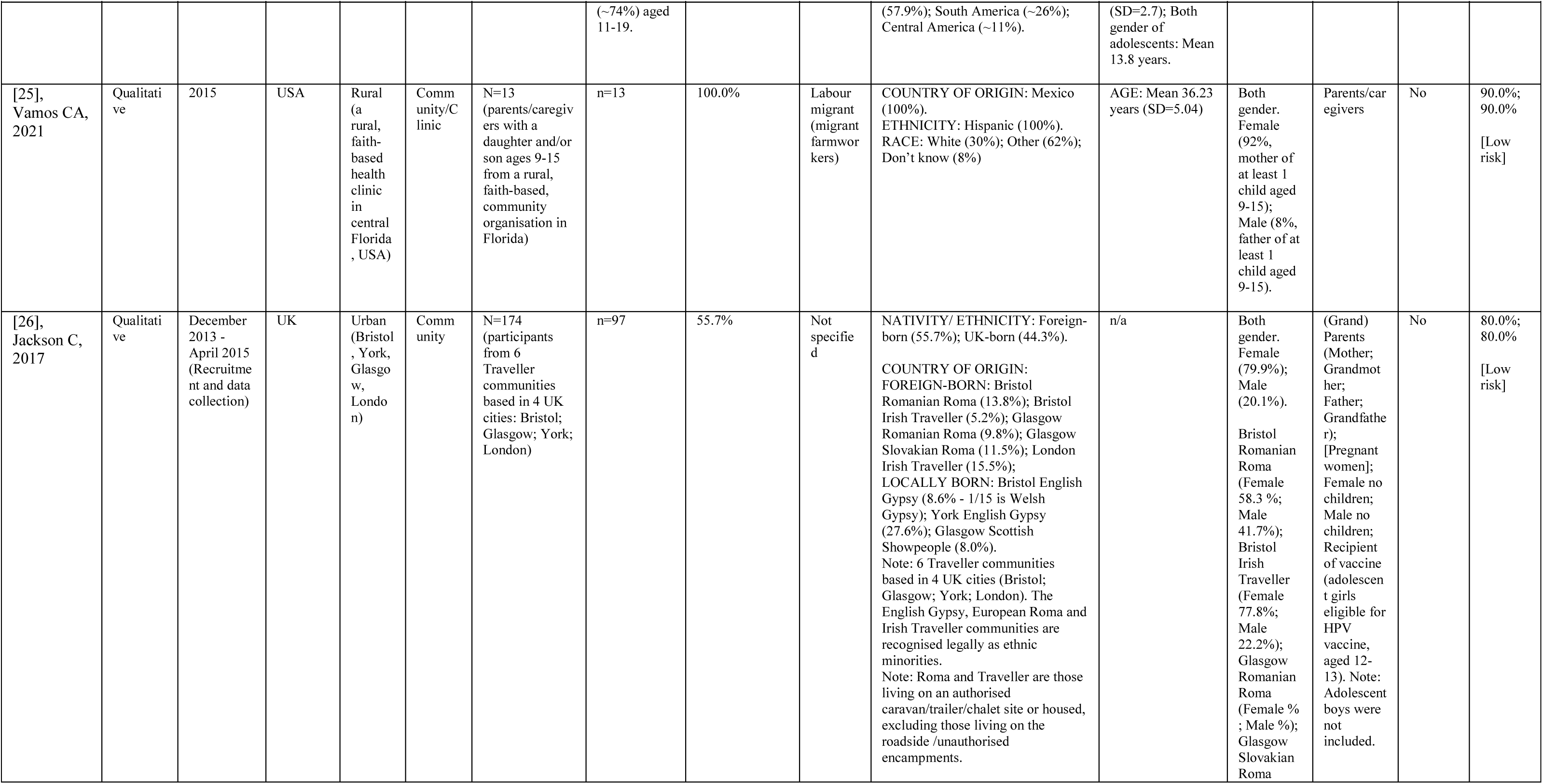
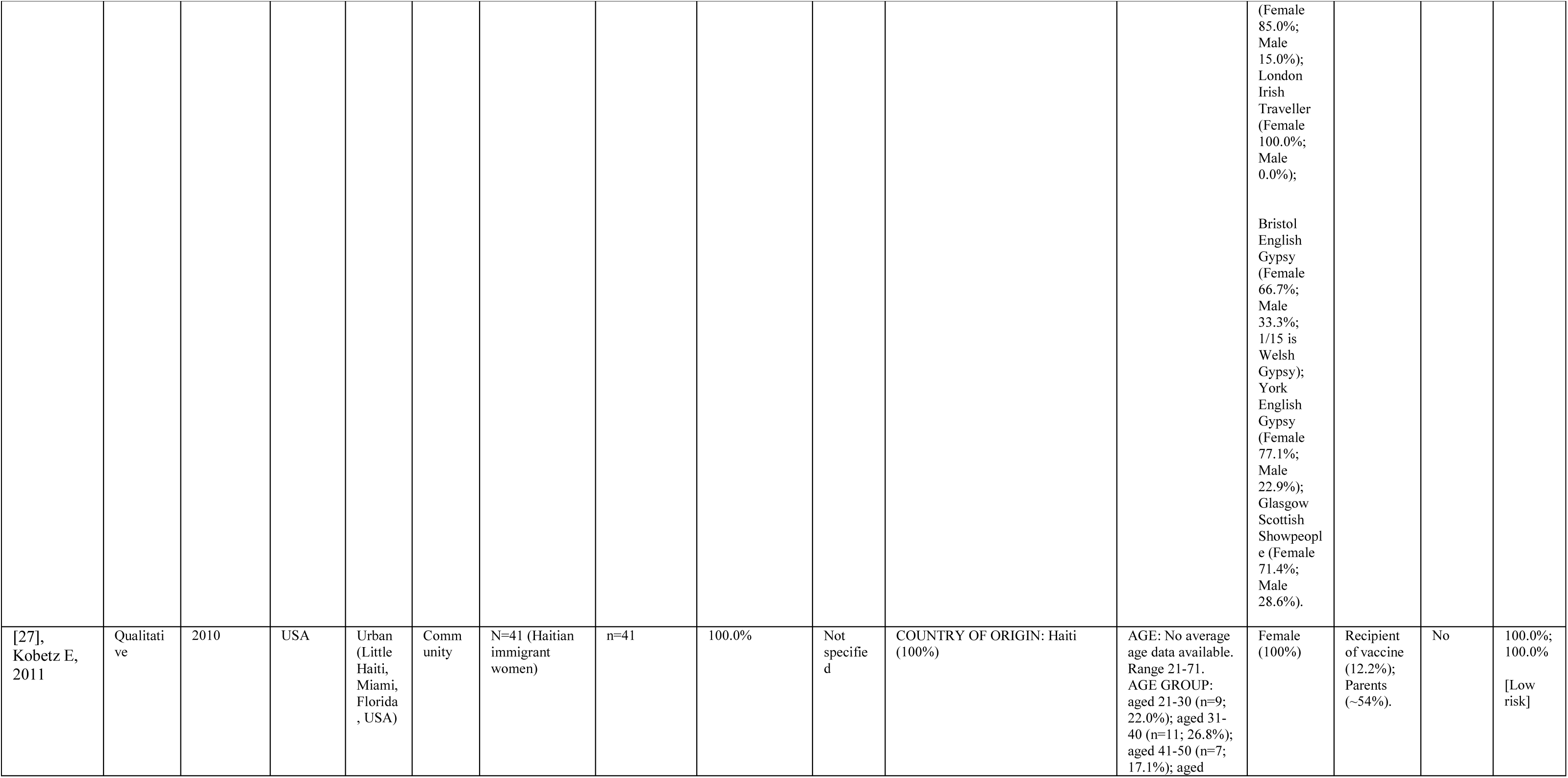
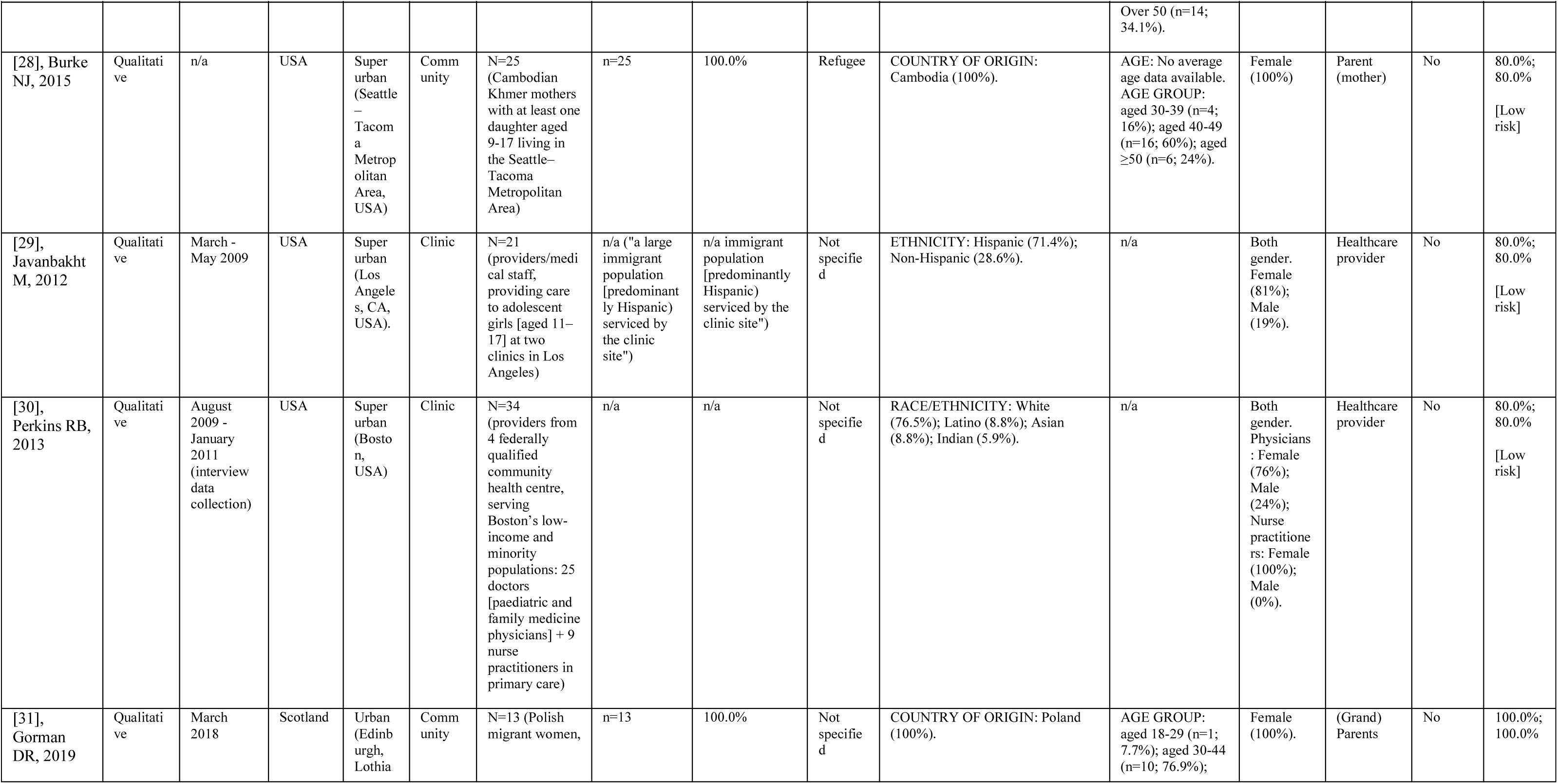
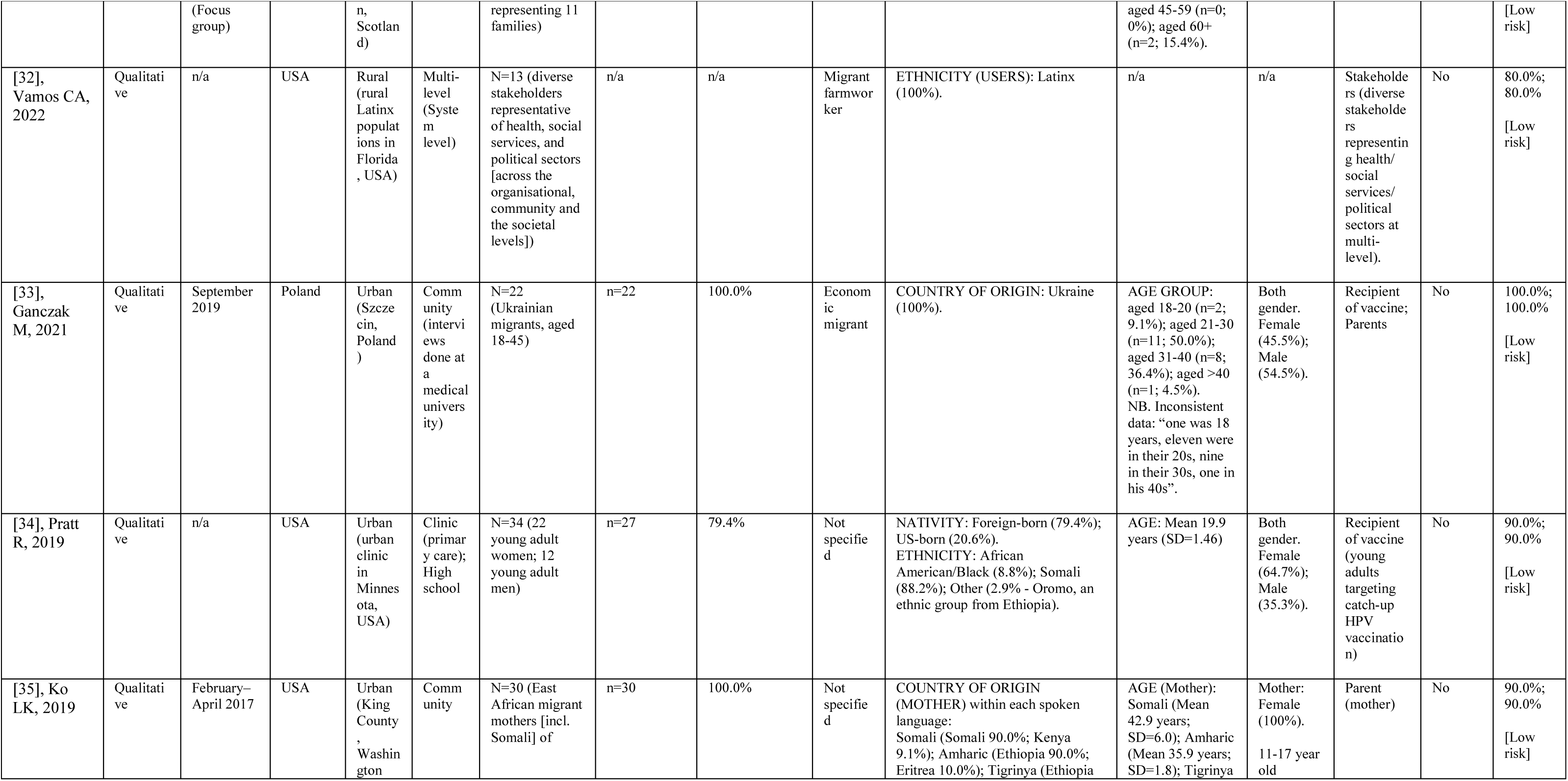
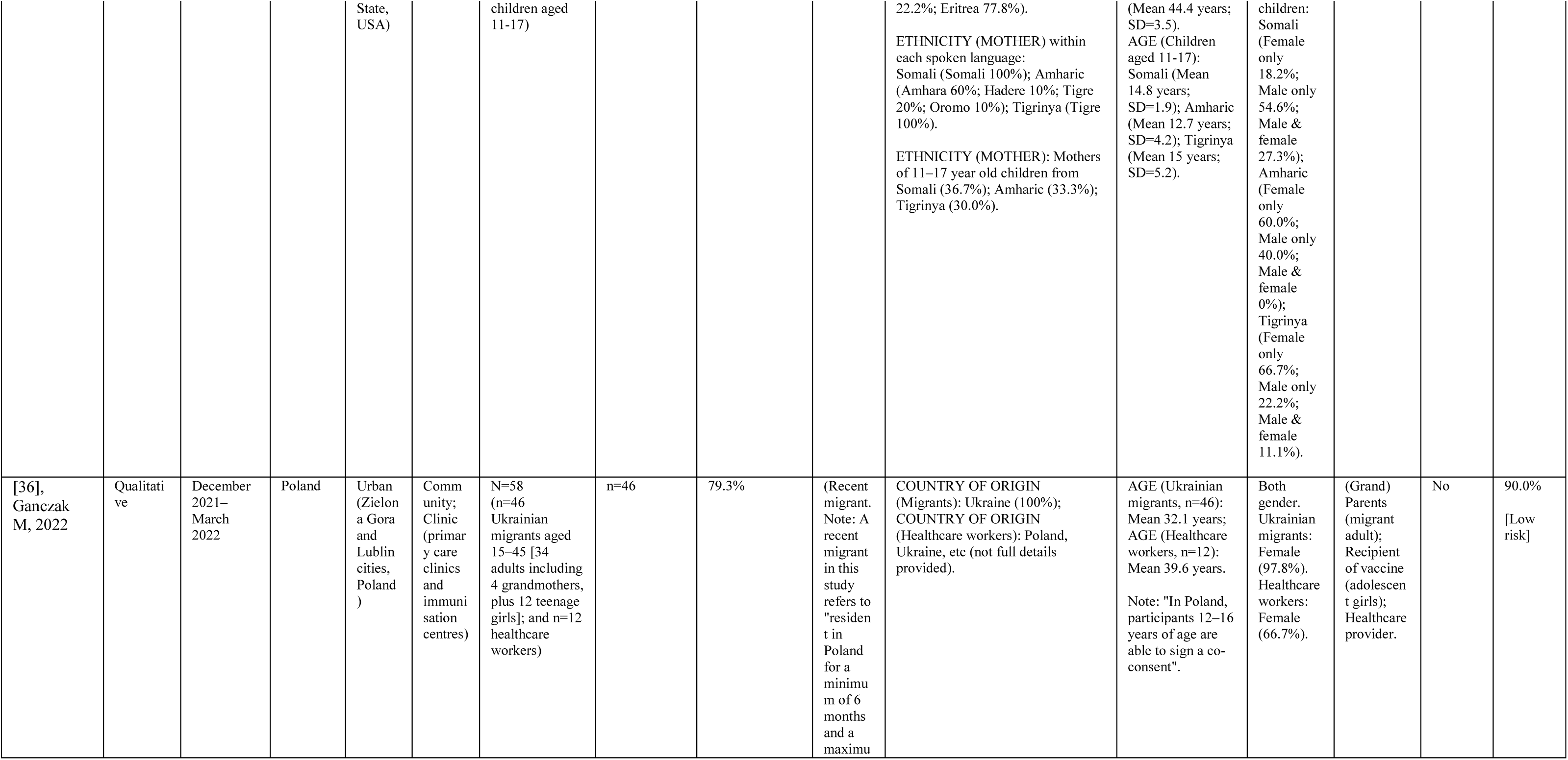
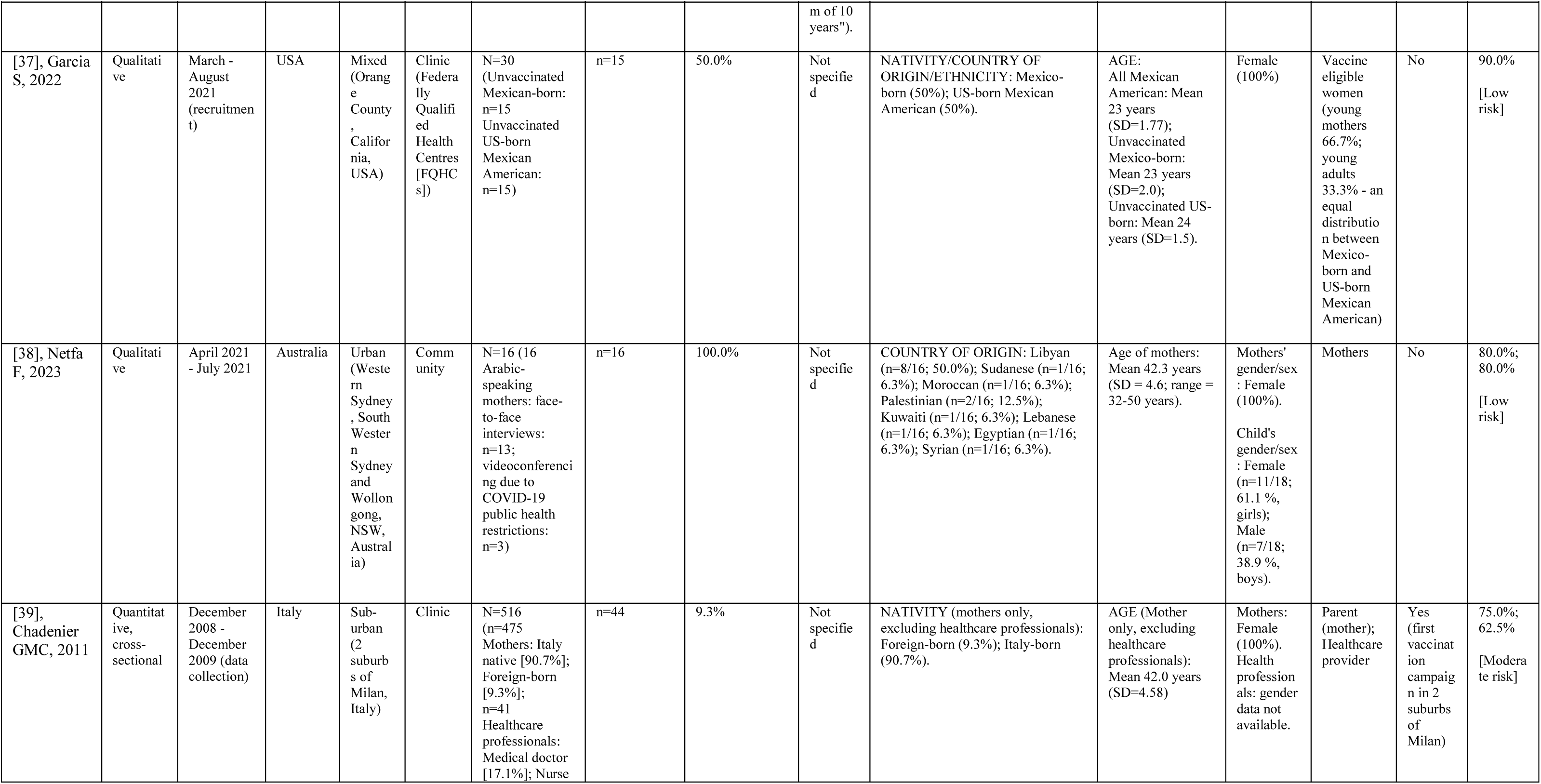
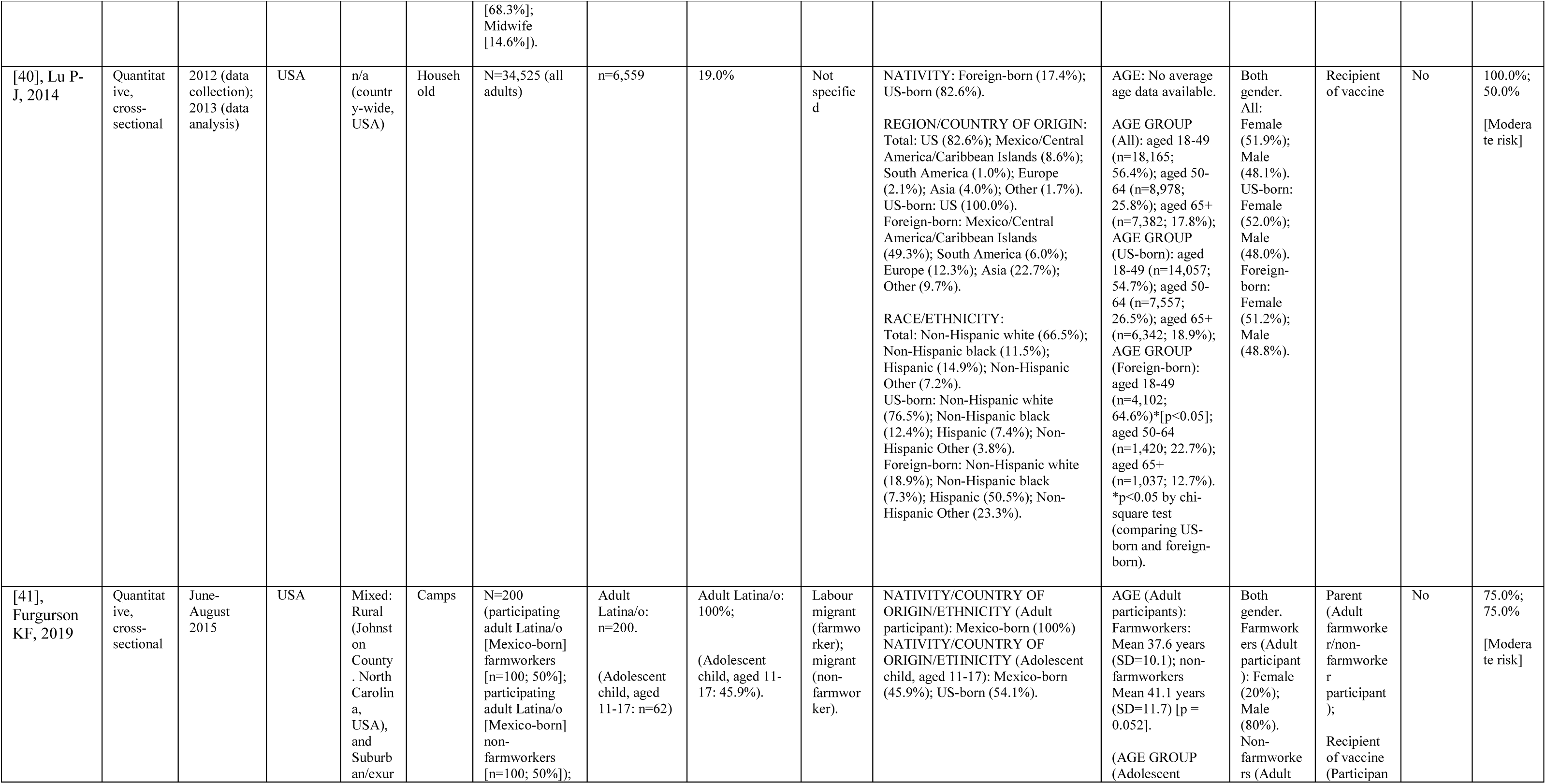
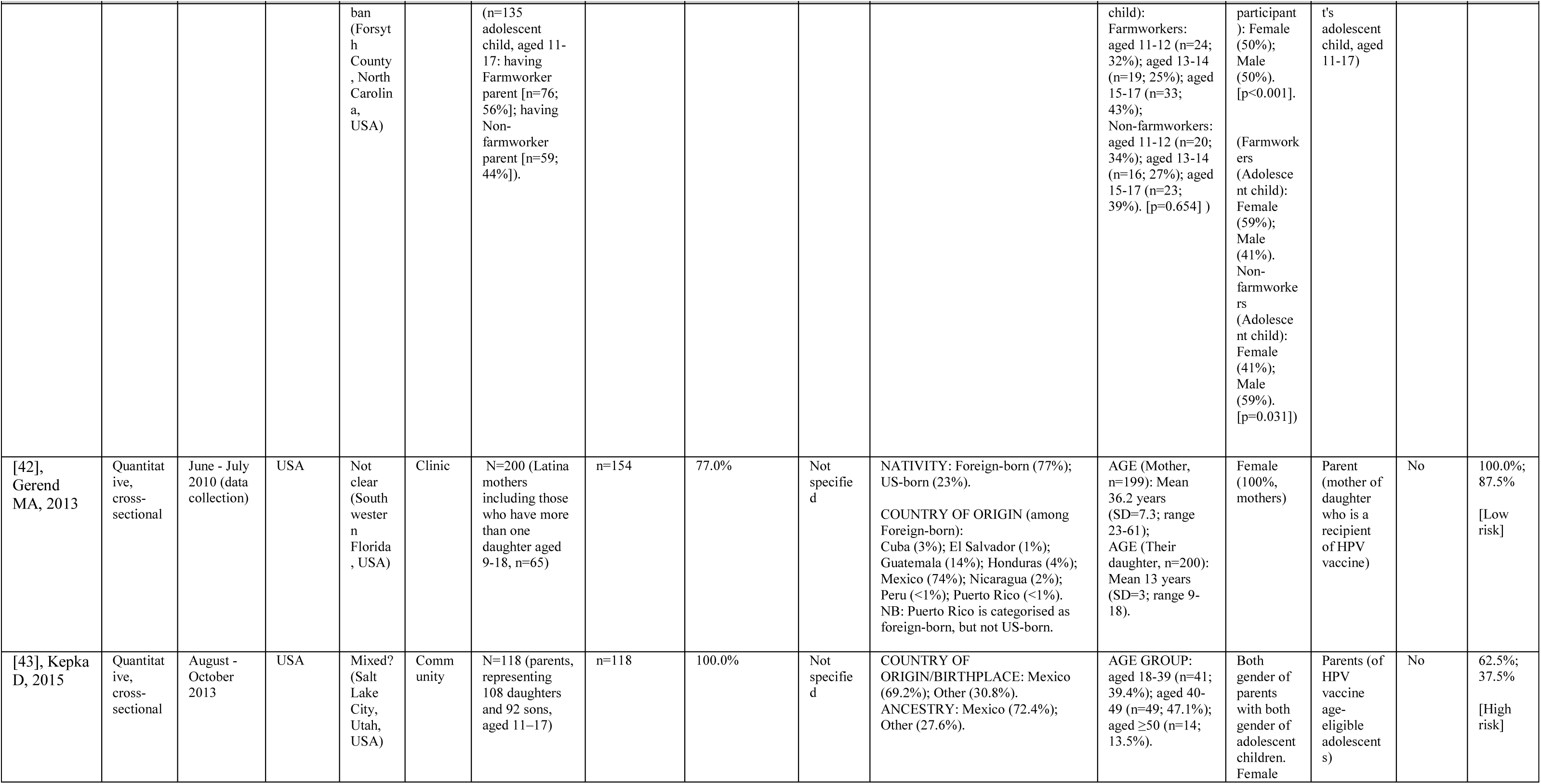
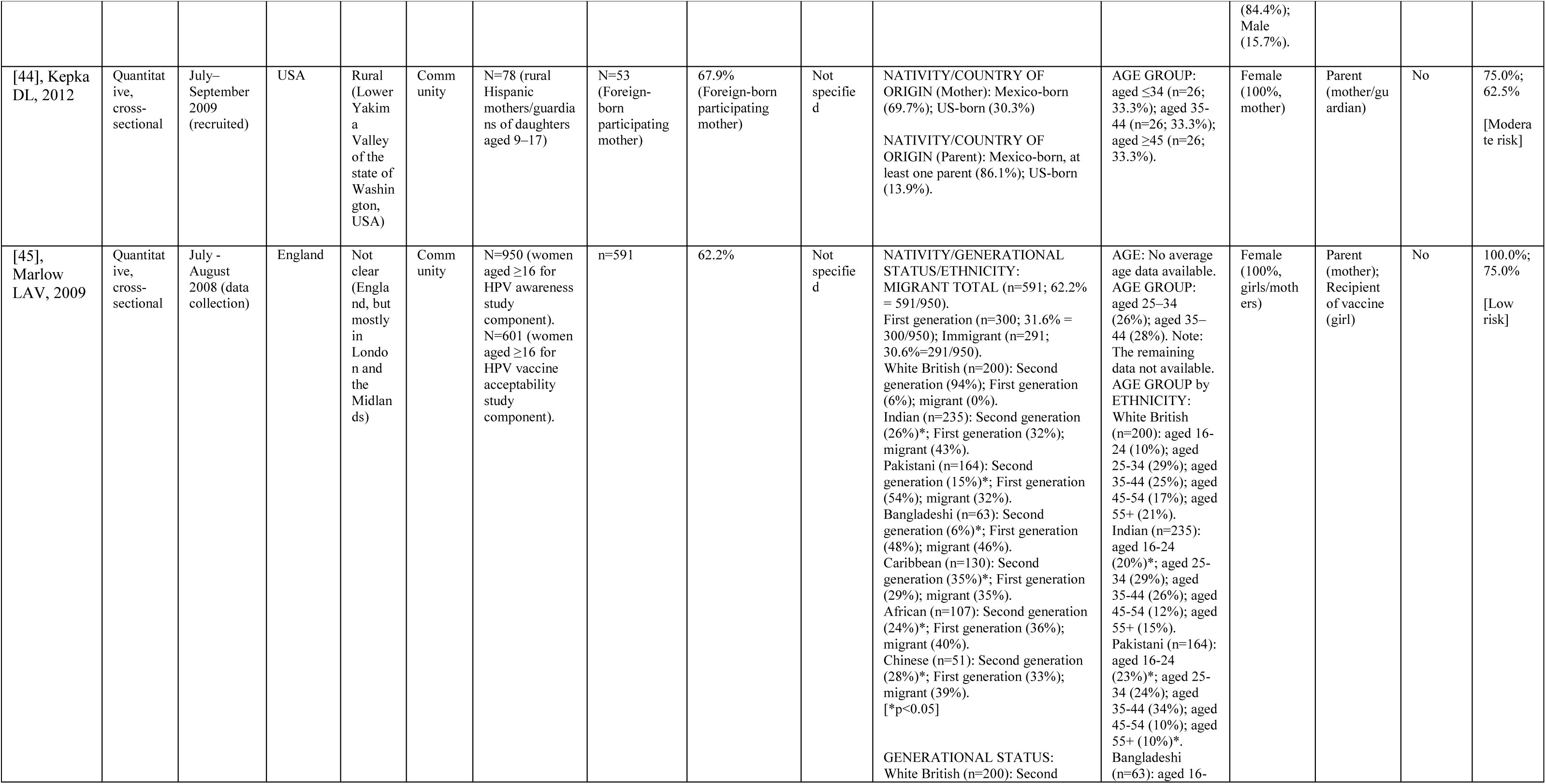
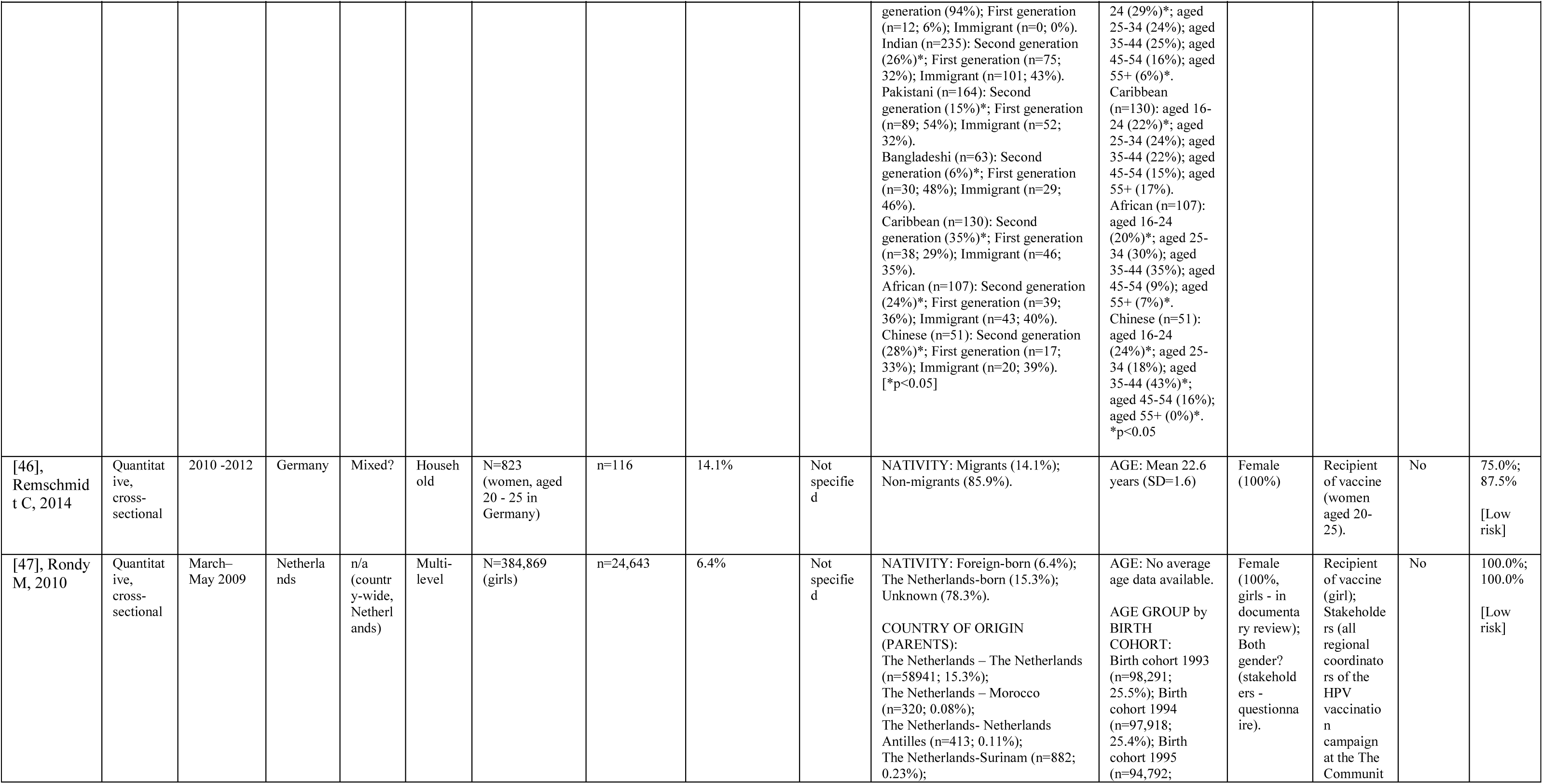
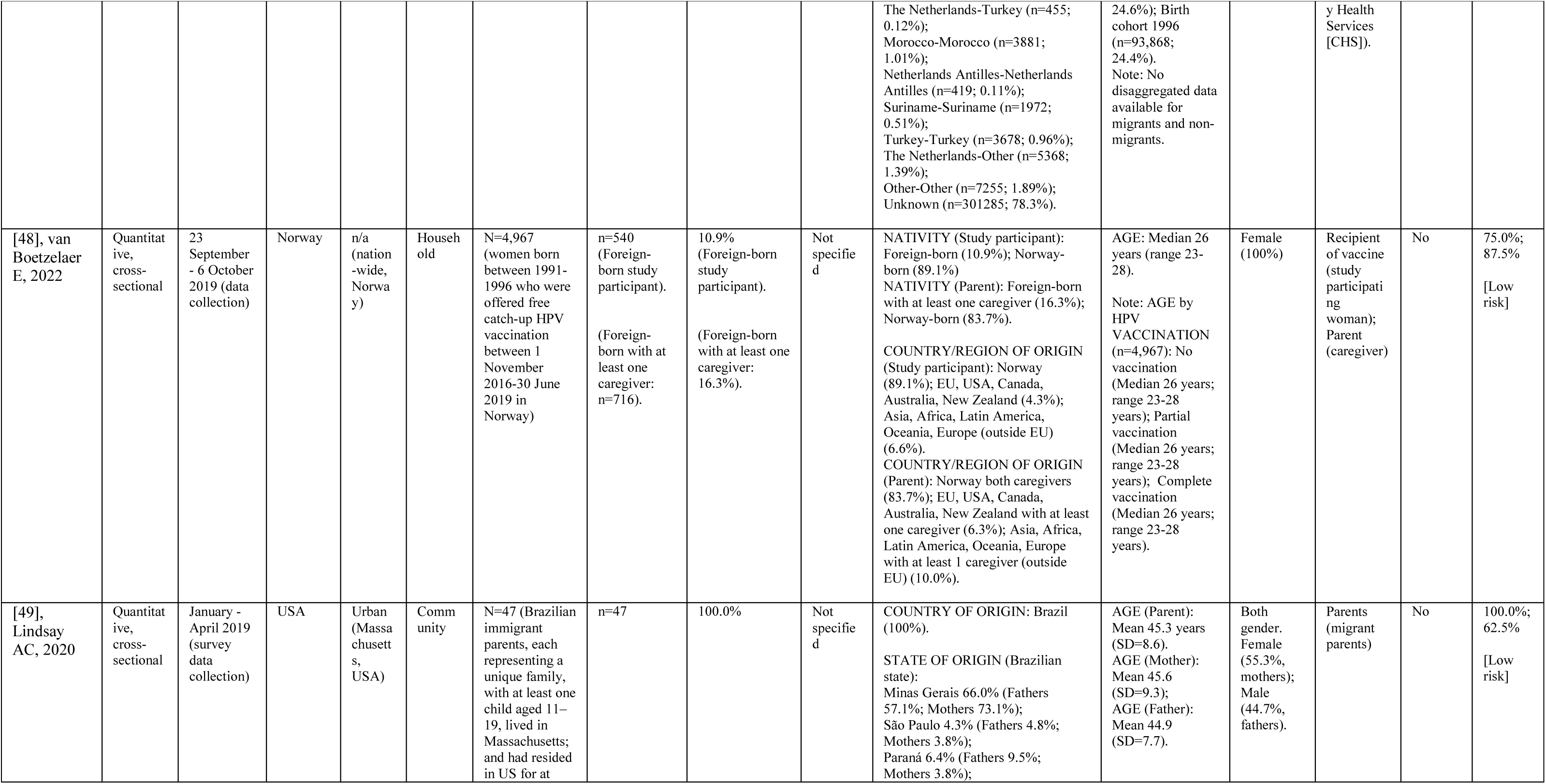
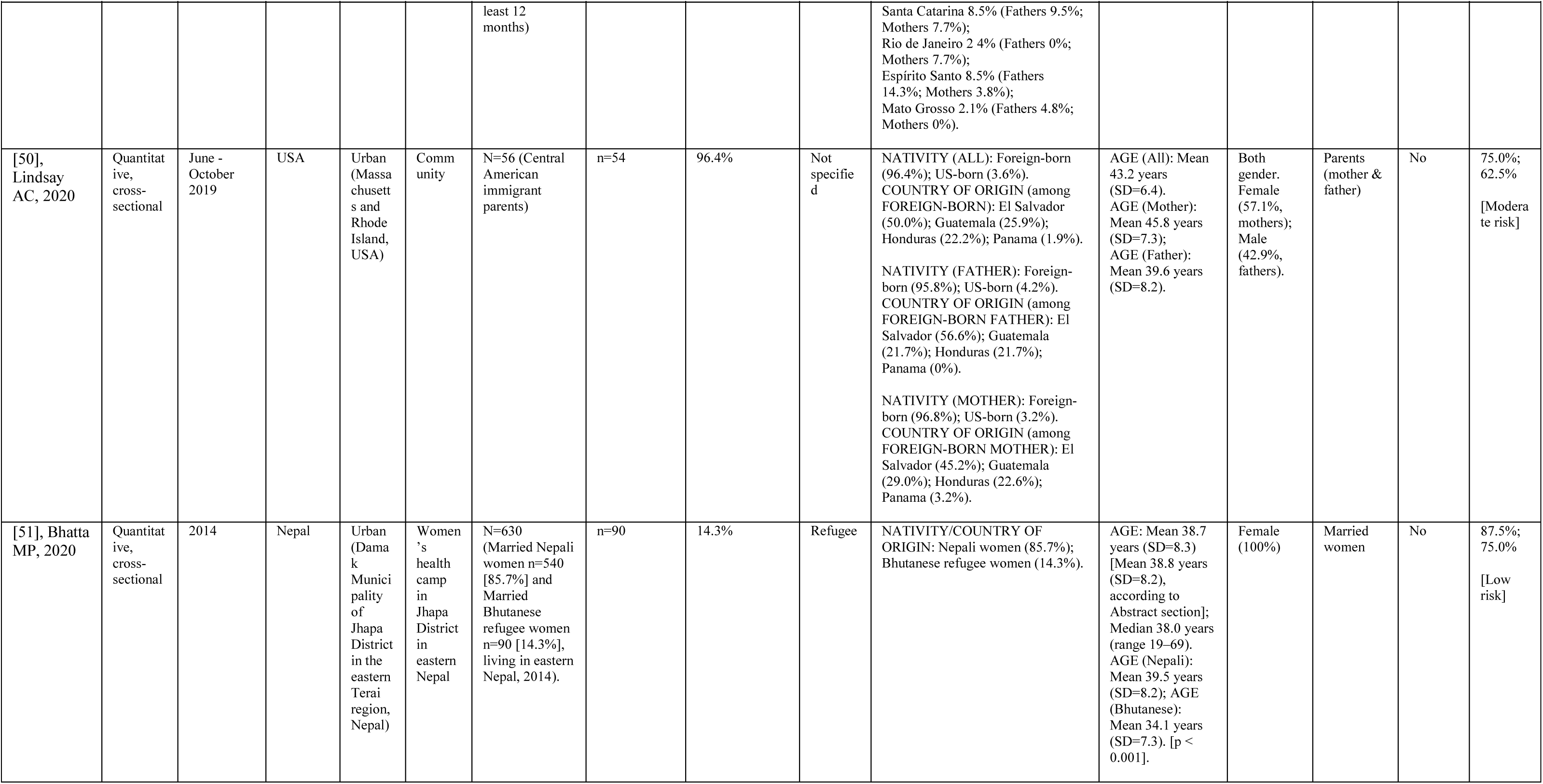
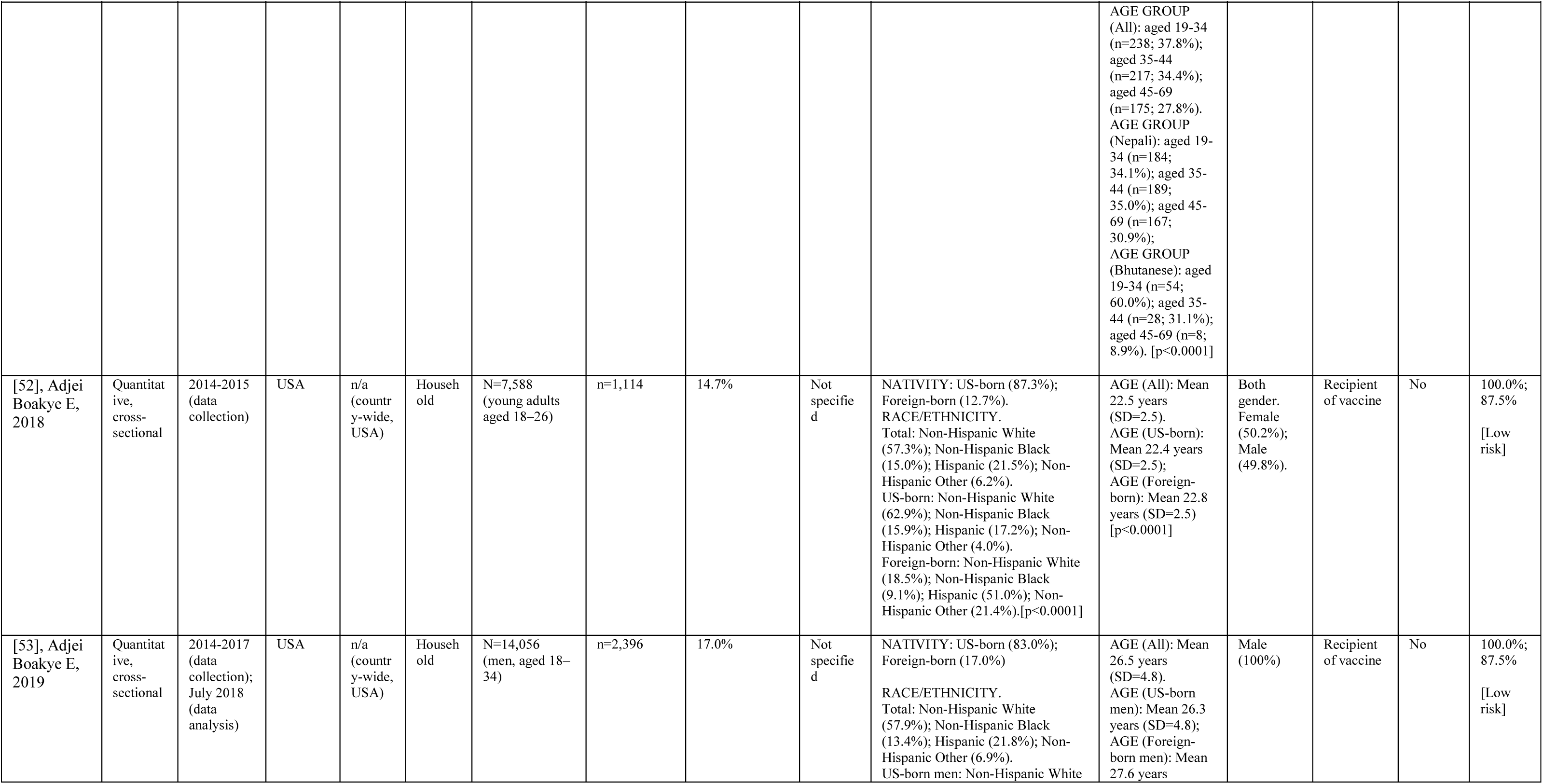
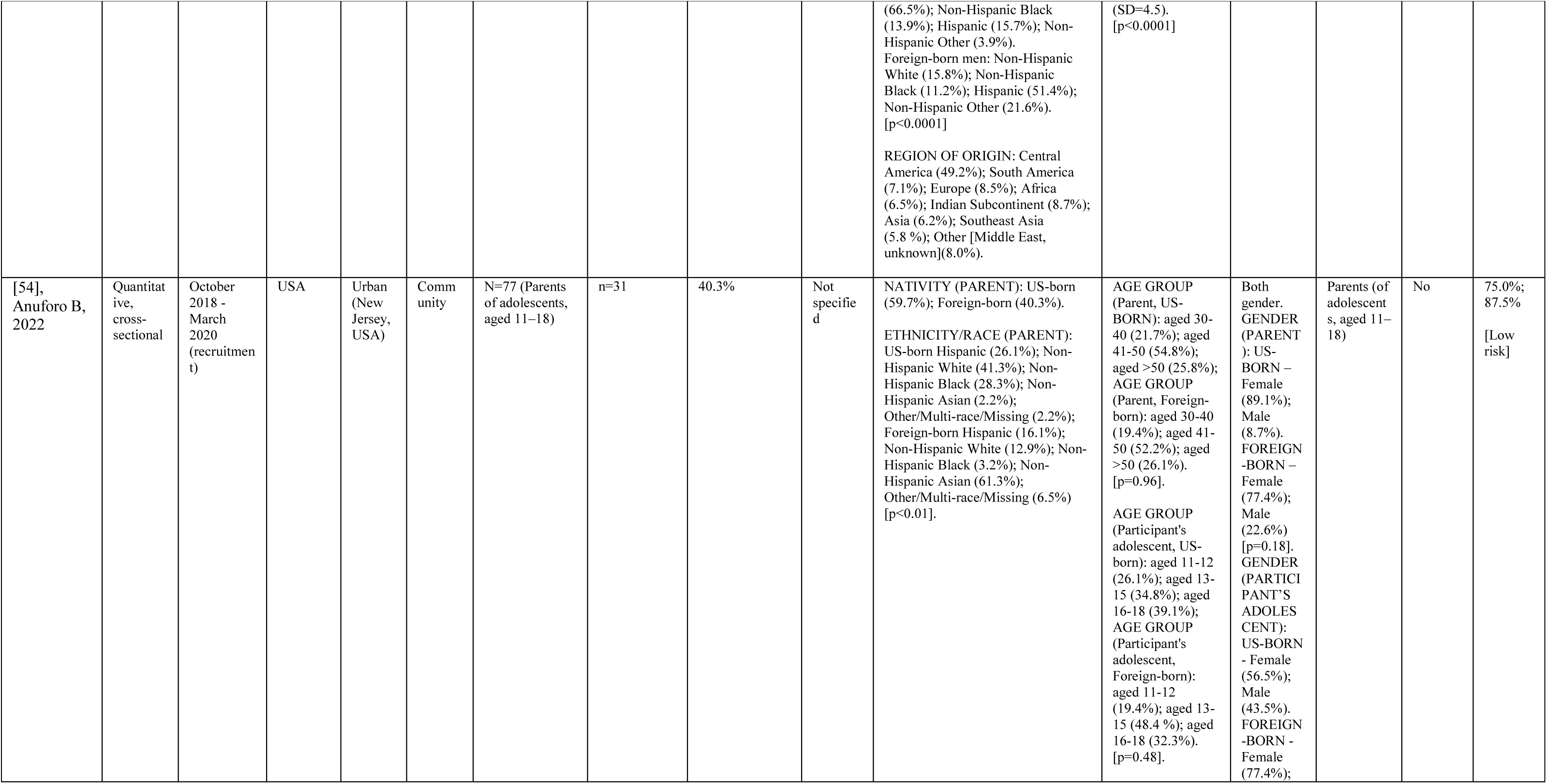
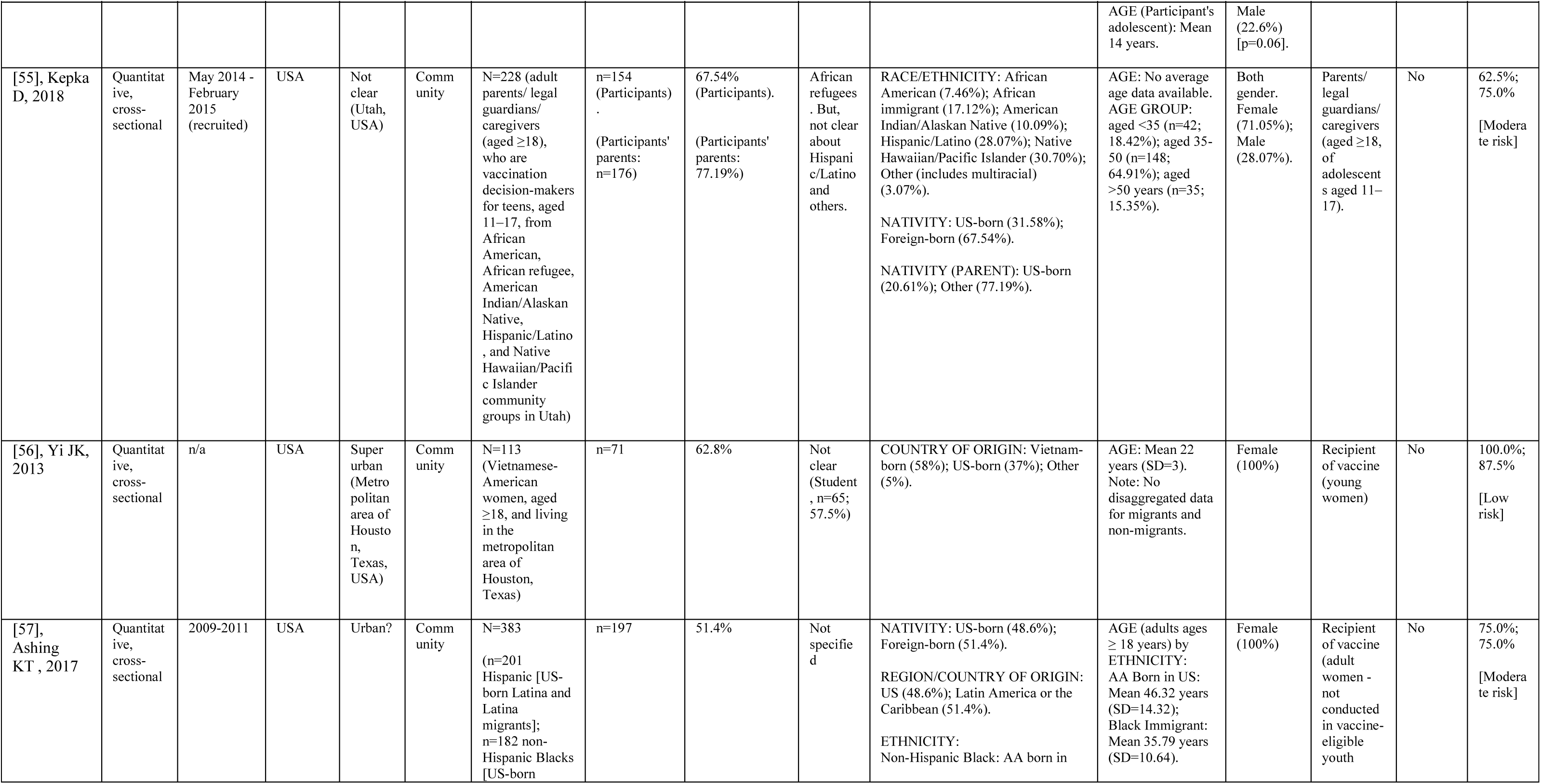
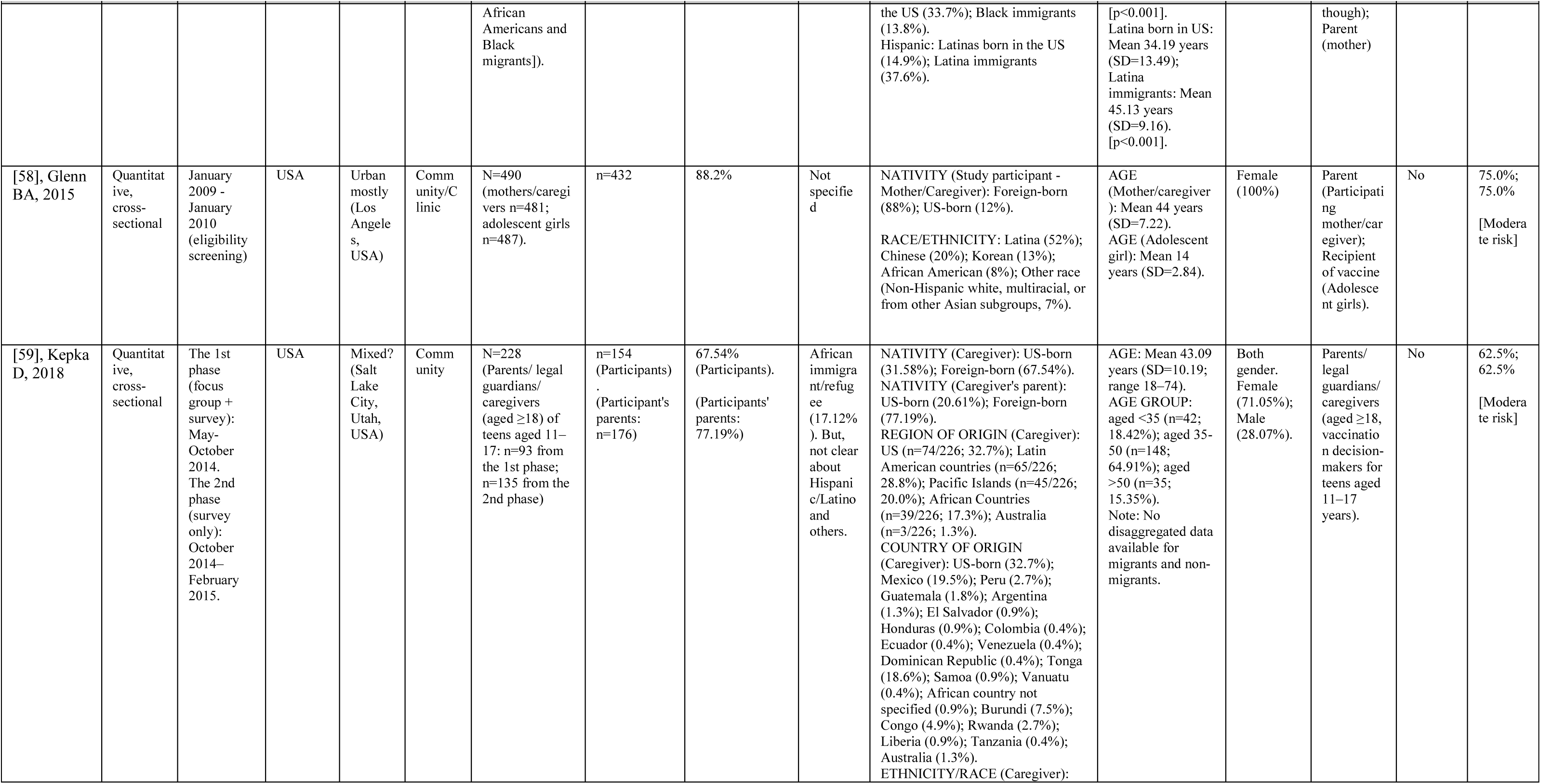
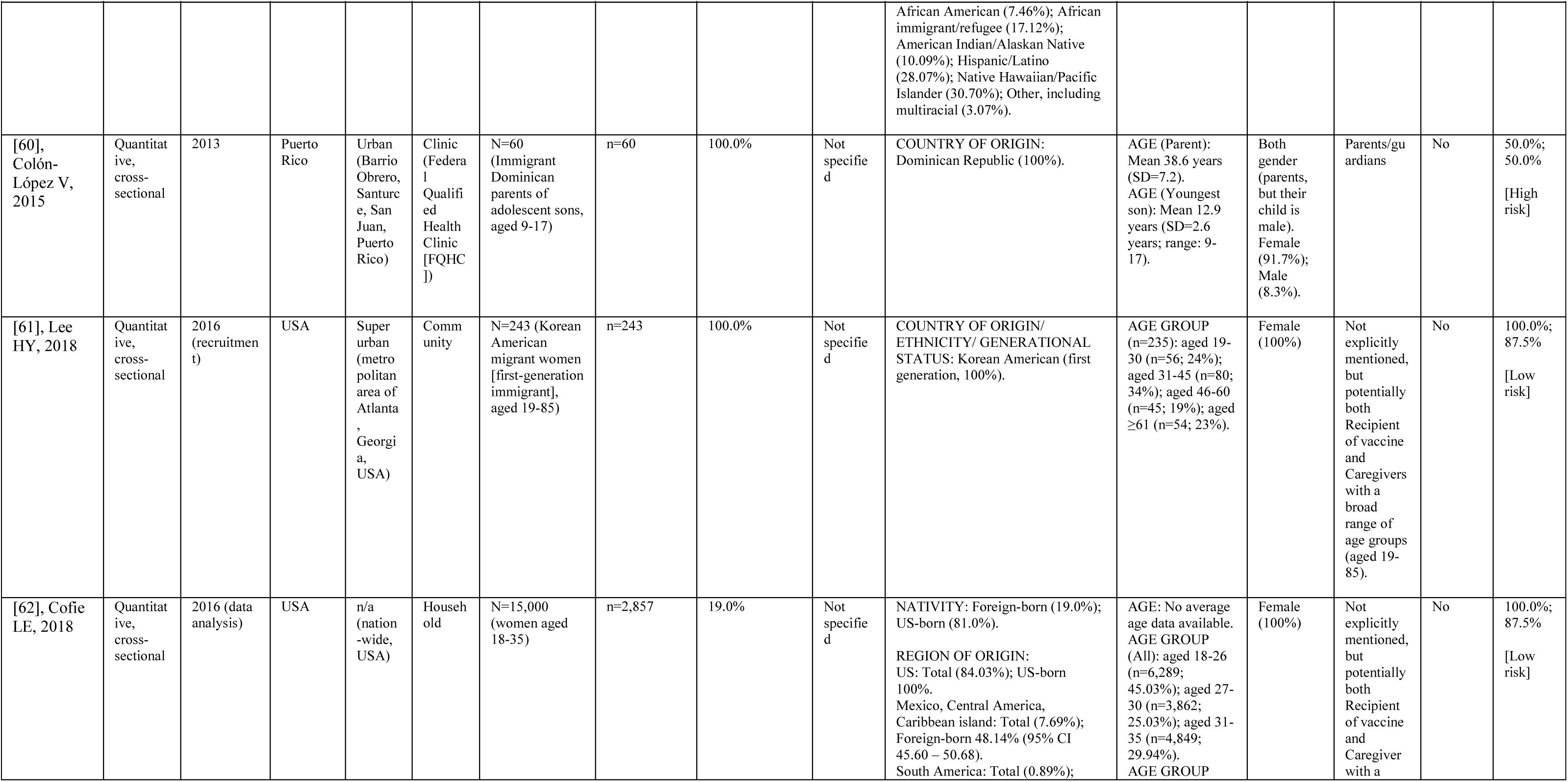
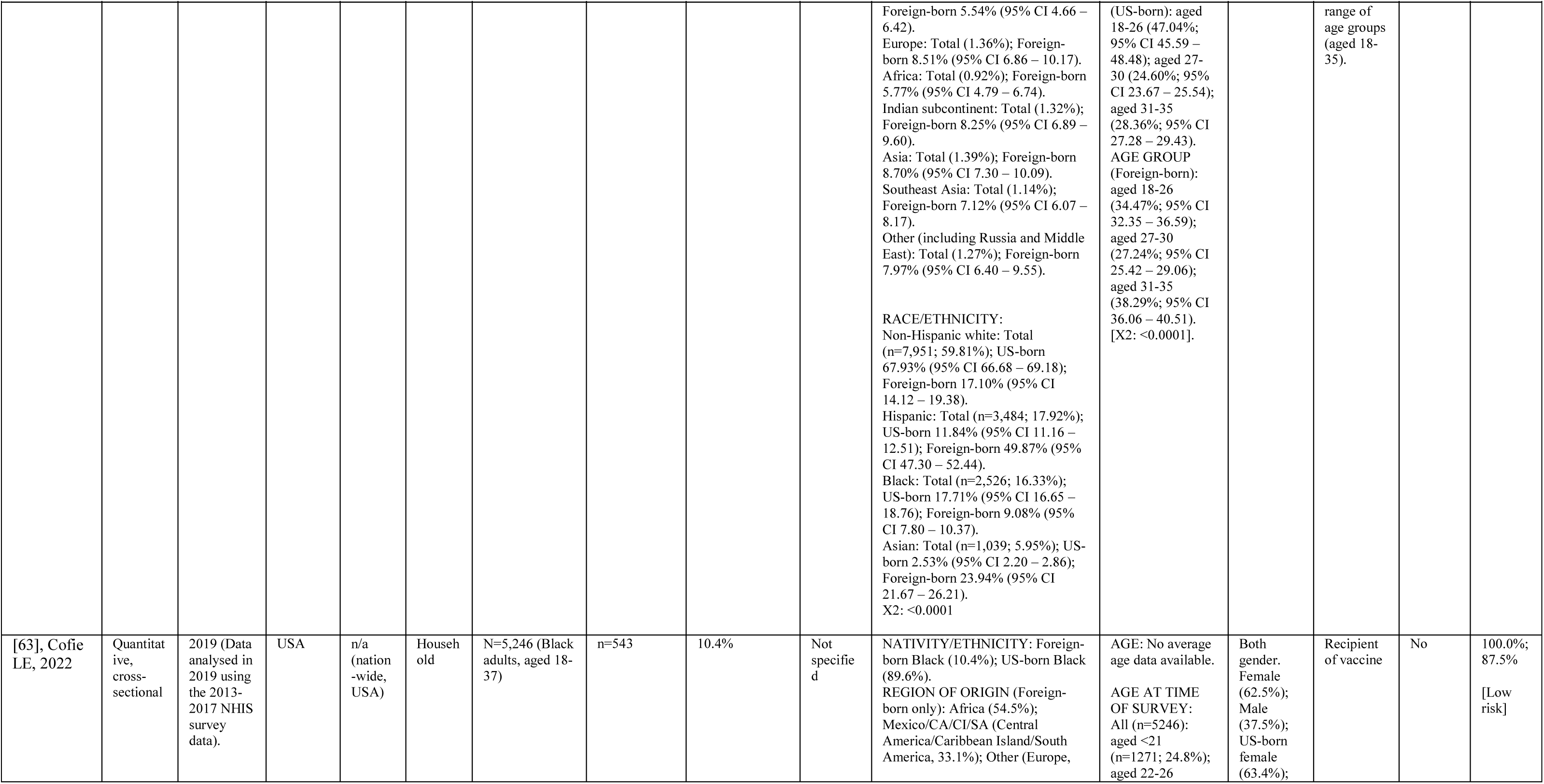
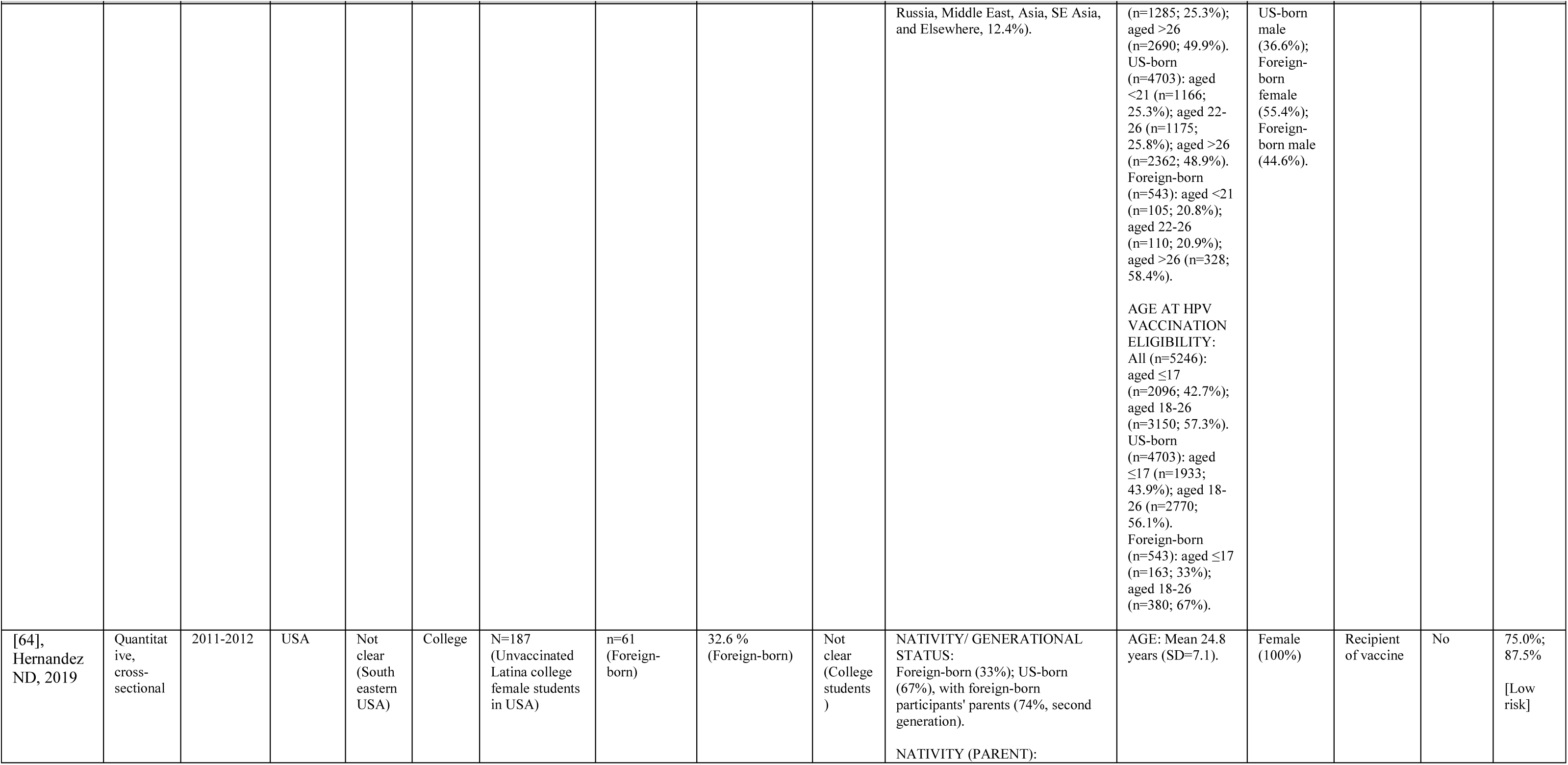
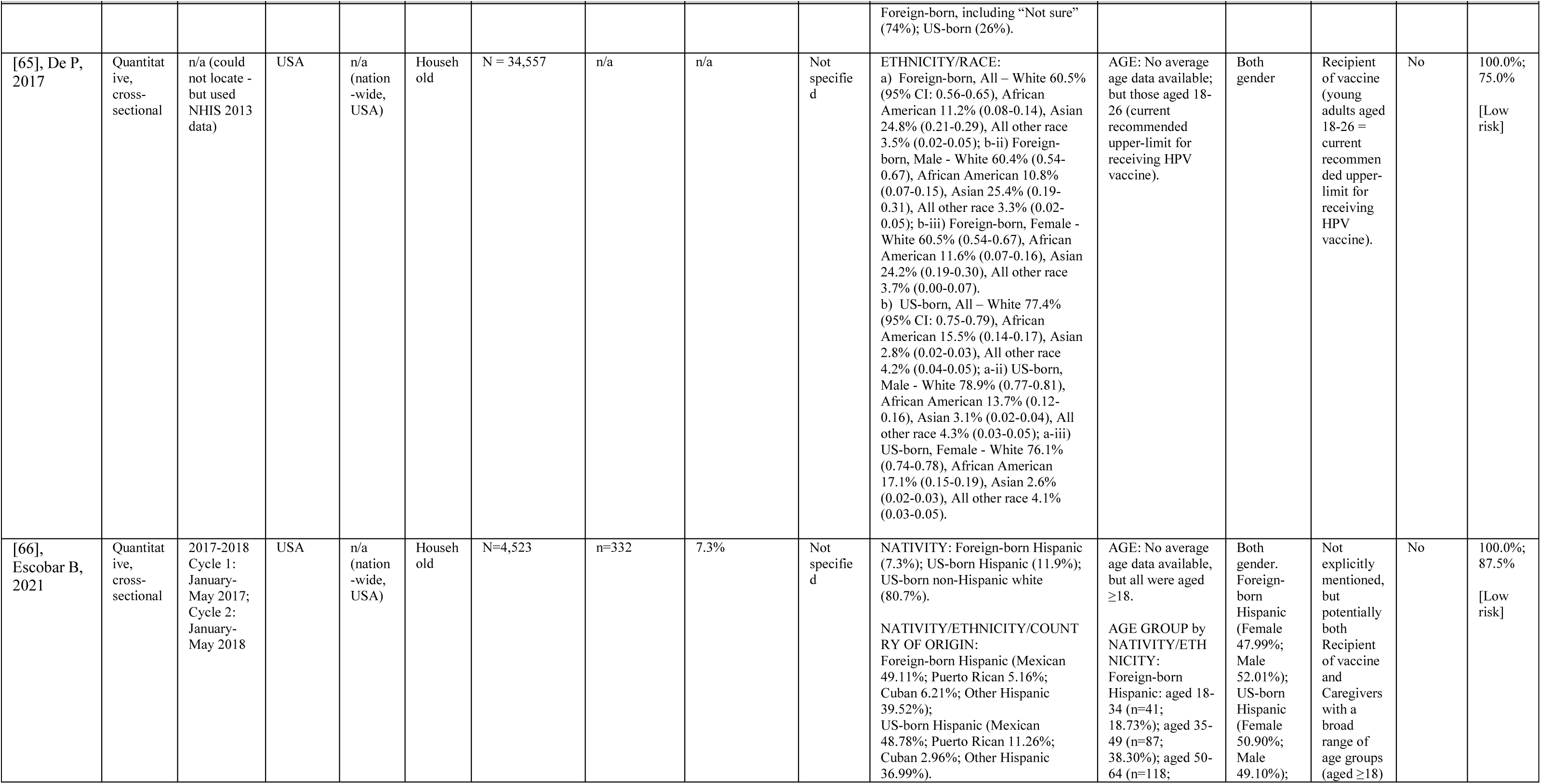
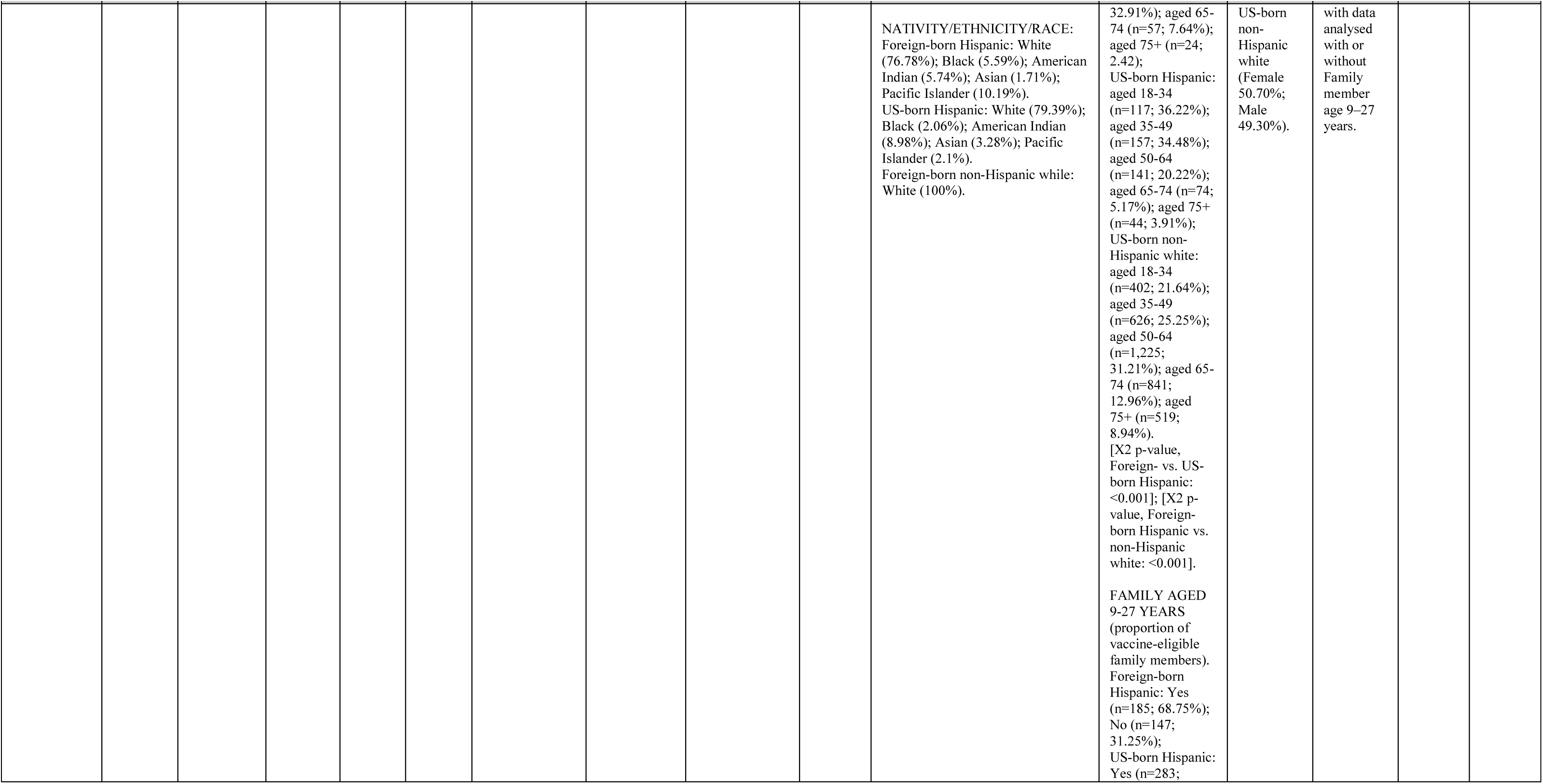
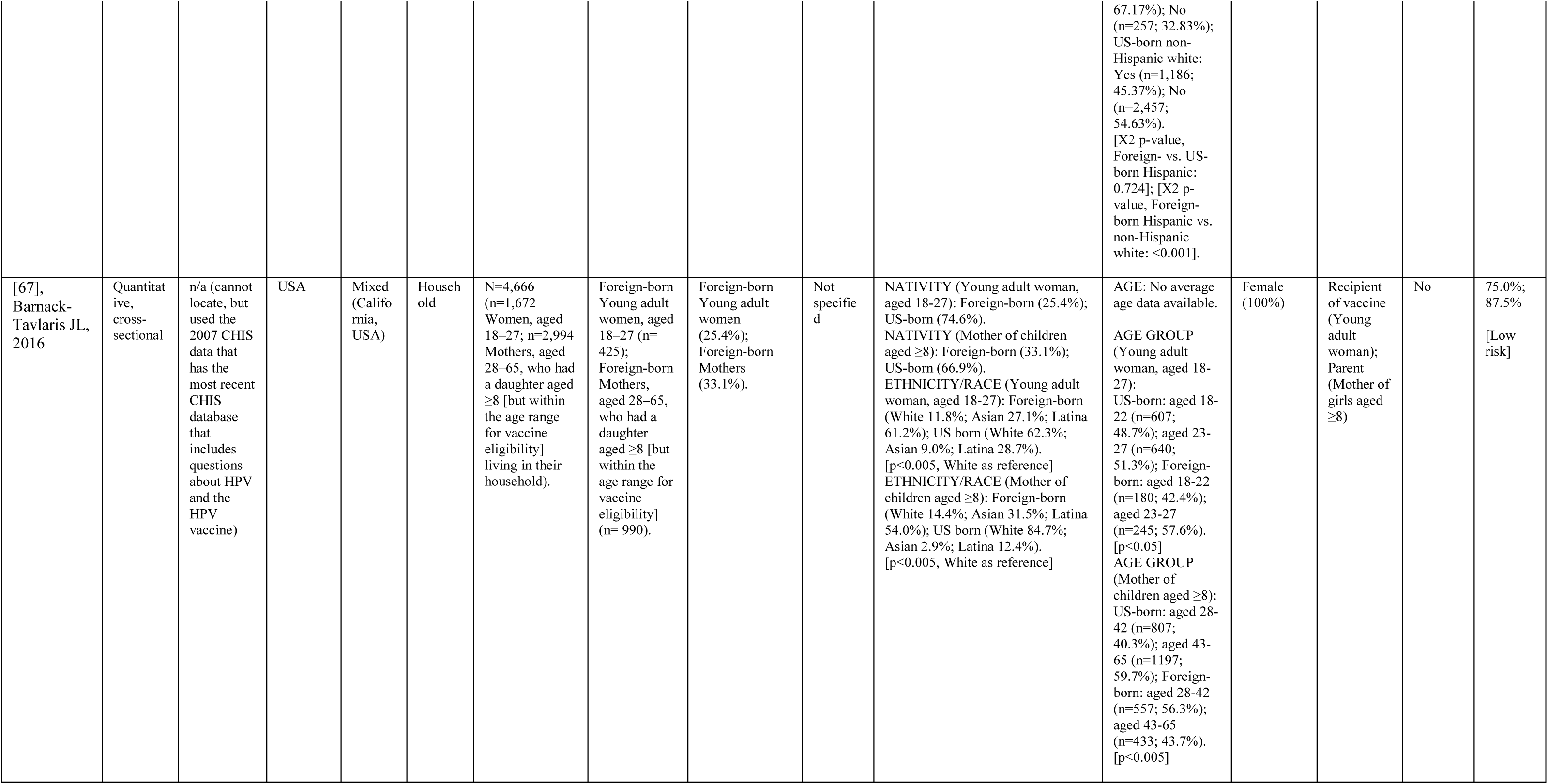
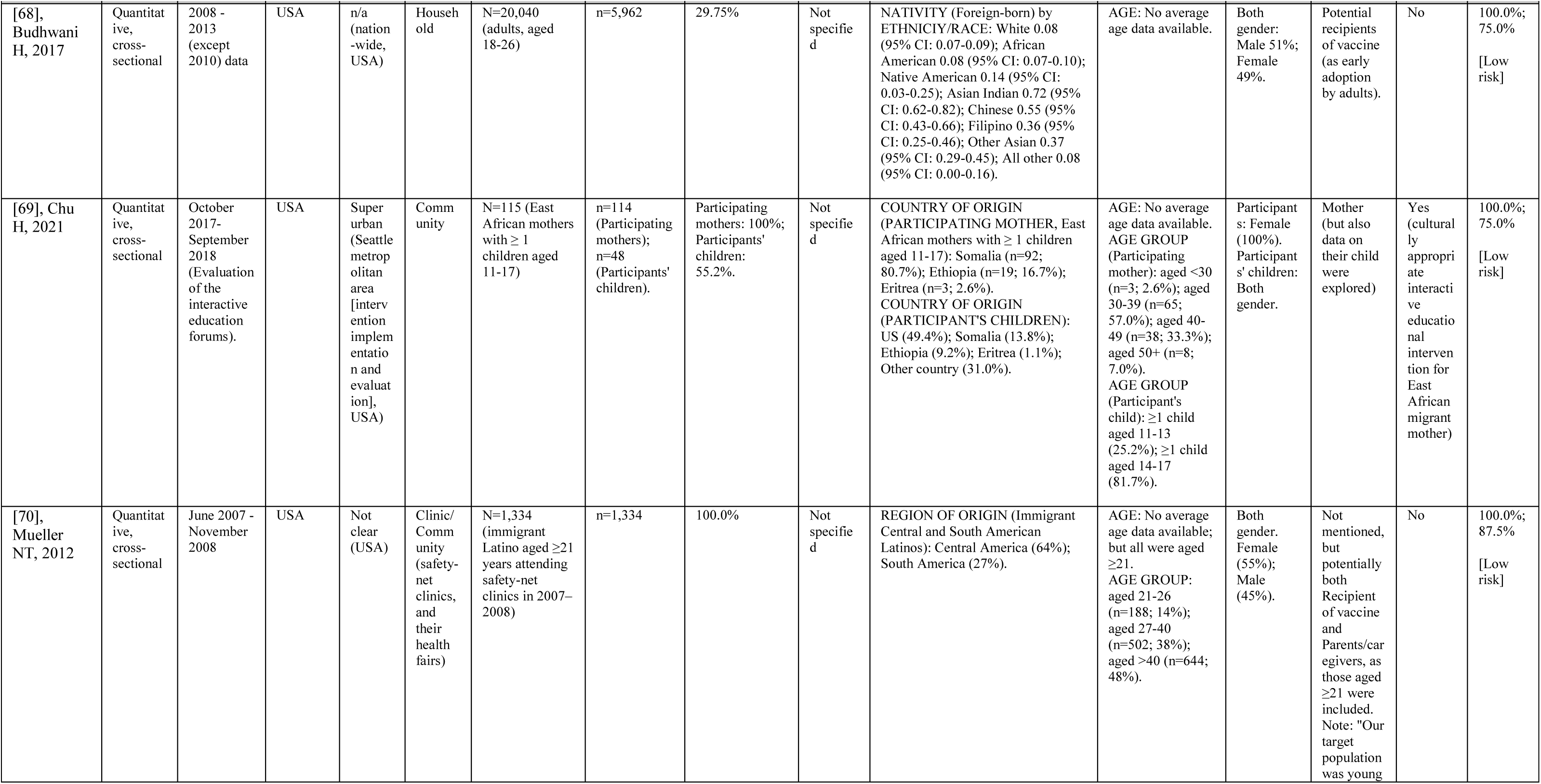
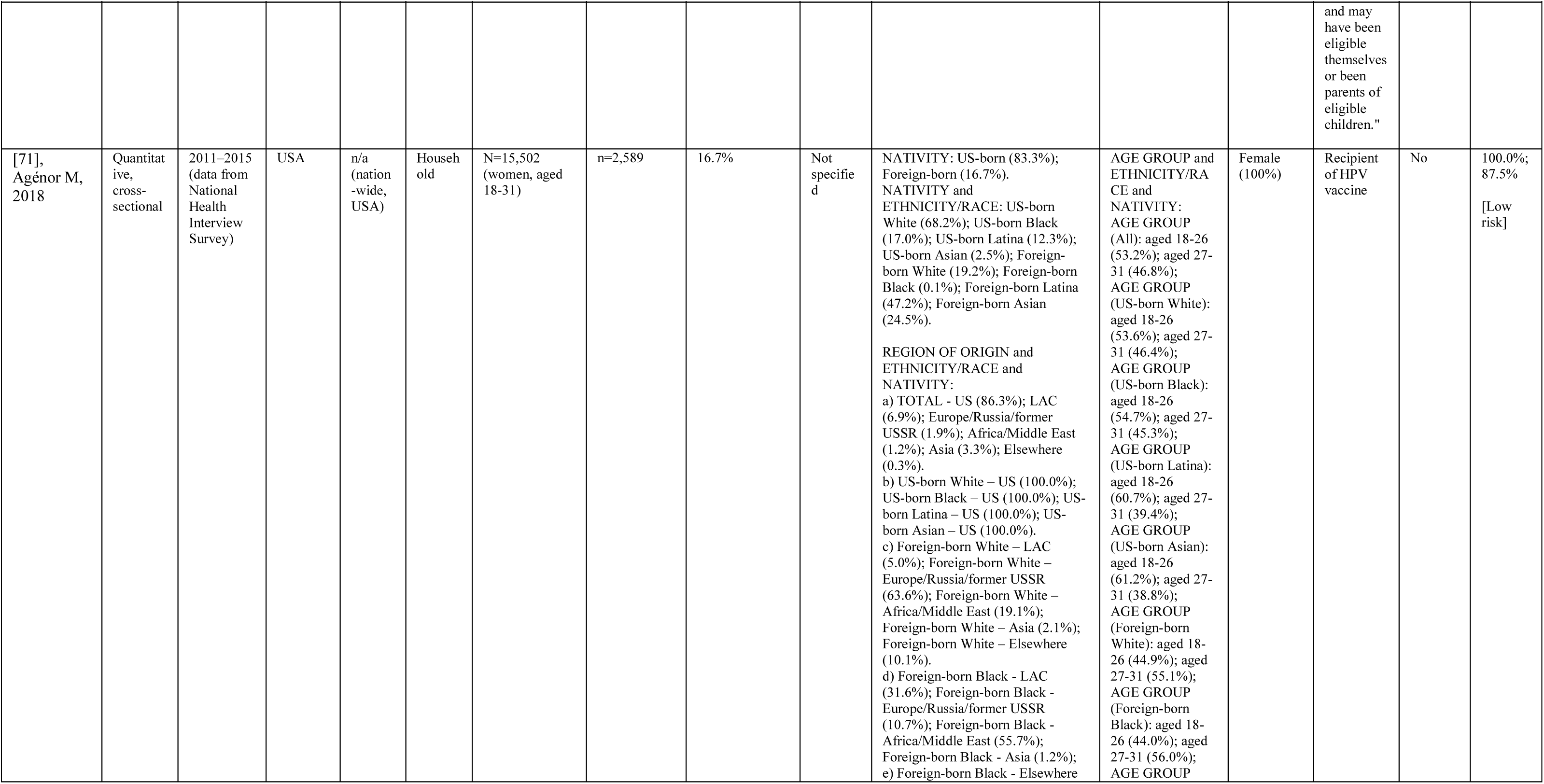
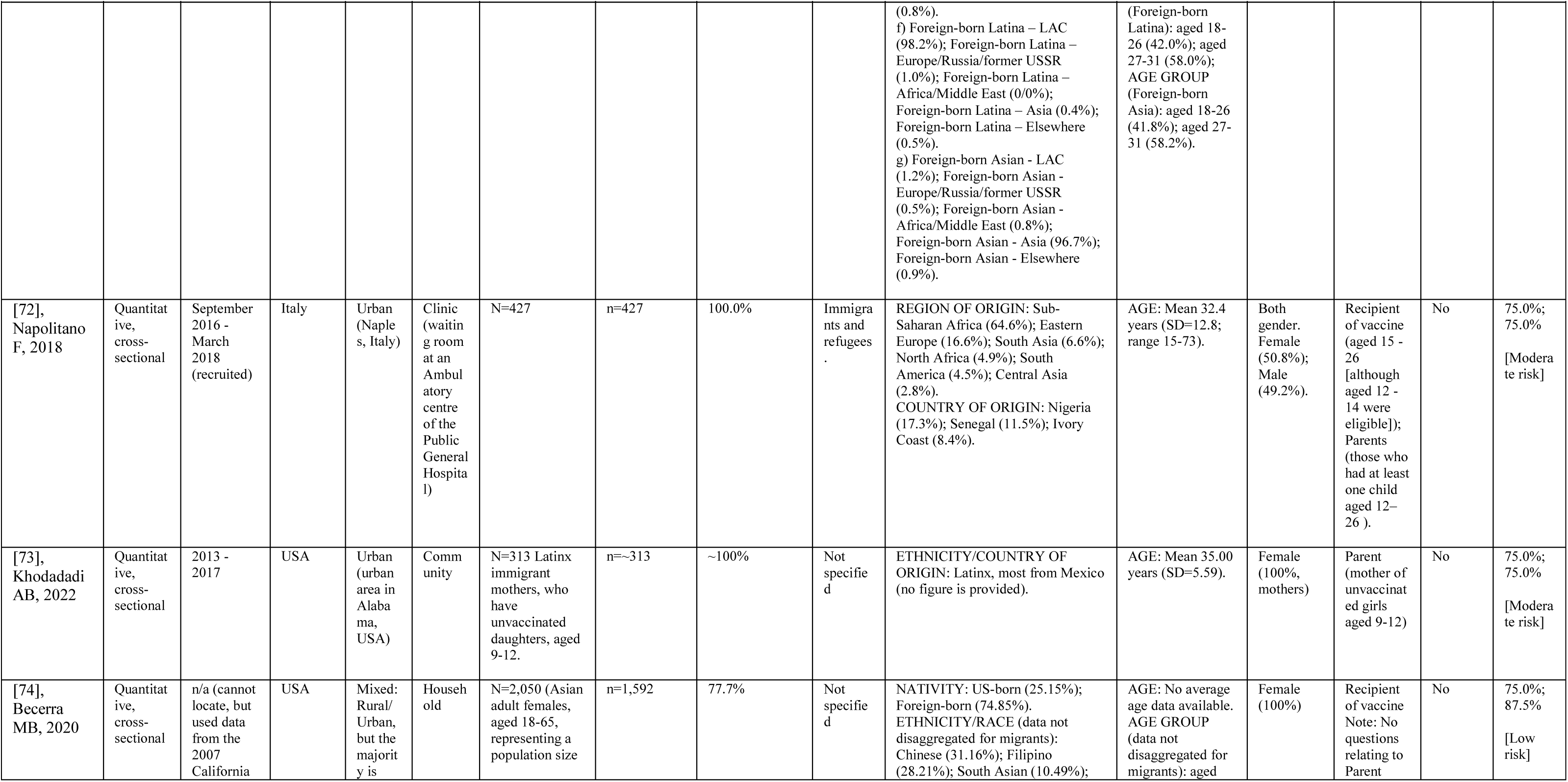
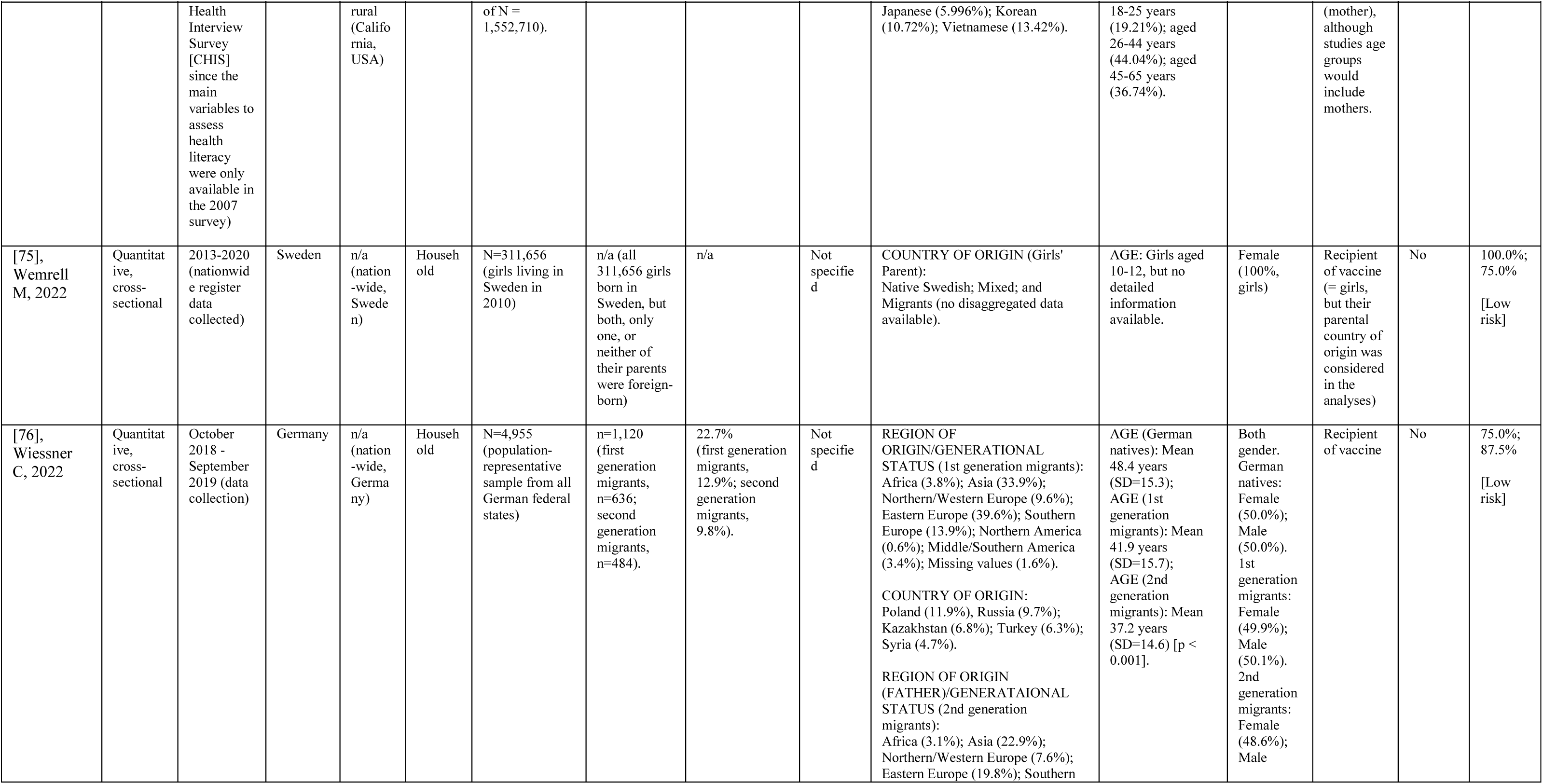
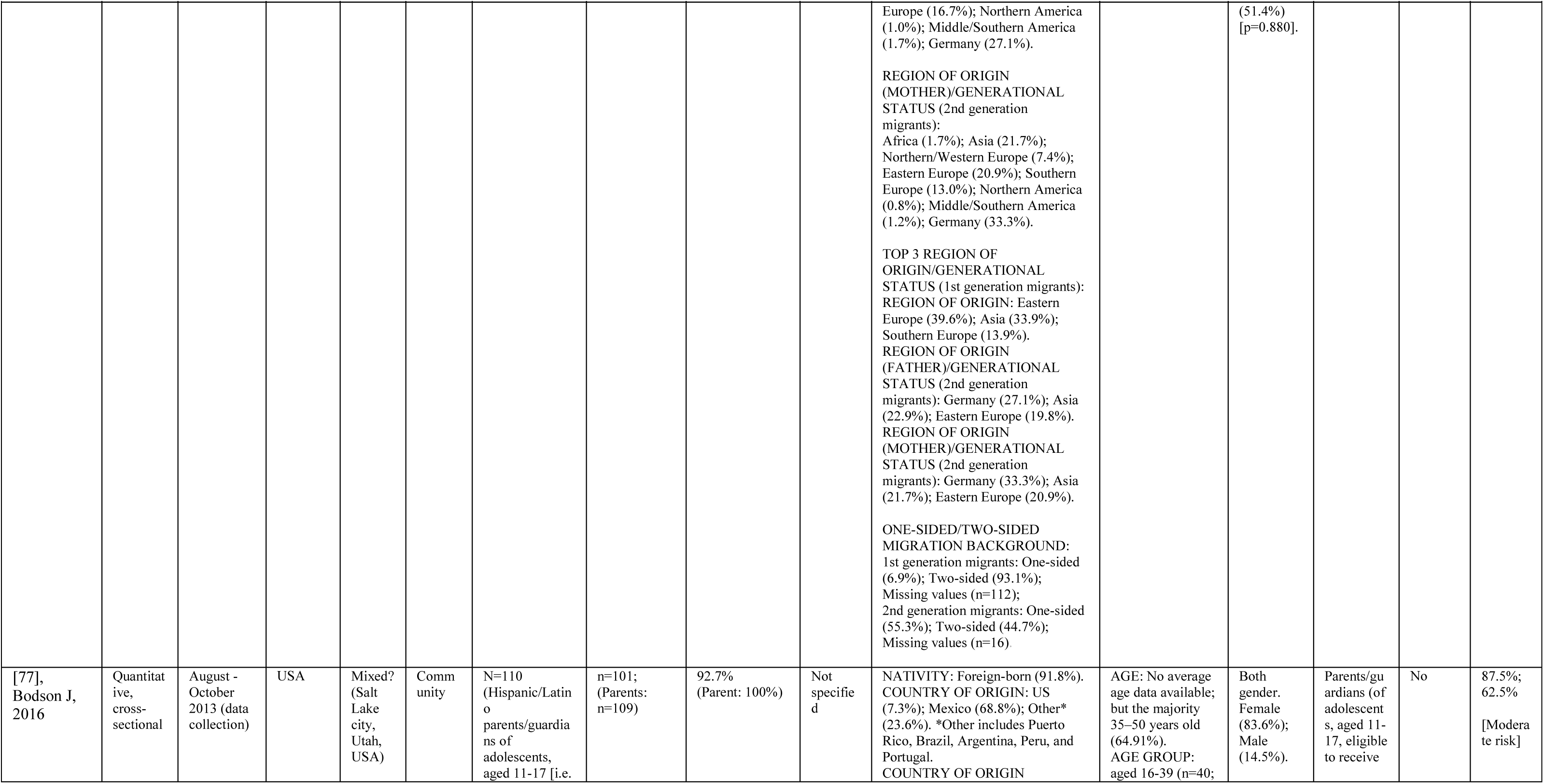
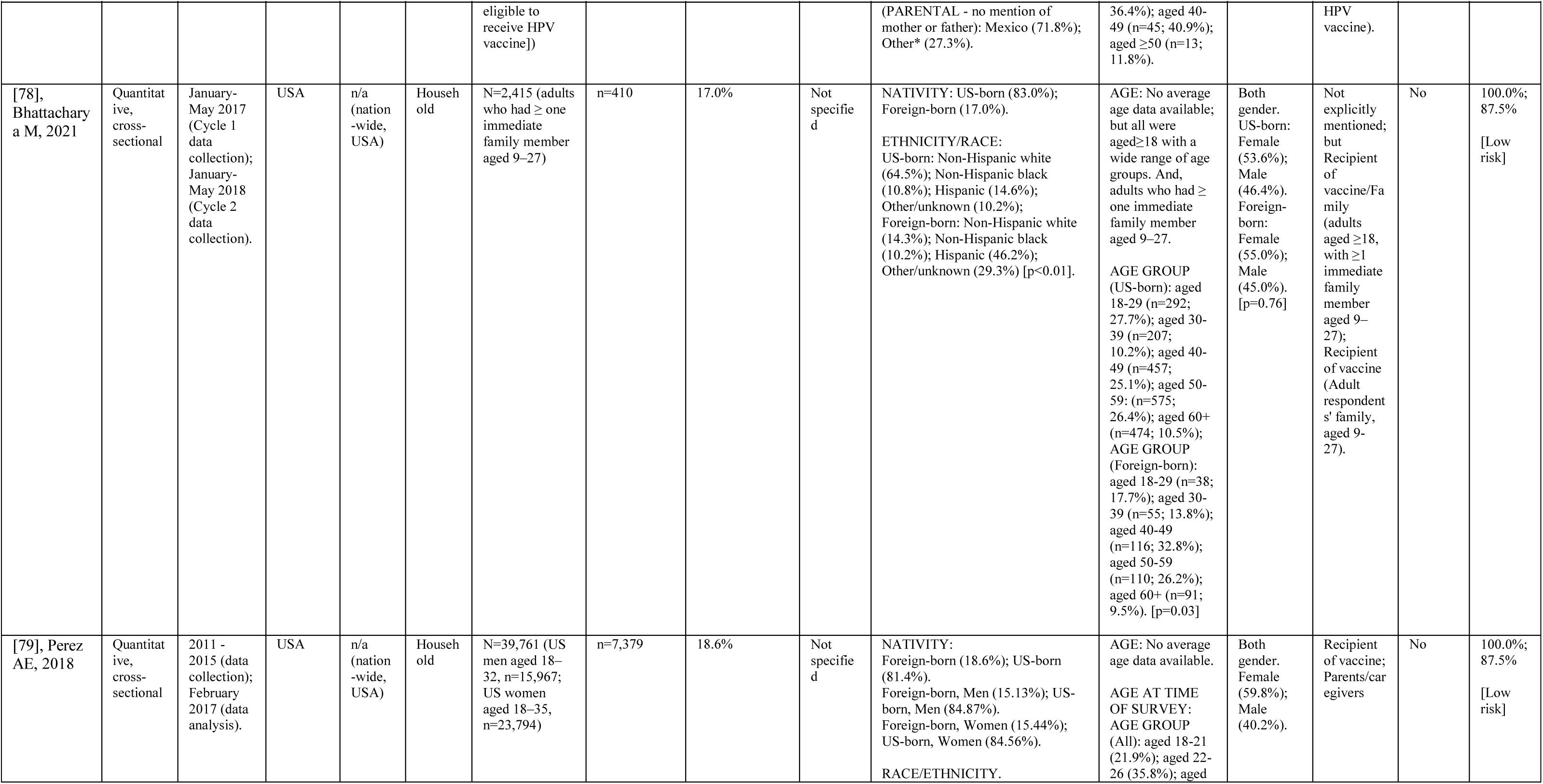
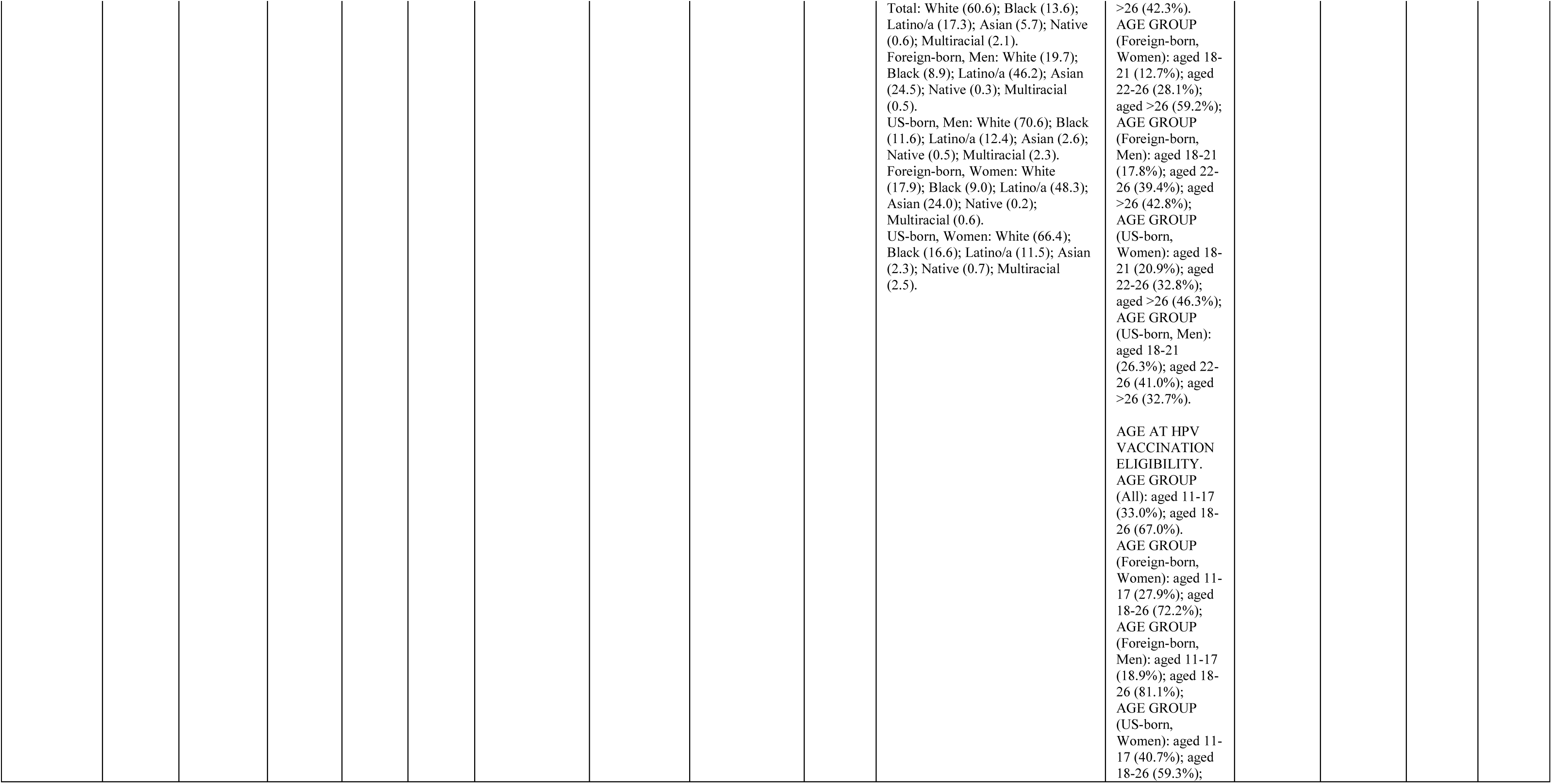
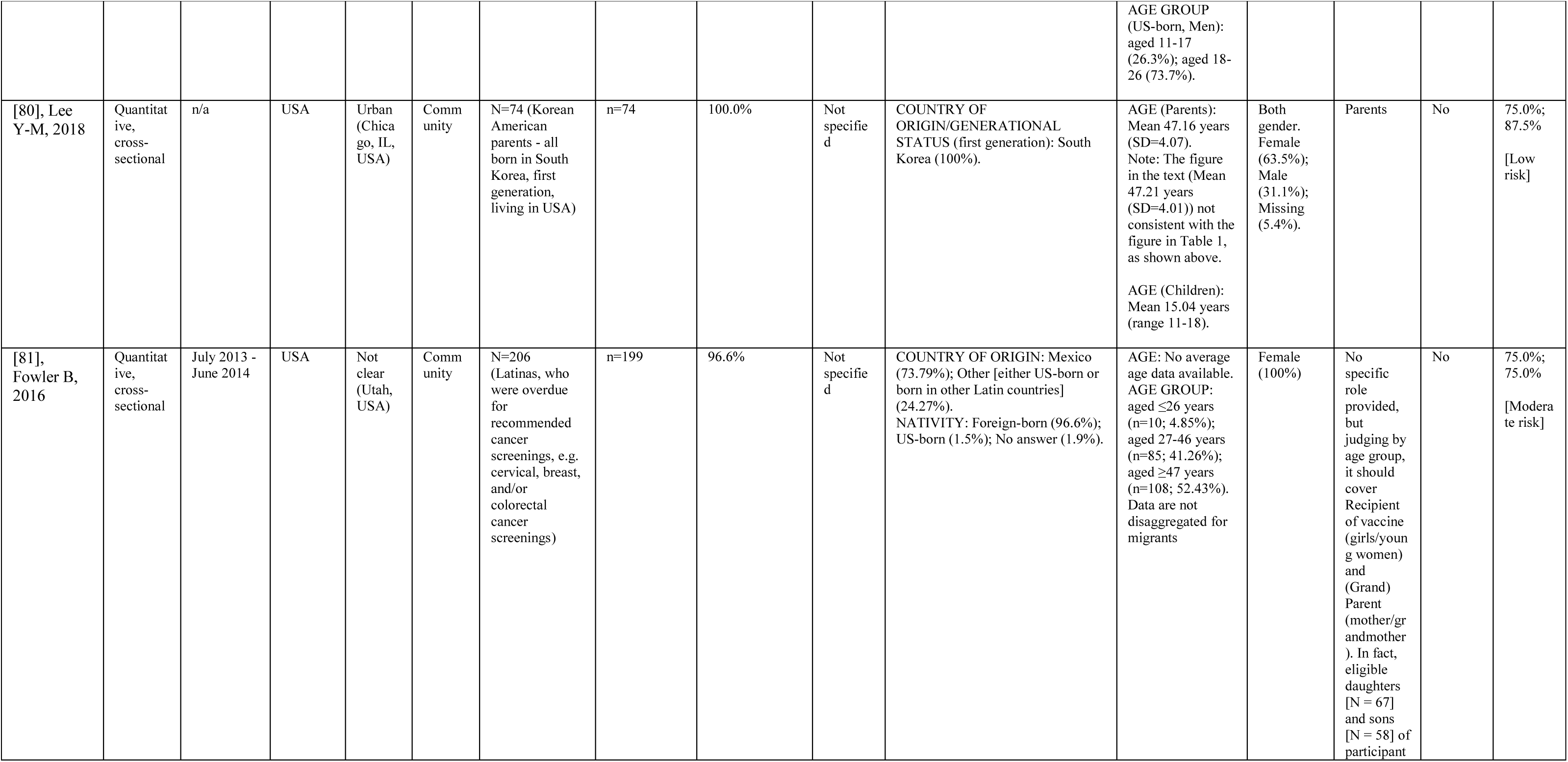
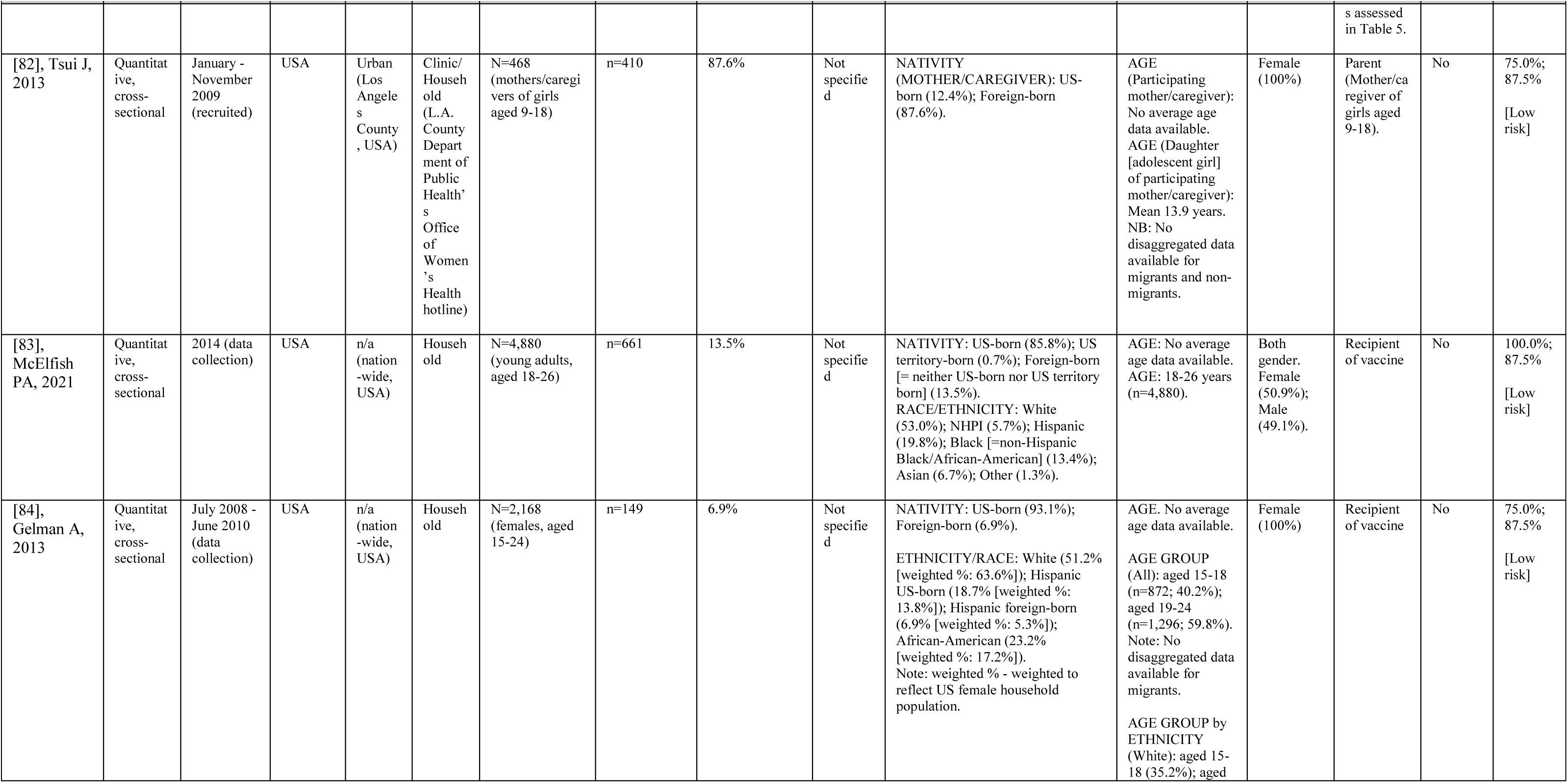
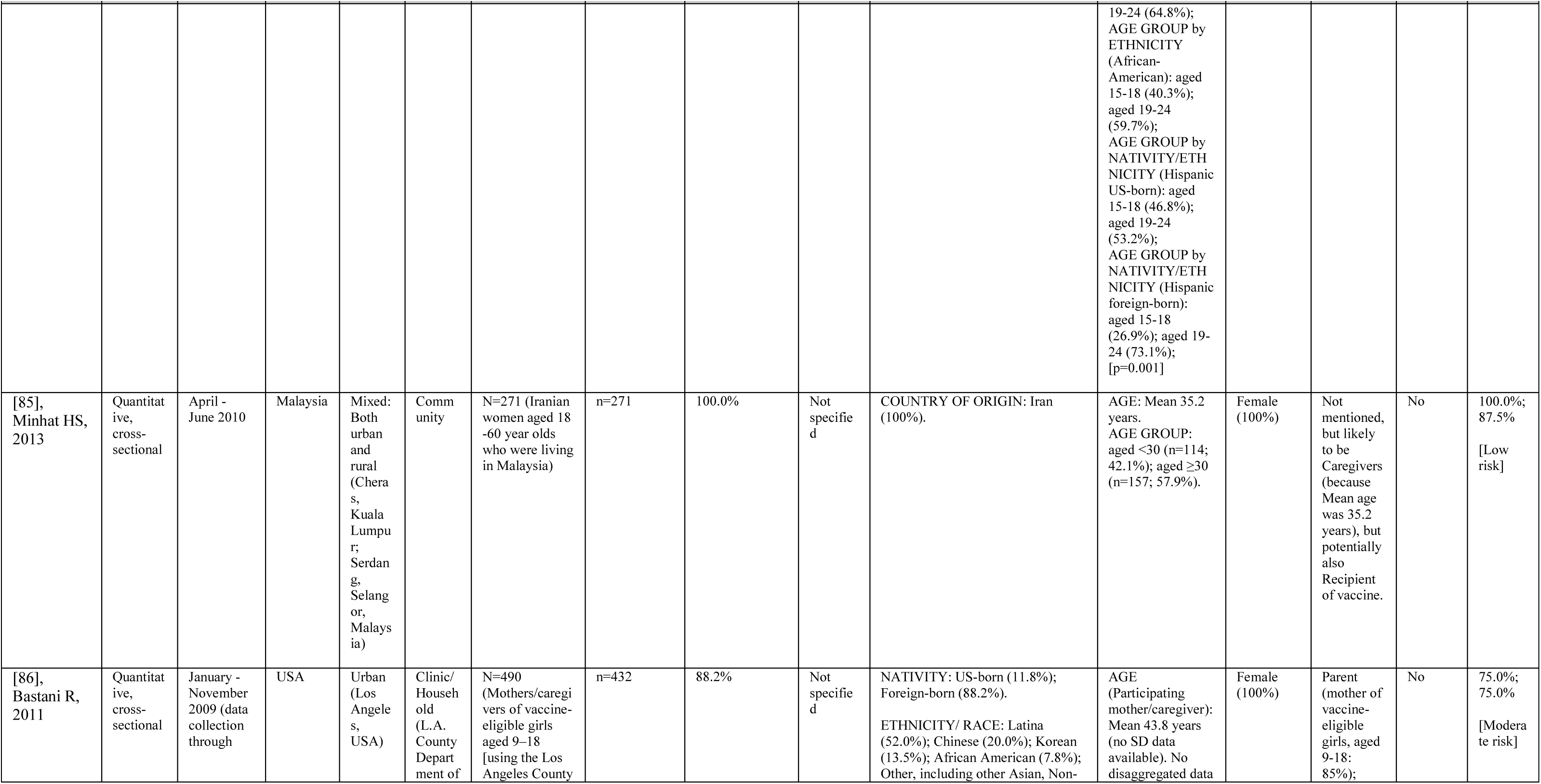
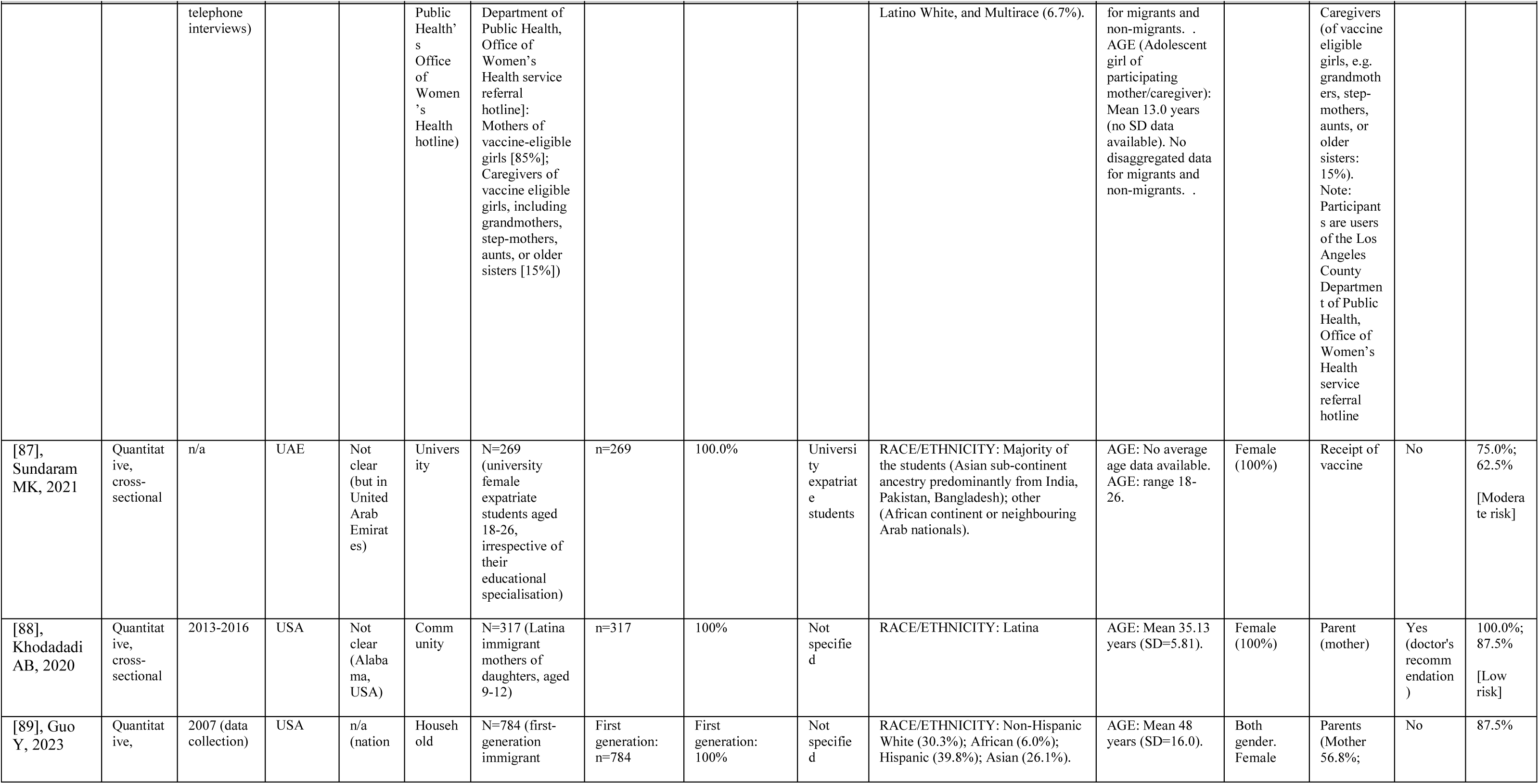
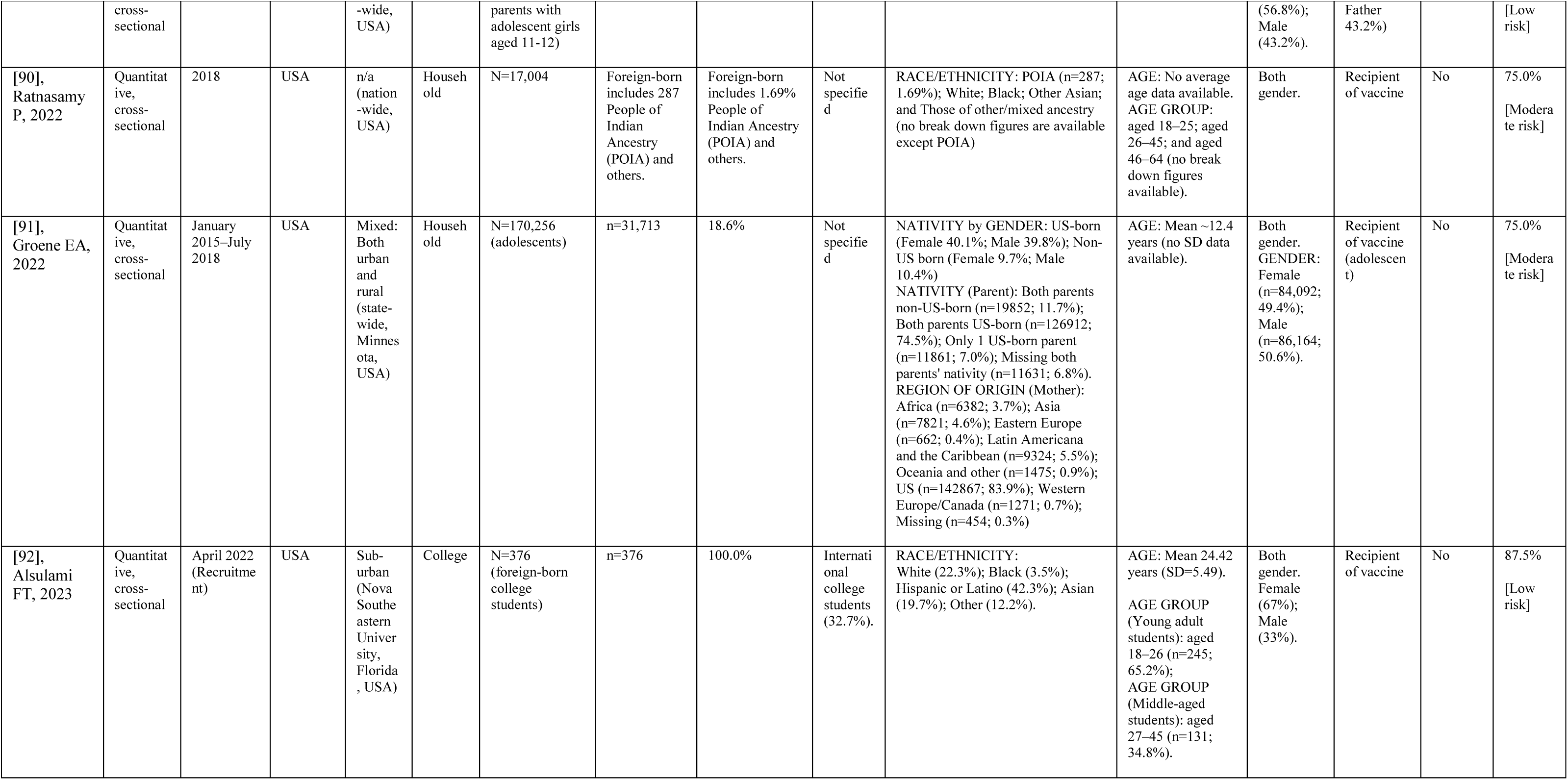
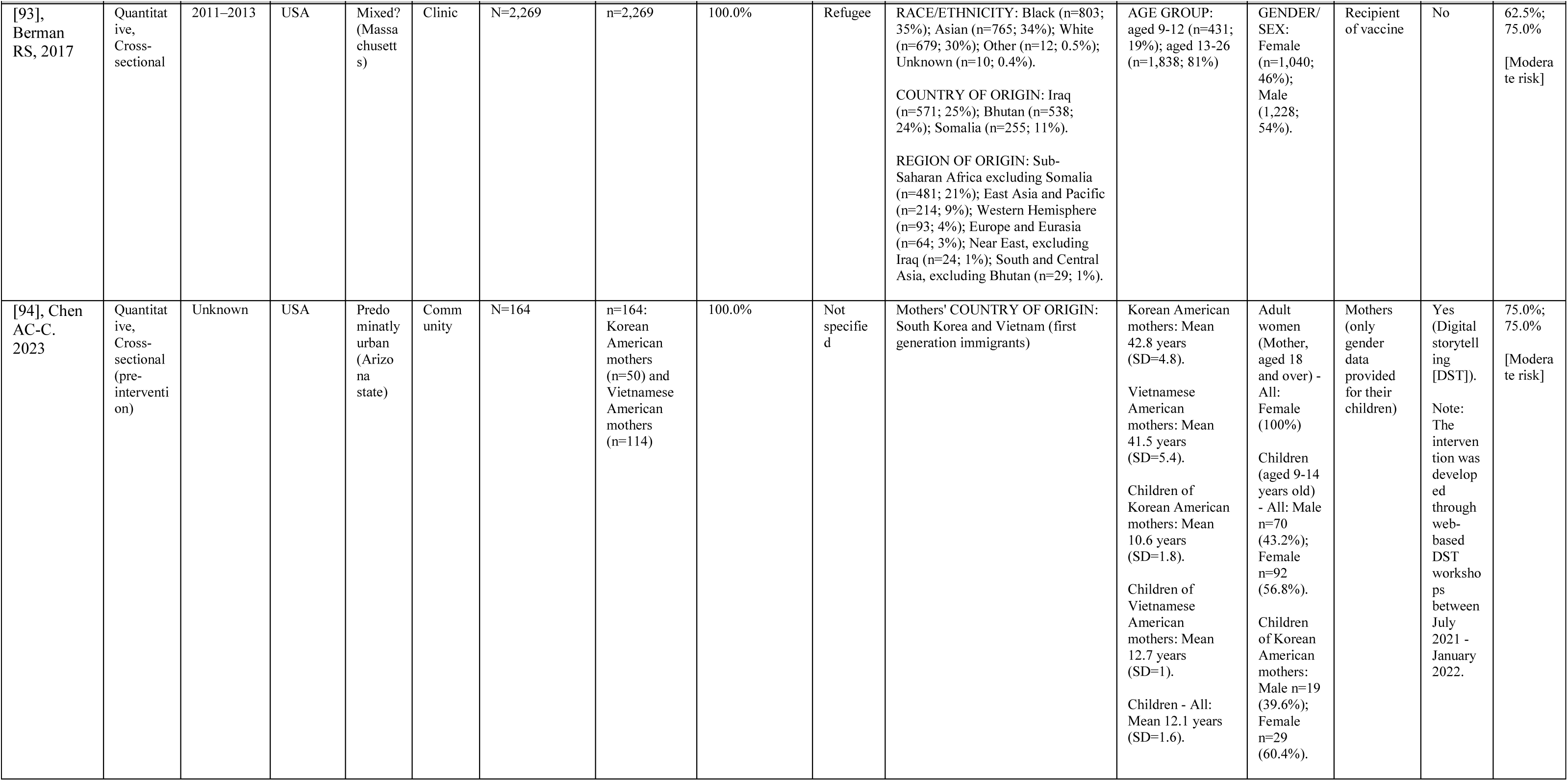
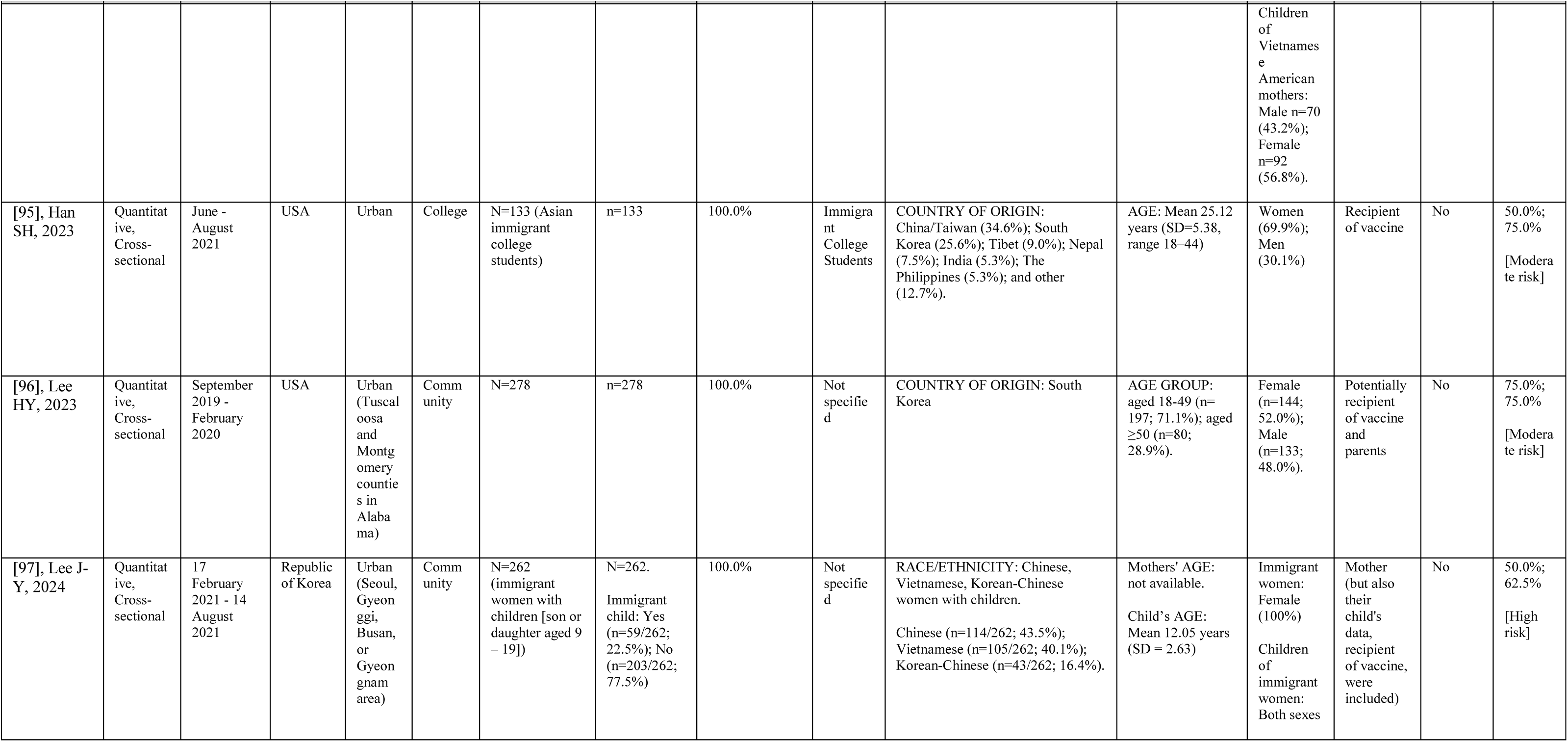
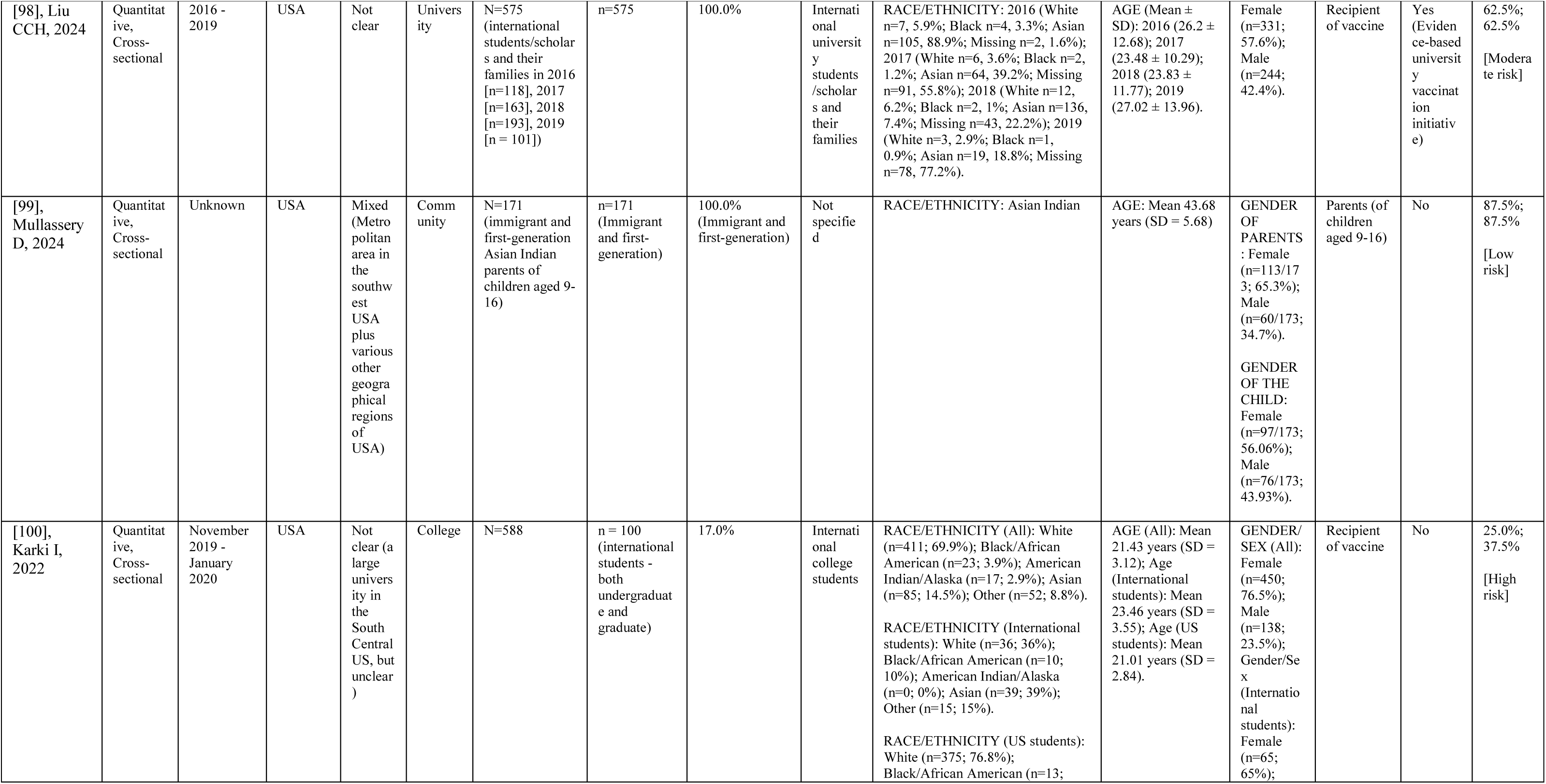
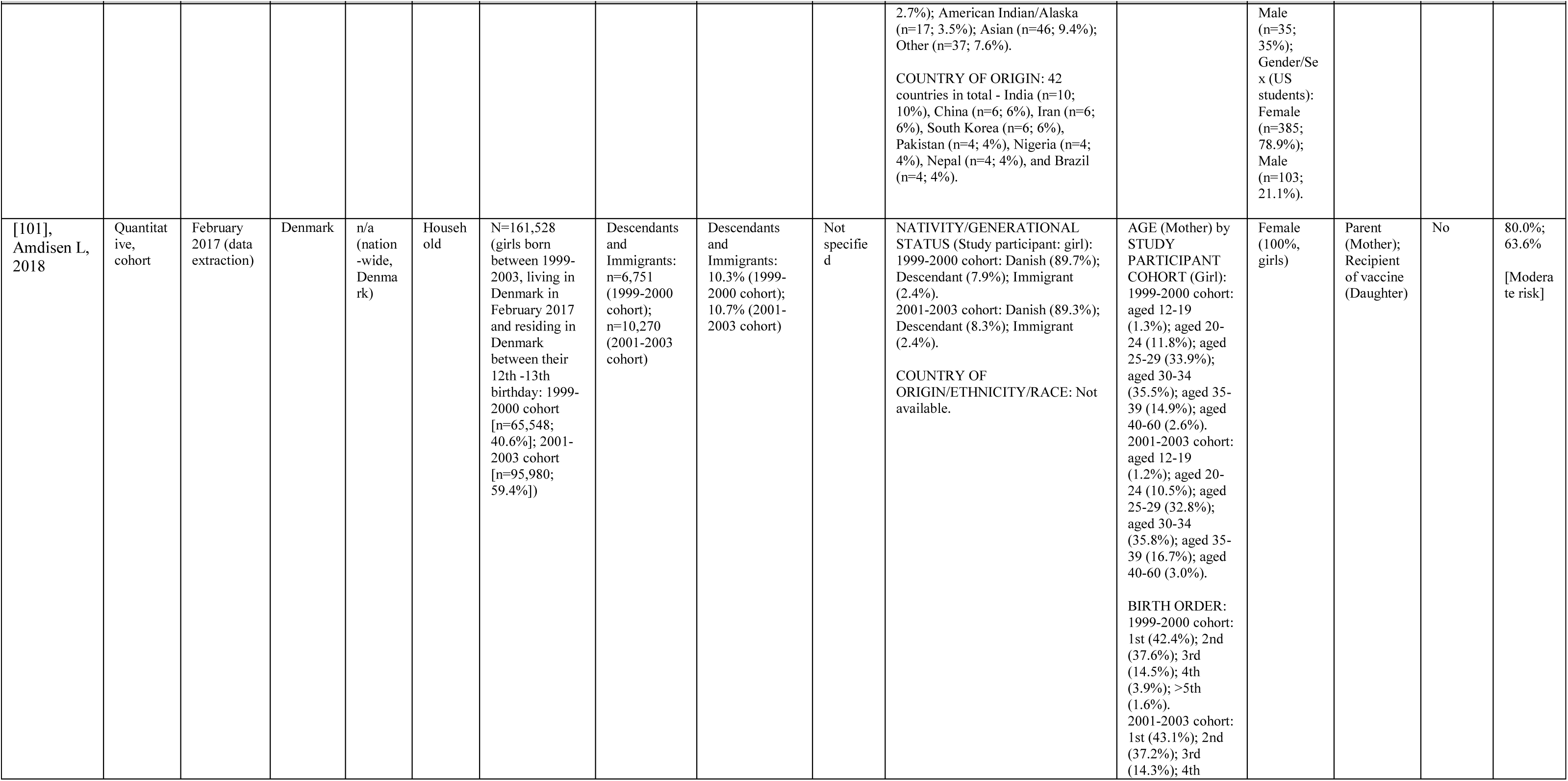
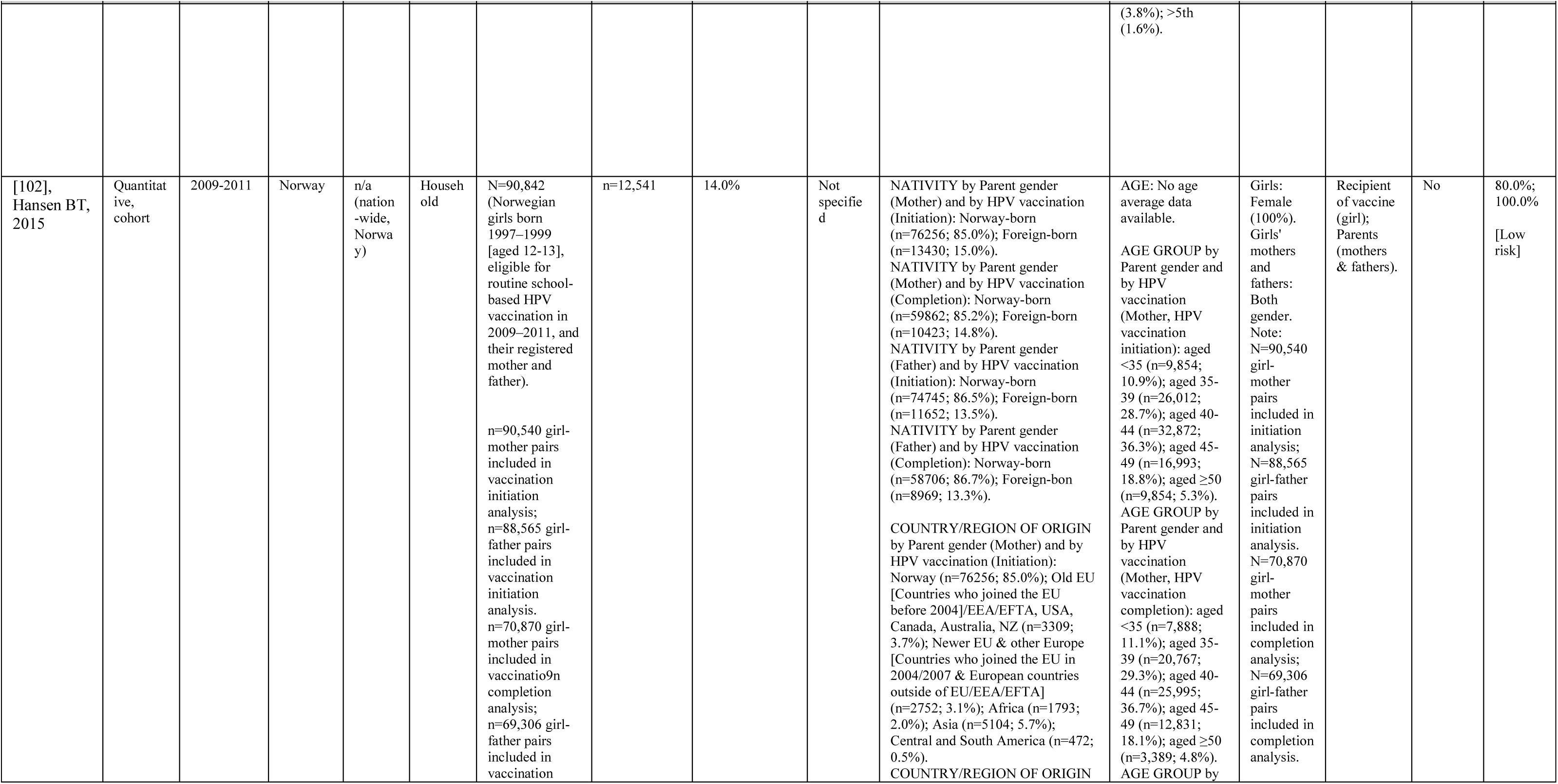
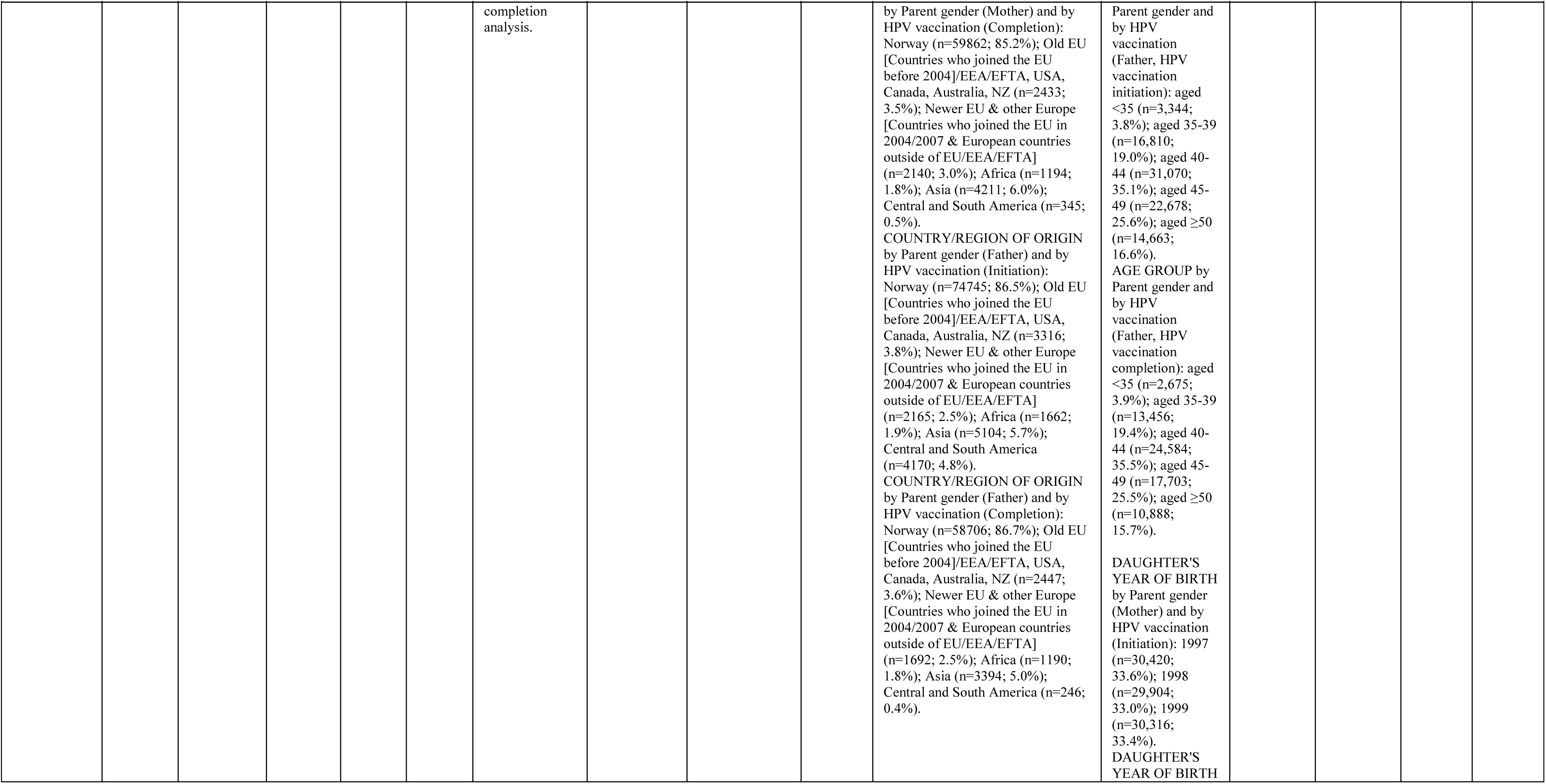
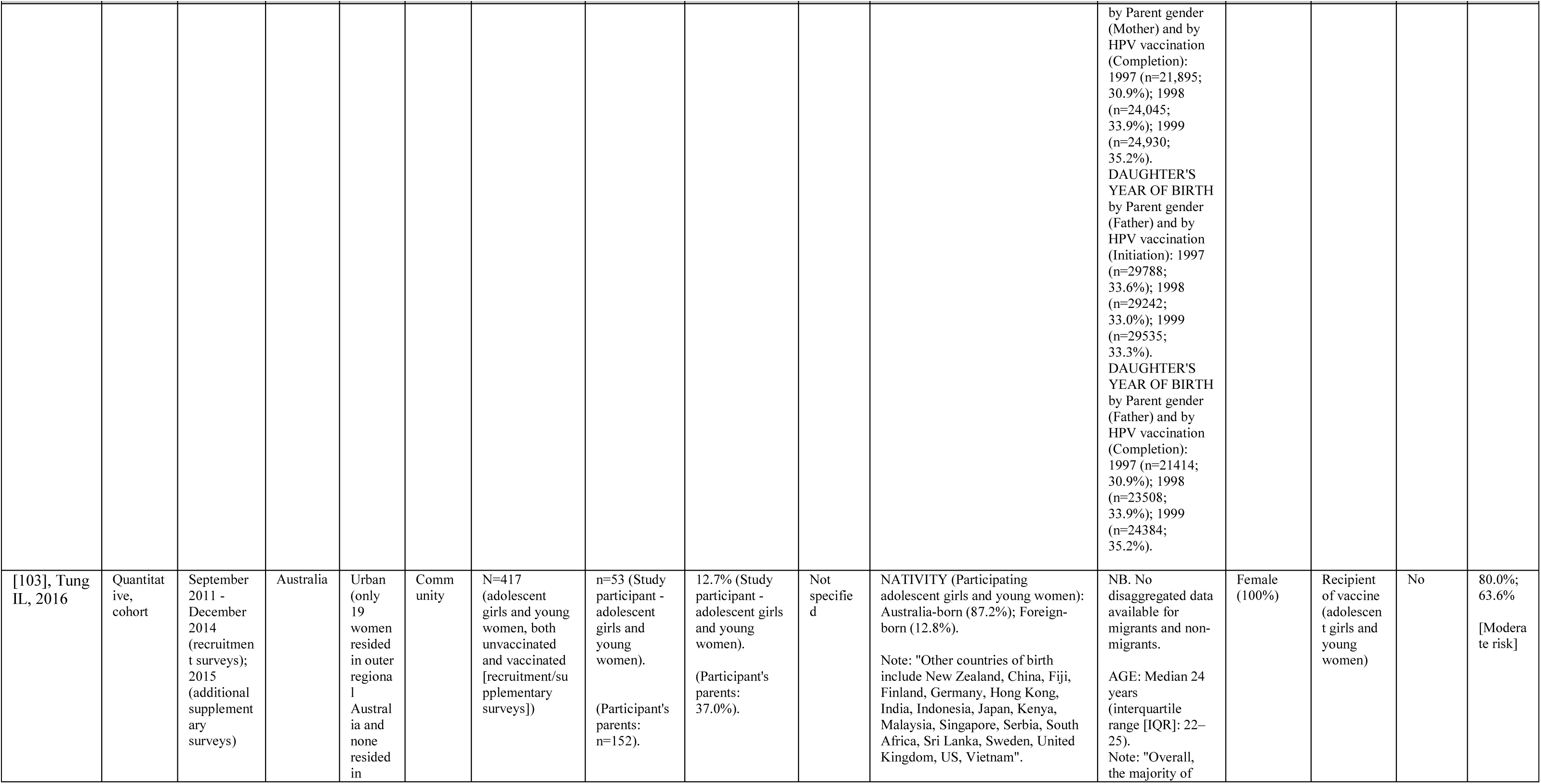
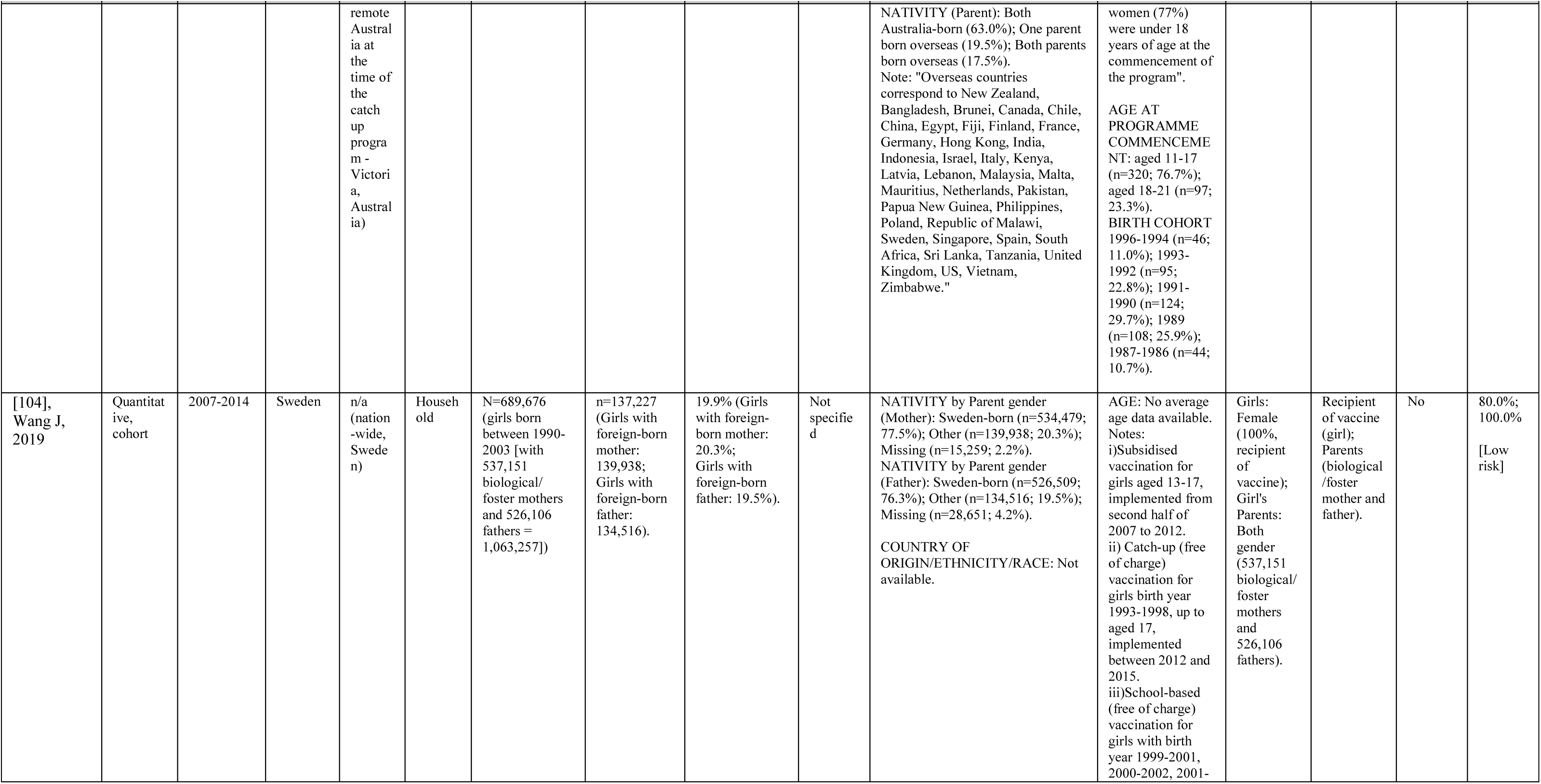
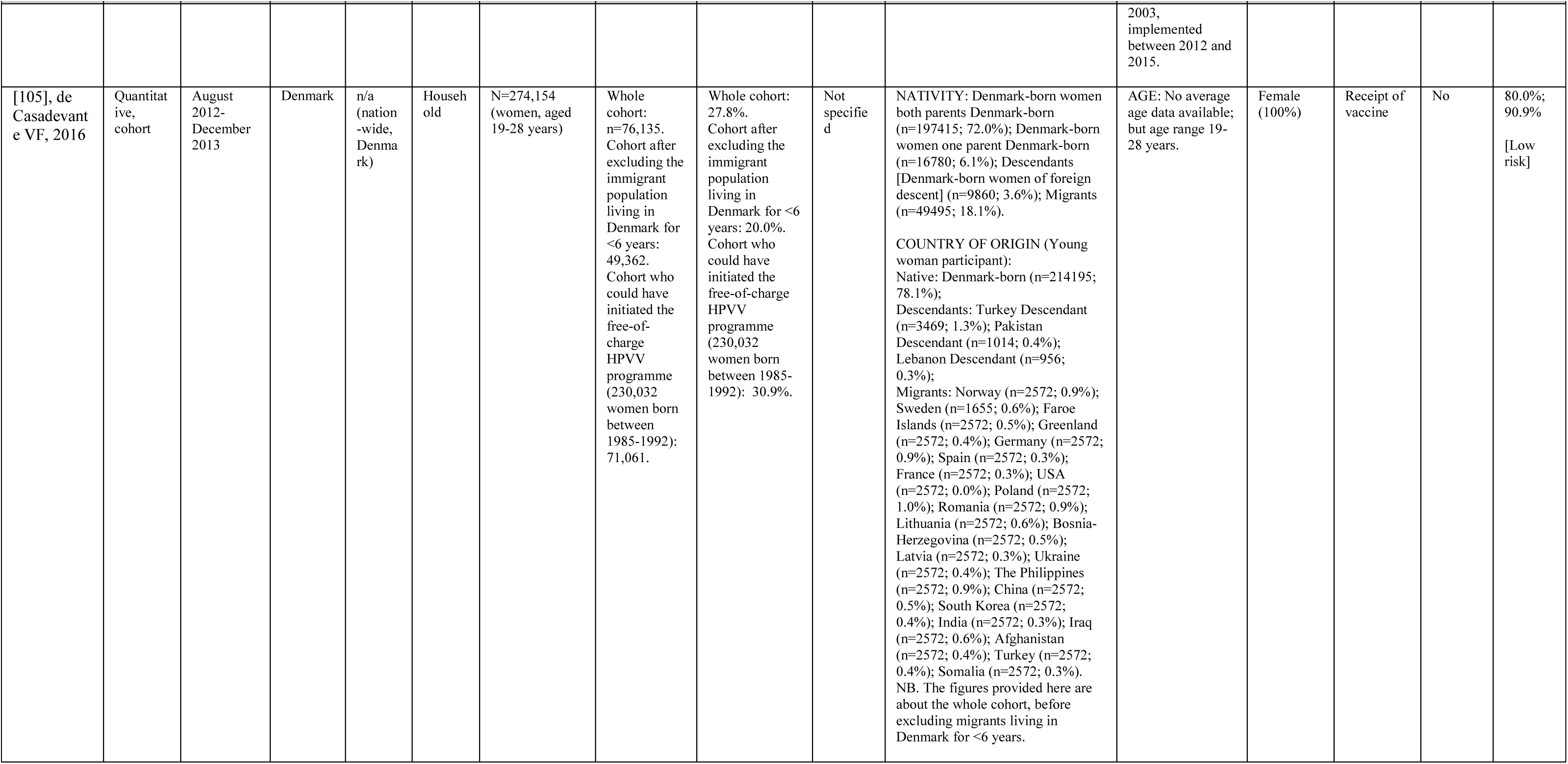
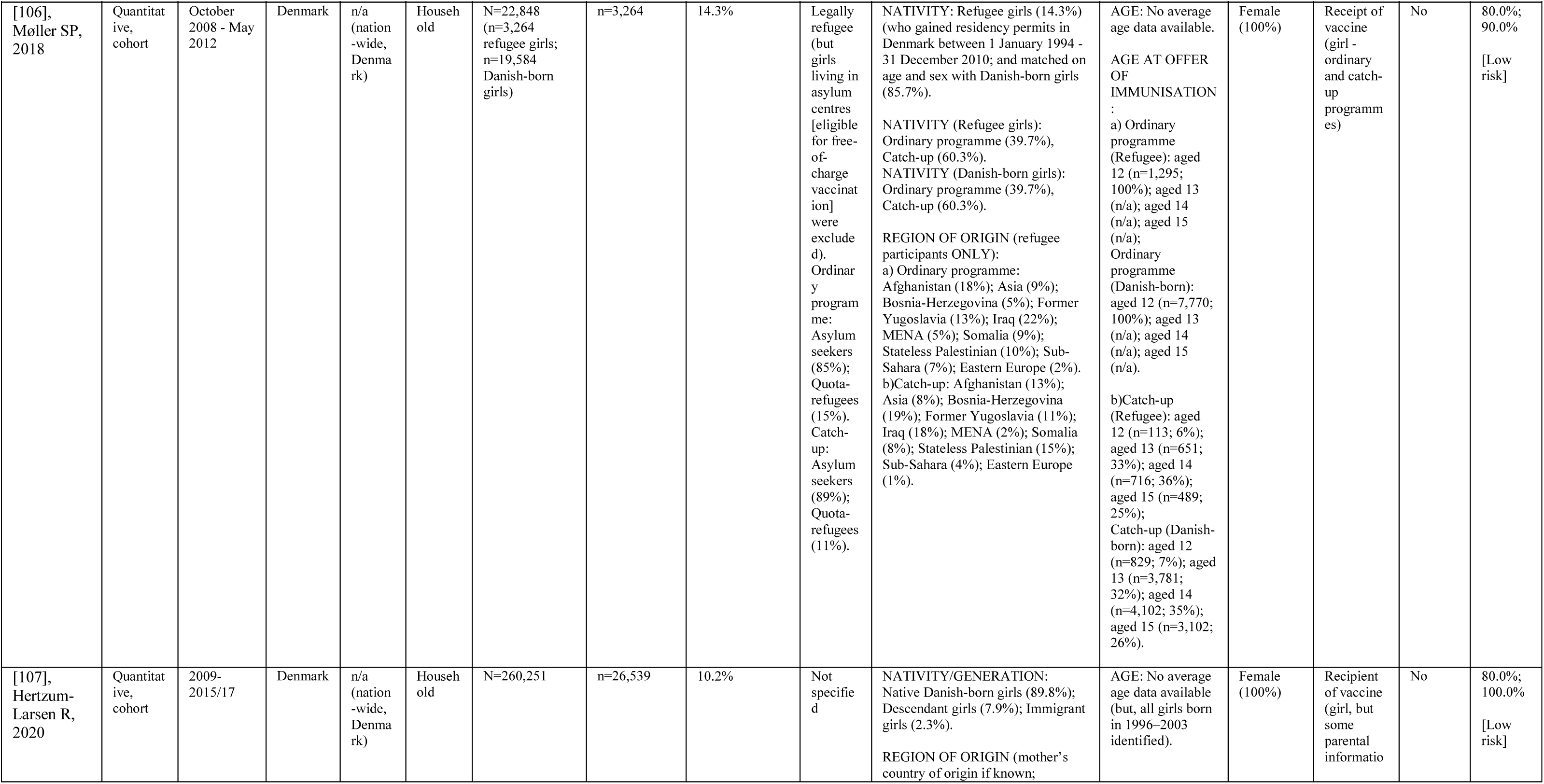
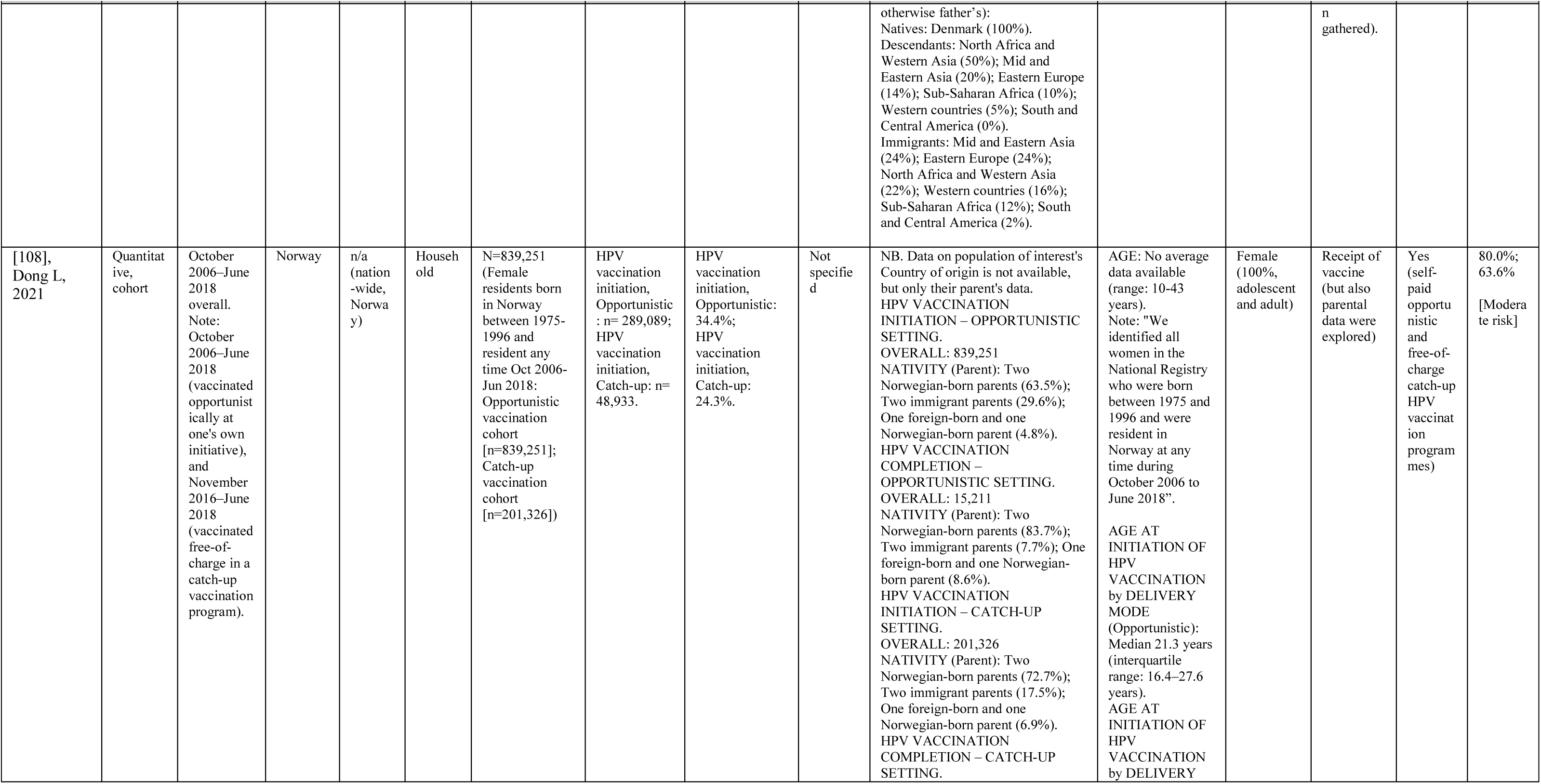
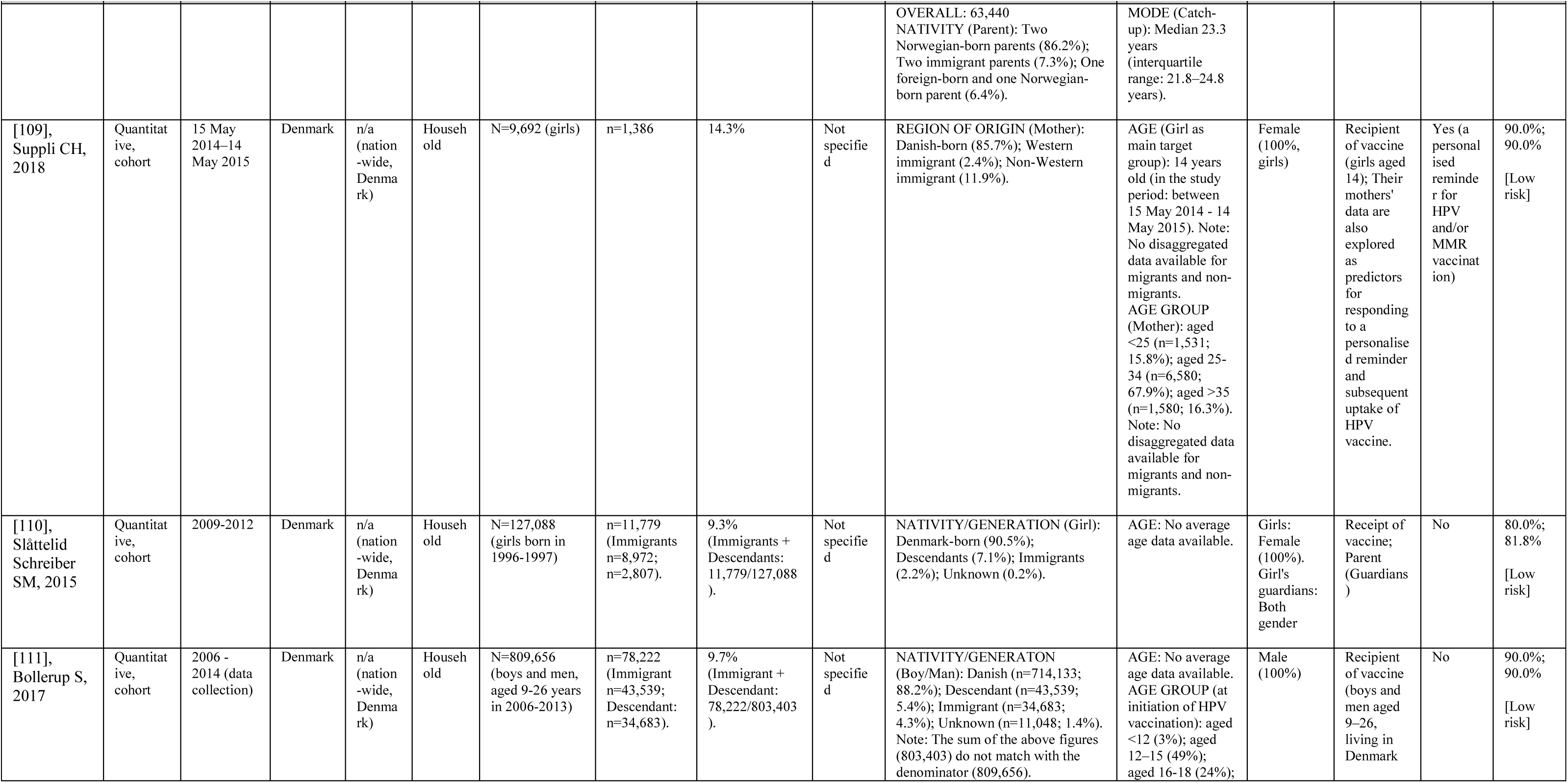
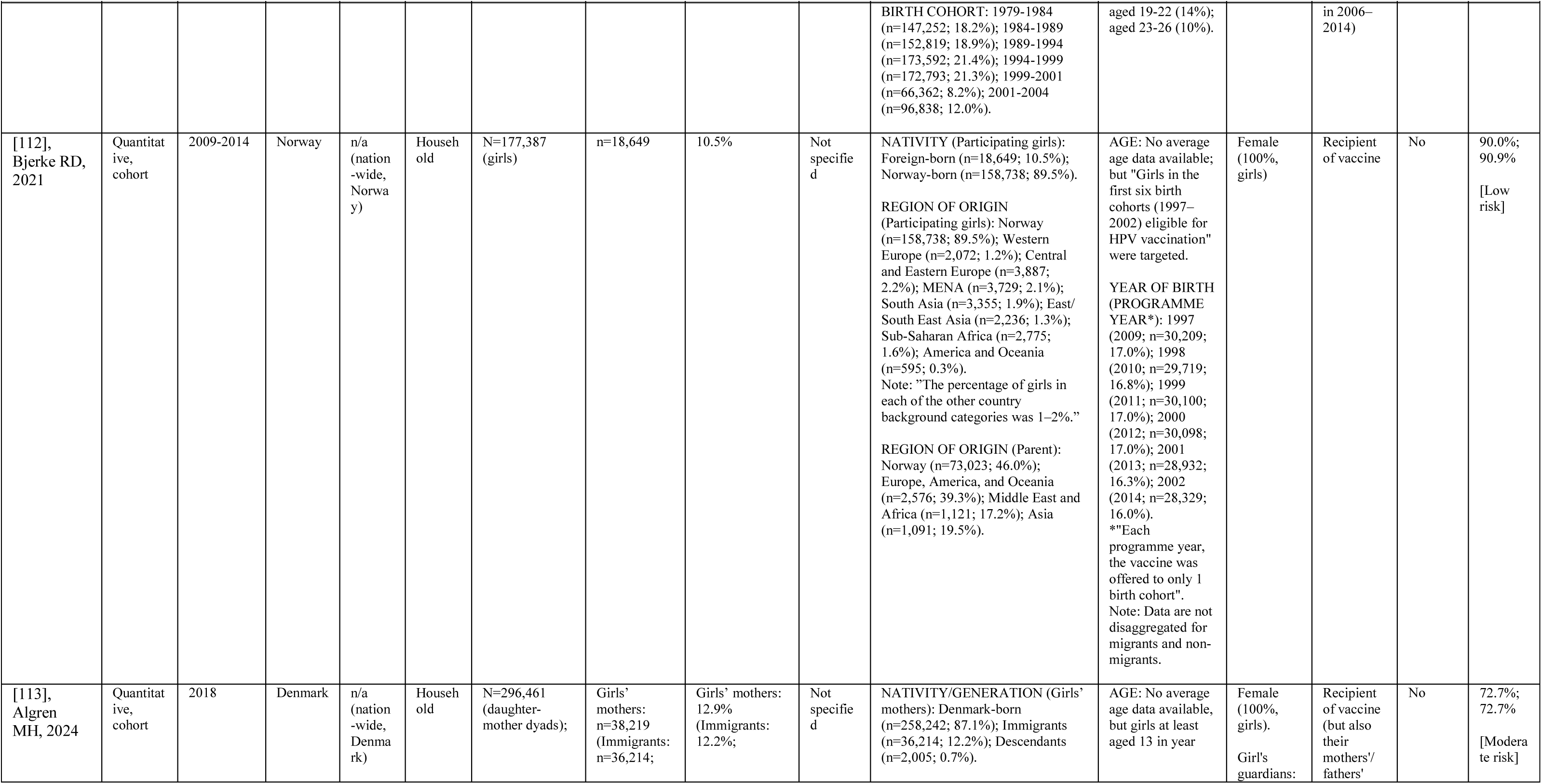
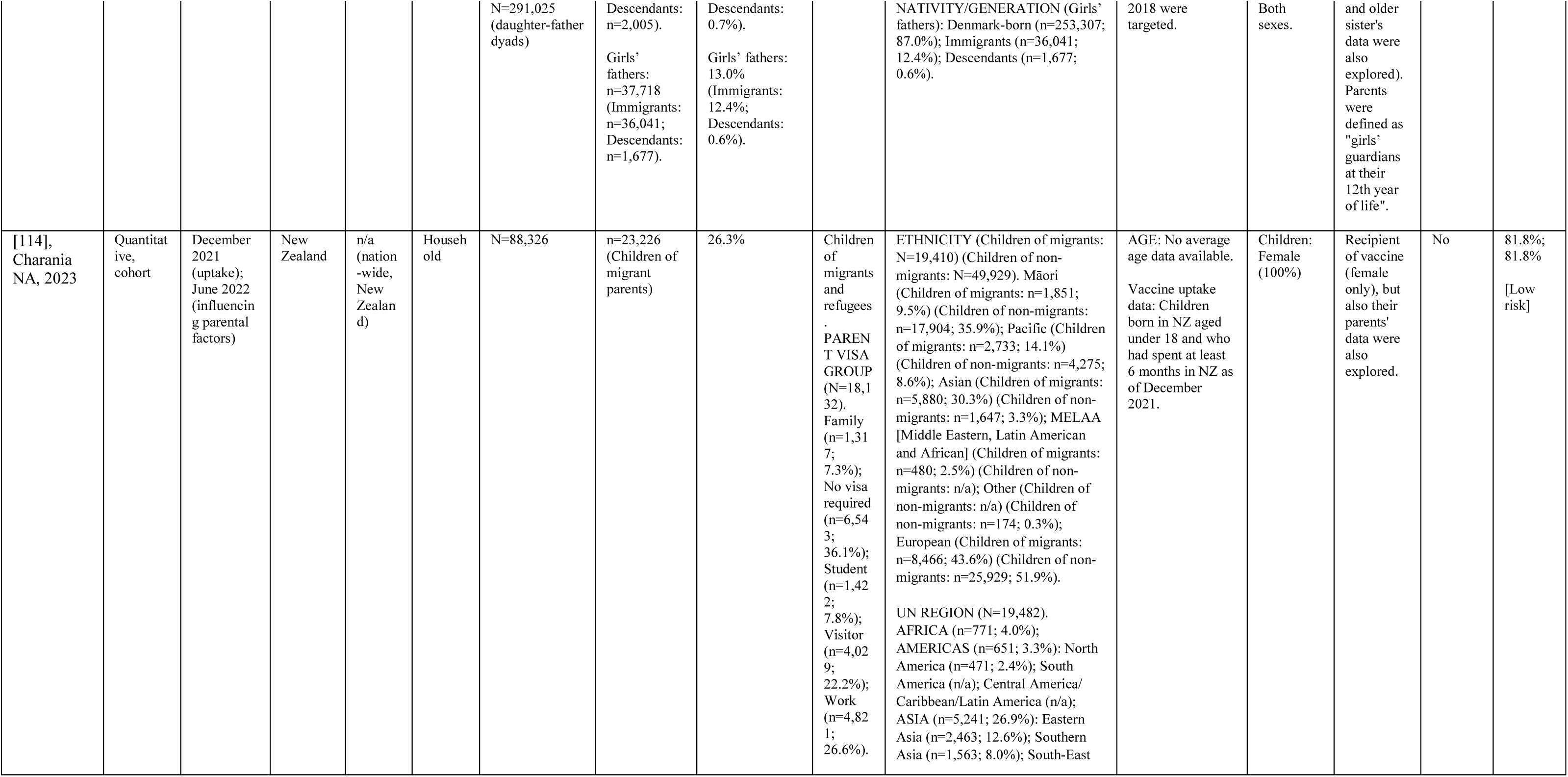
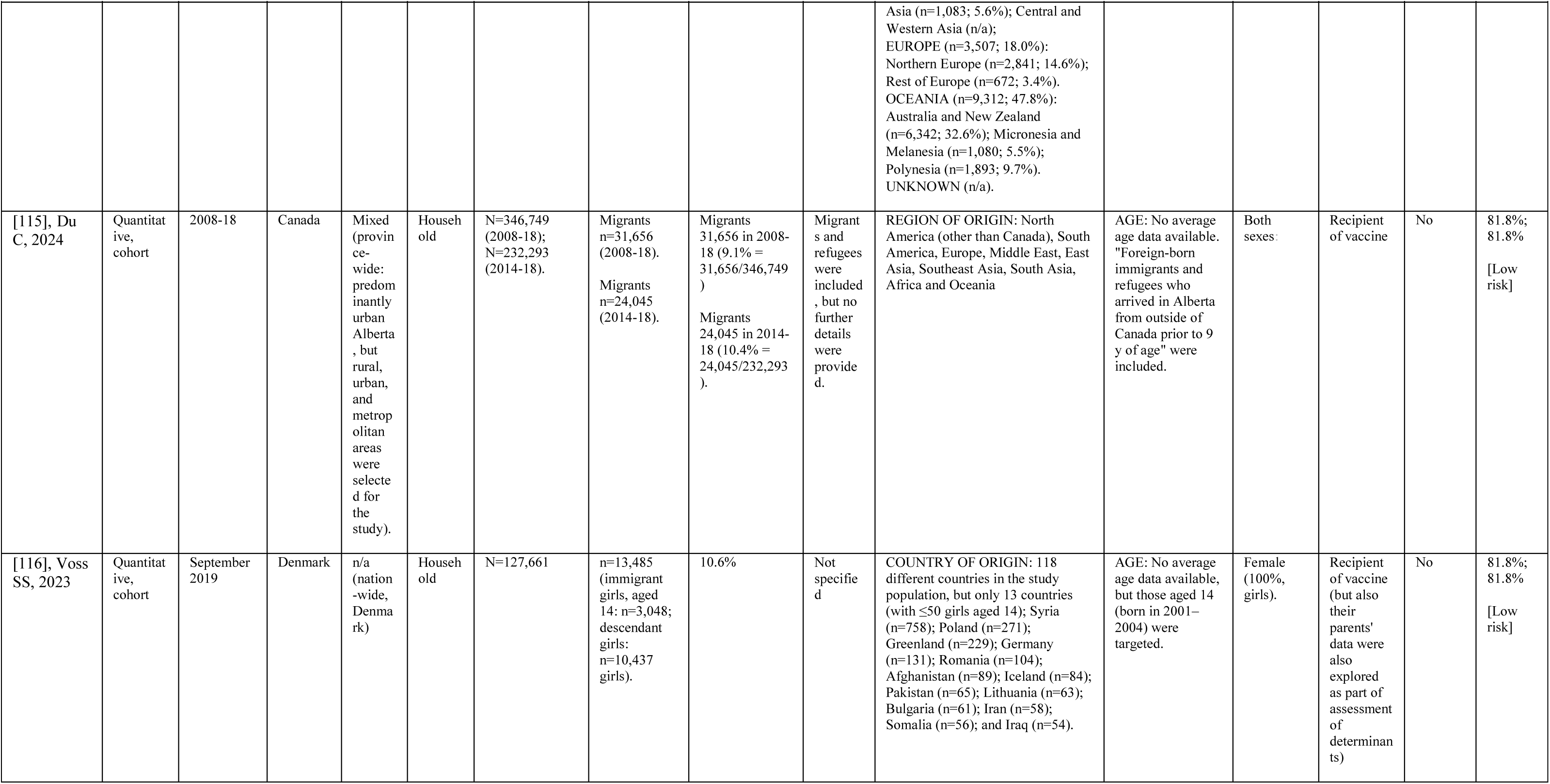
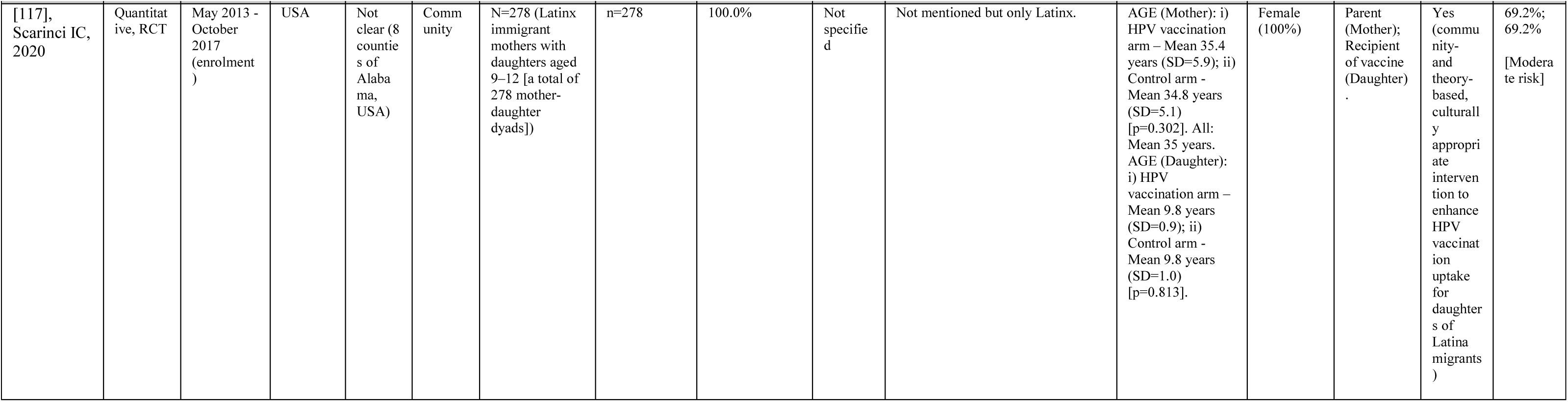
Detailed description of included studies (N=117)

**Table S4.**
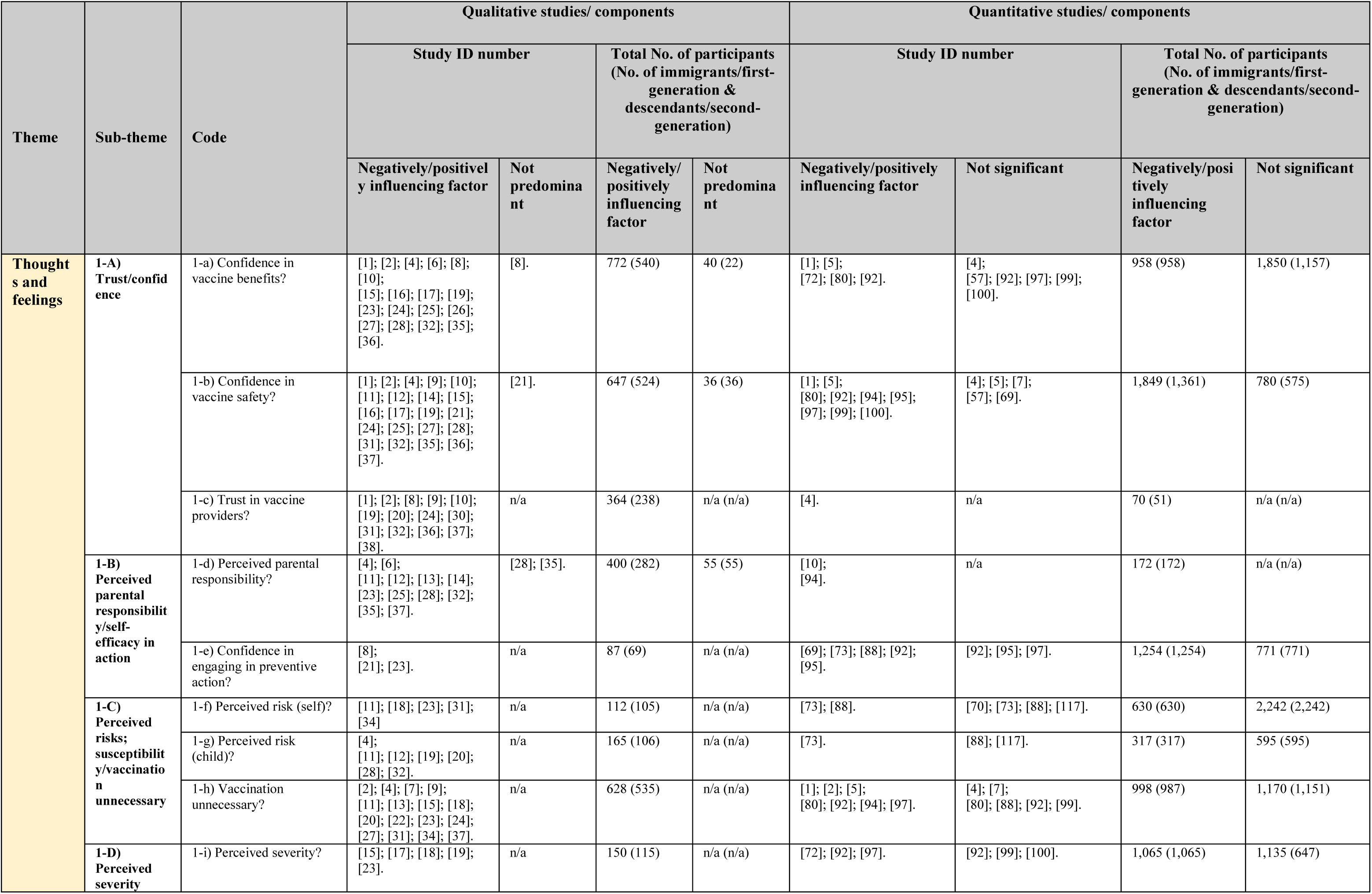
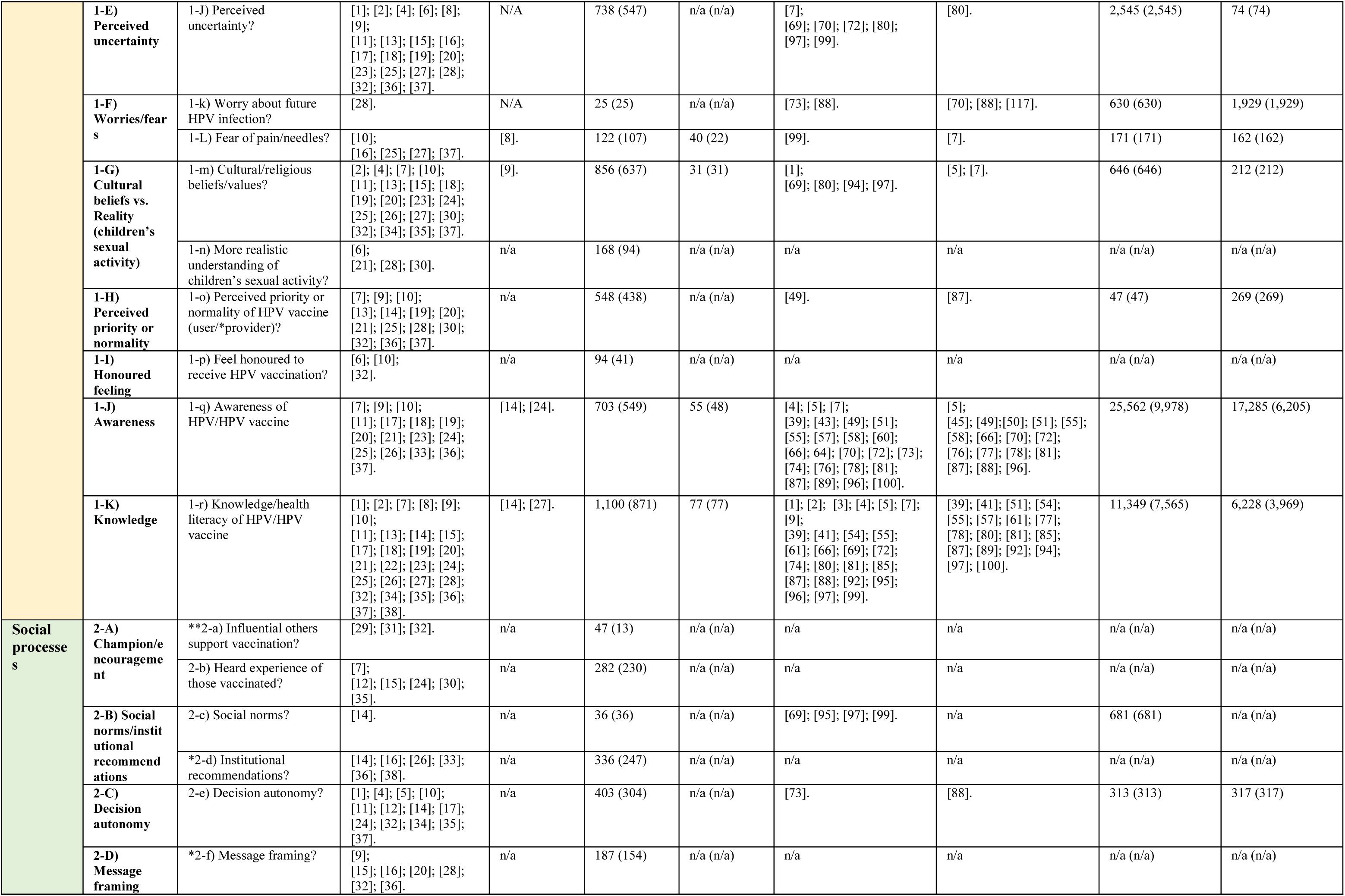
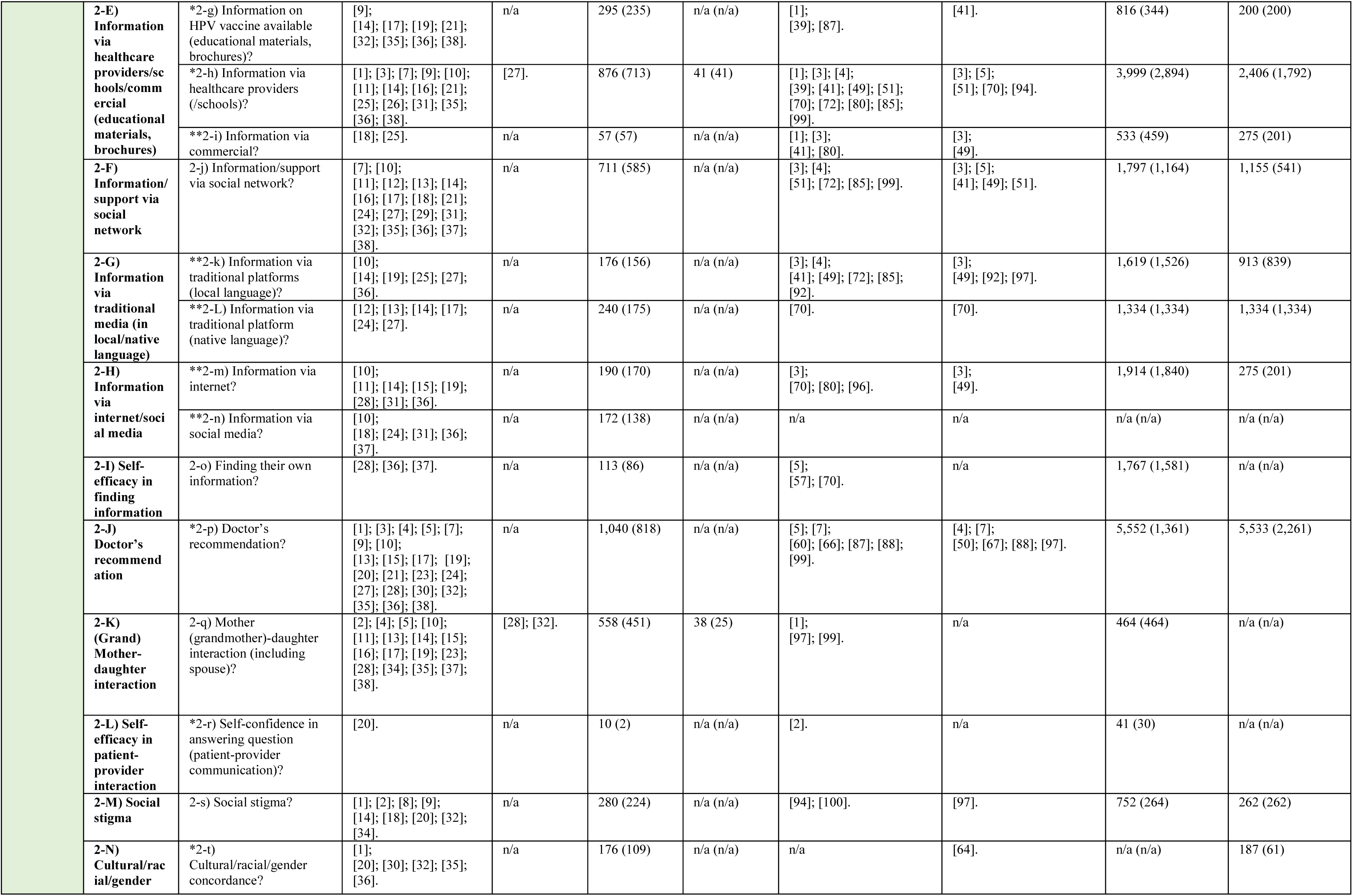
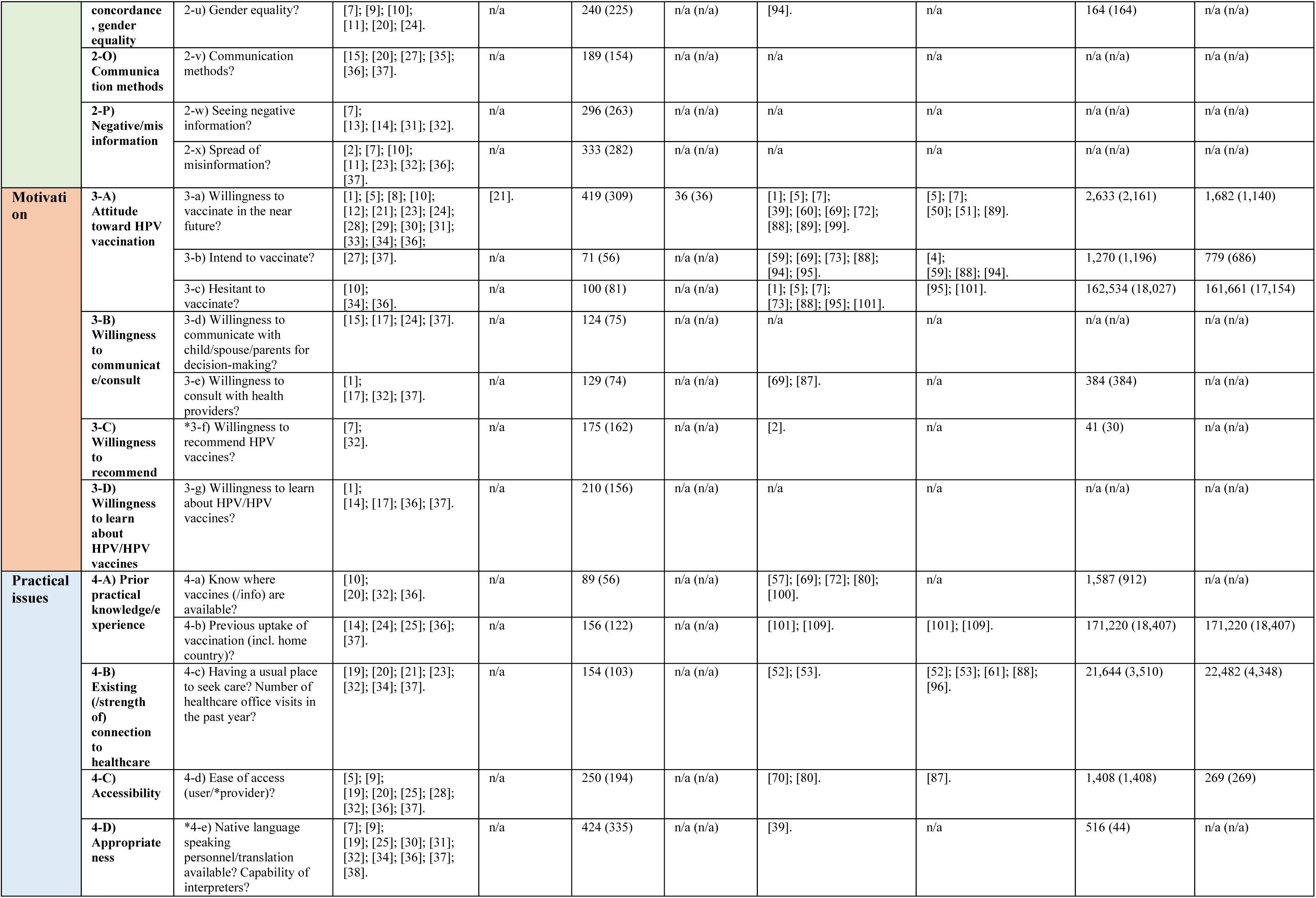
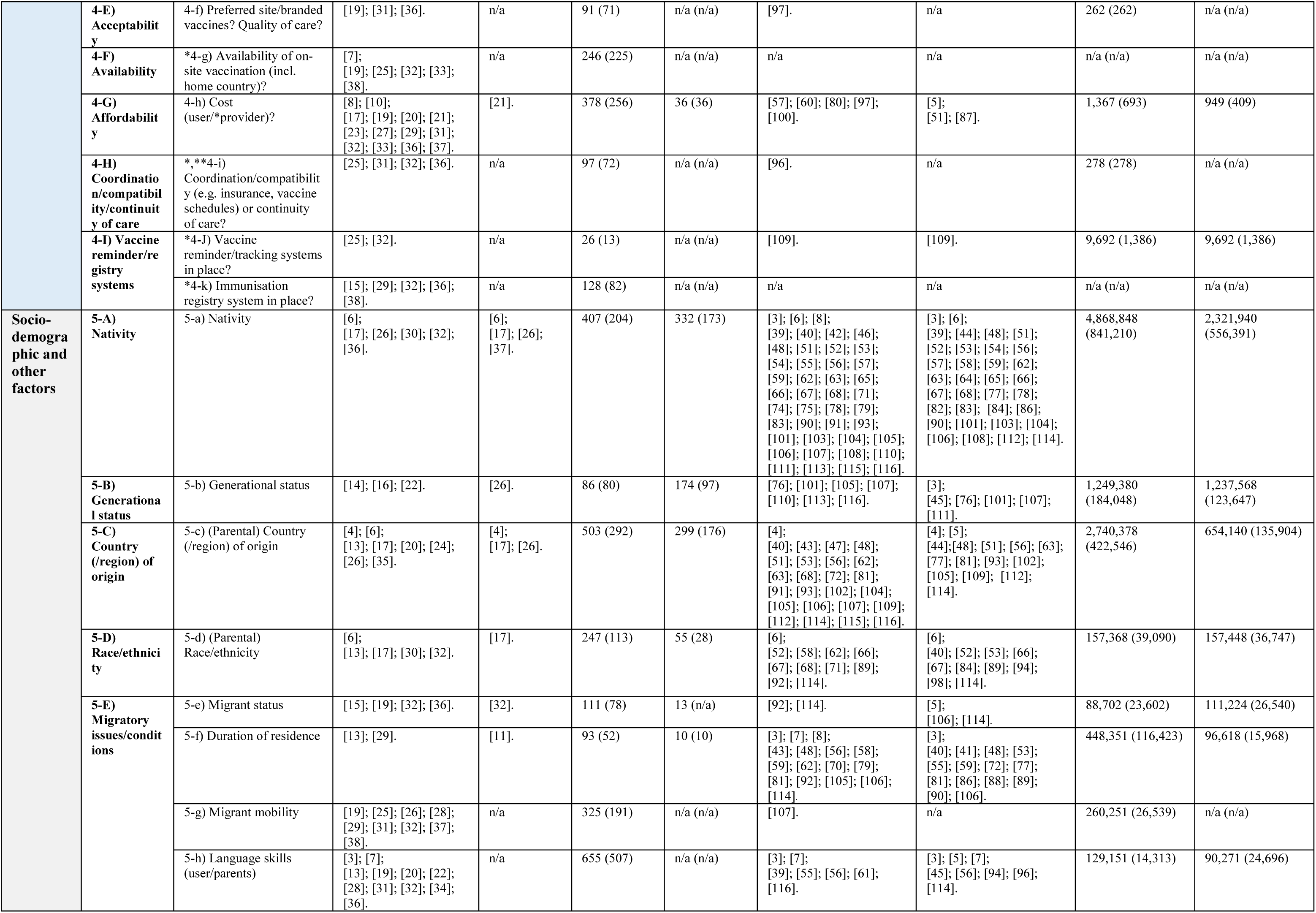
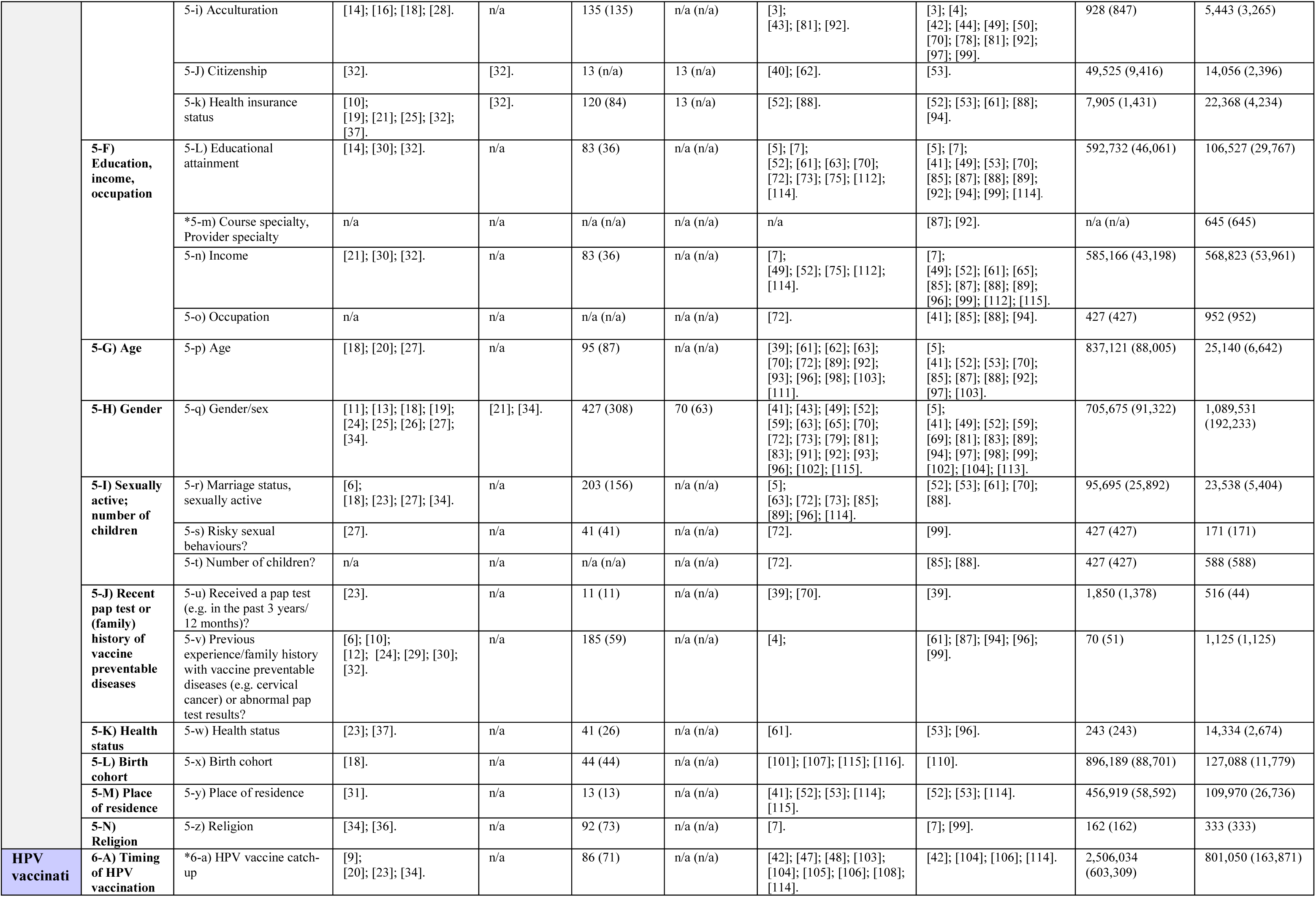
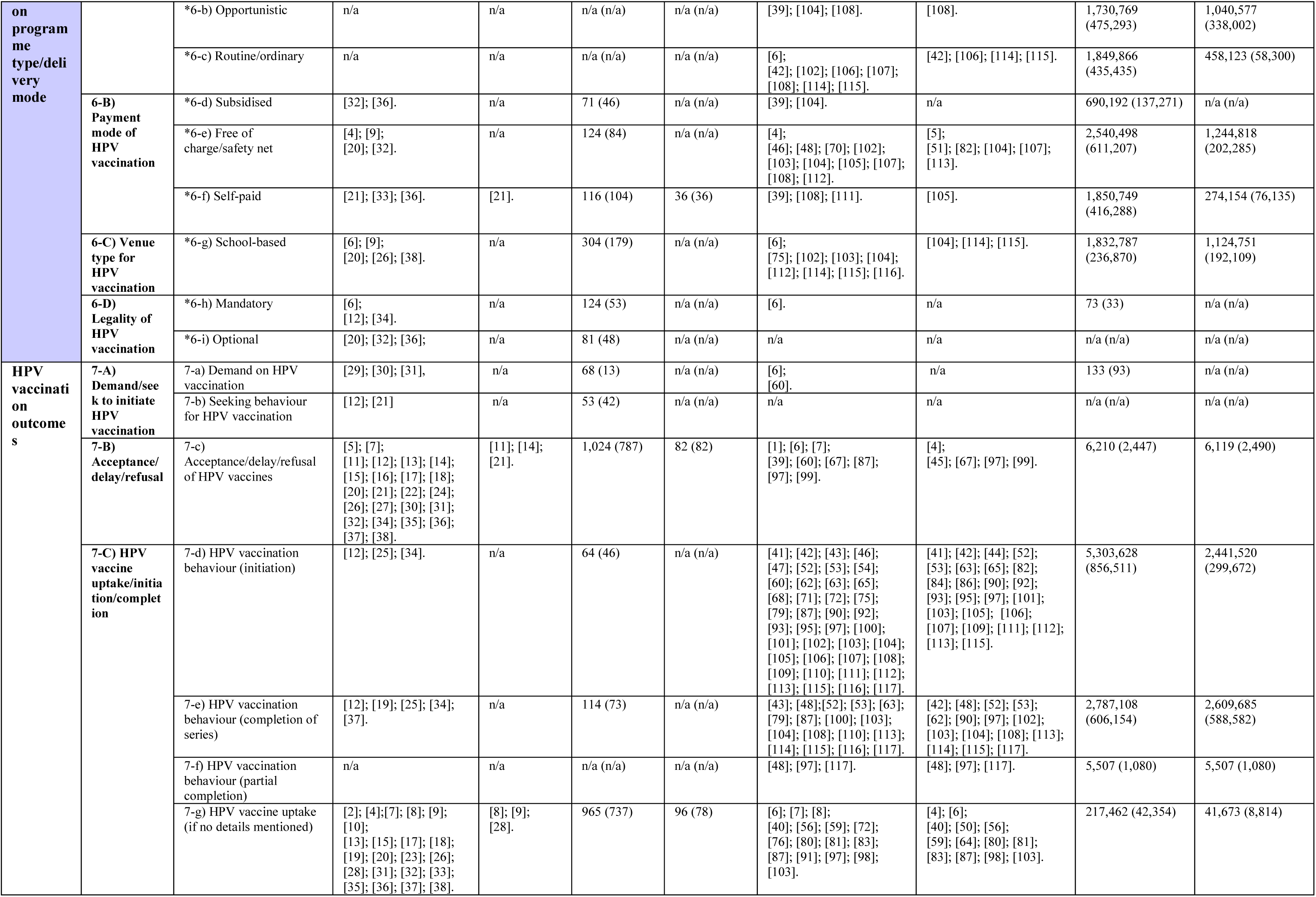
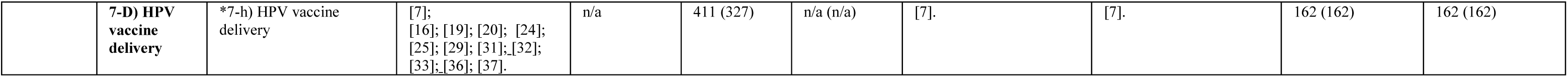
A summary of qualitative and quantitative findings.

**Figure S1.**
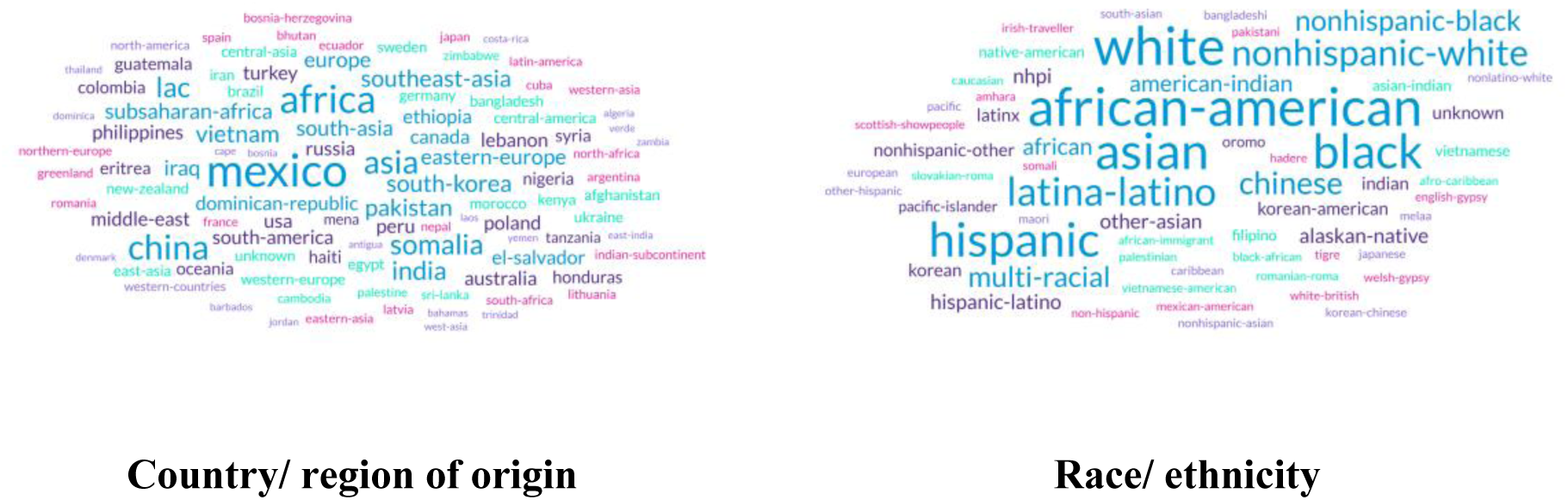
Country/region of origin and Race/ethnicity of participants [117 studies, 5,638,836 participants in total]. MENA: Middle East and North Africa; MELAA: Middle Eastern, Latin American and African; LAC: Latin America and the Caribbean; NHPI: Native Hawaiian & Pacific Islander

